# Genetic analysis of lung cancer reveals novel susceptibility loci and germline impact on somatic mutation burden

**DOI:** 10.1101/2021.04.26.21254132

**Authors:** Aurélie AG Gabriel, Joshua R Atkins, Ricardo CC Penha, Karl Smith-Byrne, Valerie Gaborieau, Catherine Voegele, Behnoush Abedi-Ardekani, Maja Milojevic, Robert Olaso, Vincent Meyer, Anne Boland, Jean François Deleuze, David Zaridze, Anush Mukeriya, Beata Swiatkowska, Vladimir Janout, Miriam Schejbalová, Dana Mates, Jelena Stojšić, Miodrag Ognjanovic, the ILCCO consortium, John S Witte, Sara R Rashkin, Linda Kachuri, Rayjean J Hung, Siddhartha Kar, Paul Brennan, Anne-Sophie Sertier, Anthony Ferrari, Alain Viari, Mattias Johansson, Christopher I Amos, Matthieu Foll, James D McKay

## Abstract

Germline genetic variants are involved in lung cancer (LC) susceptibility. Previous genome-wide association studies (GWAS) have implicated genes involved in smoking propensity and DNA repair but further work is required to identify additional LC susceptibility variants and to investigate LC disease development dynamics.

We have undertaken a family history-based genome-wide association (GWAx) study of LC, analysing 48,843 European cases with a parent/sibling with LC compared to 195,387 controls from the UK Biobank. This was meta-analysed with previously described LC GWAS results. We performed Polygenic Risk Scores (PRS) analyses and further evaluated the PRS influence on the somatic environment in exome (N=736) and genome sequencing (N=61) profiled cohorts.

Eight novel loci were identified including DNA repair genes (*CHEK1, MDM4*), metabolic genes (*CYP1A1*) and variants that were also associated with smoking propensity, such as both subunits of the neuronal α4β2 nicotinic acetylcholine receptor (*CHRNA4* and *CHRNB2)*. PRS analysis demonstrated that variants related to eQTLs and/or smoking propensity are enriched for susceptibility variants, including variants below genome-wide significant threshold. PRS of LC variants related to smoking propensity were associated with somatic mutation burden in two case cohorts, with individuals with higher polygenic genetic risk having increased numbers of somatic mutations in their lung tumours.

This study has expanded the number of susceptibility loci linked with LC and provided insights into the molecular mechanisms by which these susceptibility variants contribute to the development of lung cancer.

## INTRODUCTION

Lung cancer (LC) is the most common cause of cancer-related deaths worldwide. While most LC risk is attributable to exposure to tobacco smoke, a genetic basis for LC susceptibility was initially identified from familial aggregation studies after accounting for personal smoking habits ^1–3^, segregation based analyses^4^ and twin studies.^5^ Genome-wide association studies (GWAS) have identified multiple lung cancer susceptibility loci in genes related to propensity to smoke tobacco (*CHRNA5, CHRNA3, CHRNB4, CYP2A6*)^6–8^, DNA repair (*CHEK2, BRCA2, ATM*)^9–11^ and genes related to telomere regulation (*TERT, RTEL, OBFC1*)^12,13^ as well as many loci where the target genes are less obvious.^13^

While traditional GWAS approaches continue to expand in size, novel analytical approaches can leverage existing data from large, genotyped biorepositories to identify additional loci. An example is the genome-wide association by proxy (GWAx) approach, which considers 1^st^ degree relatives previously diagnosed with the given trait of interest as “proxy” cases and individuals without relatives with that given trait as “proxy” controls.^14,15^ In the current study, we undertook a GWAx of lung cancer in the UK Biobank and combined this with the largest GWAS of LC undertaken to date.^13^ Furthermore, we constructed Polygenic Risk Scores (PRS) with variants related to LC and used these scores to investigate the influence of these germline susceptibility variants on the somatic mutation burden in two independent cohorts.

## MATERIALS AND METHODS

### Cohorts

A detailed description of each dataset (Transdisciplinary Research for Cancer in Lung (TRICL, the traditional LC GWAS),^13^ UK Biobank (GWAx and a subset left out of the GWAx, forming the germline PRS test set)^16^, The Cancer Genome Atlas (TCGA, somatic mutations and signatures analysis (https://www.cancer.gov/tcga)) and the GeniLuc cohort (somatic mutation and signatures replication cohort (unpublished)) can be found in the supplementary information.

### Genome-wide association by using a family history and genetic correlations analysis

The UK Biobank resource was accessed under project number 15825. The sample selection process and variant filtering from the UK Biobank is detailed in Supplementary Table 1 along with the UK biobank data ID fields. We performed a traditional GWAS on the family history status (GWAx) of LC and adjusted the betas and standard error as described previously.^14^ Individuals diagnosed with lung cancer directly were not included in the GWAx. All samples were reported as having a European ancestry (confirmed with ancestry inferred by genetic profile) with non-European individuals excluded from the study due to different genetic architecture. Consent and ethics were approved for all cohorts used. Genetic correlation analysis was undertaken using the LDSC package.^17^ Summary statistics were obtained from the LC traditional GWAS which has been previously published elsewhere. ^13^ Each summary statistic file from the Sequencing Consortium of Alcohol and Nicotine (GSCAN) consortium (with UK Biobank samples removed) along with the traditional LC GWAS and the GWAx were tested for genetic similarity using LDSC regression. ^18^

Both the LC family history GWAx and the LC GWAS were meta-analysed using a fixed effect model. LD clumping was undertaken using PLINK (R^2^ < 0.1 and 10,000 kb). eQTL analysis was performed using GTEx version 8 data for both lung and brain tissues containing all variant and gene pairs. Coffee intake and forced vital capacity (FVC) summary statistics were obtained from the Benjamin Neale UK Biobank work (http://www.nealelab.is/uk-biobank/). To estimate the colocalization between genetic associations of two traits at a given locus, we calculated the Bayesian posterior probability (PP4) for colocalisation of two datasets for the H_4_ (one shared variant across both traits),^19^ by firstly calculating the log bayes factor for each SNP in each dataset, then the PP4 was calculated by the COLOC package in python (https://github.com/anthony-aylward/coloc).

### Mutation burden analysis

The somatic mutations from the TCGA samples were retrieved from the study of Ellrott *et al 2018* ^20^ excluding individuals flagged for QC issues (see supplementary methods for further QC details). Germline genotypes were derived from Affymetrix 6.0 arrays.

For the GeniLuc cohort, 61 lung cancer patients were identified from central and eastern Europe as described previously.^13^ Subsequent to histopathological review, to ensure appropriate tumour purity, DNA was extracted from normal material (blood) and the lung tumour resection. Whole genome sequencing (WGS) was undertaken using PCR free whole genome library preparation and sequenced to a depth of 30X for the paired tumour normal for each patient using an Illumina HiSeq X 5 DNA sequencer at the National Center of Human Genomic Research (CNRGH) laboratory in Paris, France. Raw sequencing data was processed by inhouse Nextflow pipelines (https://github.com/IARCbioinfo). Somatic mutations were defined using Mutect2 and germline calls using Strelka2.^21,22^ For germline genotypes from WGS tissue, PRS SNPs were extracted from VCF files and put in the PLINK BED format.^23^ Individual PRS scores were generated using PRsice2 from the normal calls.^24^

### Mutational signatures computation

In order to compute mutational signatures, mutational matrices for each mutation type (Single Base Substitution (SBS), Doublet Base Substitution (DBS), and small Insertion and Deletion (ID)) were generated using SigProfilerMatrixGenerator (v1.1.20) with default parameters.^25^ Mutational signatures were then extracted with SigProfilerExtractor (v1.0.17) from the TCGA-WES (LUAD and LUSC) samples and GENILUC-WGS lung cancer cohorts, separately, using the default options.^26^ SigProfilerExtractor extracted *de novo* signatures for each context (SBS96, DBS78, and ID83) and the optimum number of *de novo* signatures (suggested solution method) were decomposed into COSMIC (version 3.1) reference signatures. Previously reported smoking tobacco-related signatures, SBS4, DBS2, and ID3 (ID83A and ID83B), and the absolute mutation counts for each COSMIC signature per sample were assessed.

### Statistical analysis

For the GWAx analysis, association testing was performed using a logistic regression model using the --glm function in PLINK 2.0 on European ancestry individuals. Each model was adjusted by age at recruitment, sex, array type, and the first 5 principal components that define genetic ancestry (PCs) to account for population structure. The meta-analysis was done using METASOFT using a fixed-effects model based on an inverse-variance-weighted effect size.^27^ Germline PRS analysis in the UK Biobank samples was performed using a logistic regression model after standardising raw PRS scores. Covariates that were used in the model included sex, array type, age of recruitment and the first 5 principal components from genetic inferred ancestry. Odd ratios for PRS are given as a one unit increase per a standard deviation in score. For the analysis of PRS associations with mutational signatures, a model diagnostic was used to compare a linear model, negative binomial model and a Quasi-Poisson model due to frequent zero-inflation for mutational signatures. Covariates included in the models were age, gender, the 5 first principal components resulting from Eigenstrat and tumour purity. In TCGA, a categorical variable indicating the cohort type was included in the model as appropriate.

## RESULTS

### The 8 novel susceptibility loci

The family history GWAS (GWAx) on 48,843 self-reported “family history lung cancer cases” and 197,029 “controls” (Supplementary Table 1) identified five loci (5p15.33, 6p21.32, 12p13.33, 13q13.1 and 15q25.1) that had previously been discovered from the traditional GWAS (Supplementary Table 2 and Supplementary Figure 2) and LDSC confirmed a strong relationship between both GWAx and the GWAS (r_g_ = 1, se = 0.066, p = 4.0 × 10^−52^) supporting the utility of this approach (see supplementary material document for further details). Meta-analysis between the GWAx and the traditional LC GWAS ^13^ identified 65 variants that achieved a P-value of less than 5×10^−8^ across 21 distinct genomic loci defined by cytoband (Figure 1), after LD clumping genetic variants (Supplementary Table 2). At previously described lung cancer susceptibility loci, the meta-analysis also identified independent (R^2^ < 0.1) low-frequency (MAF < 0.05) variants associated with lung cancer at 5p15.33 (rs35812074), 19q13.2 (rs1801272), 15q25.1 (rs2229961, rs8192479, rs151118057) and at 12p13.33 (rs7487683) in addition to previously described common genetic variants (Supplementary Table 2). At 13q13.1, where a rarer lung cancer susceptibility allele has previously been described (rs11571833, K3326X *BRCA2*, MAF = 0.01), a common susceptibility allele was noted (rs11571734, MAF = 0.28).

**Figure 1:**
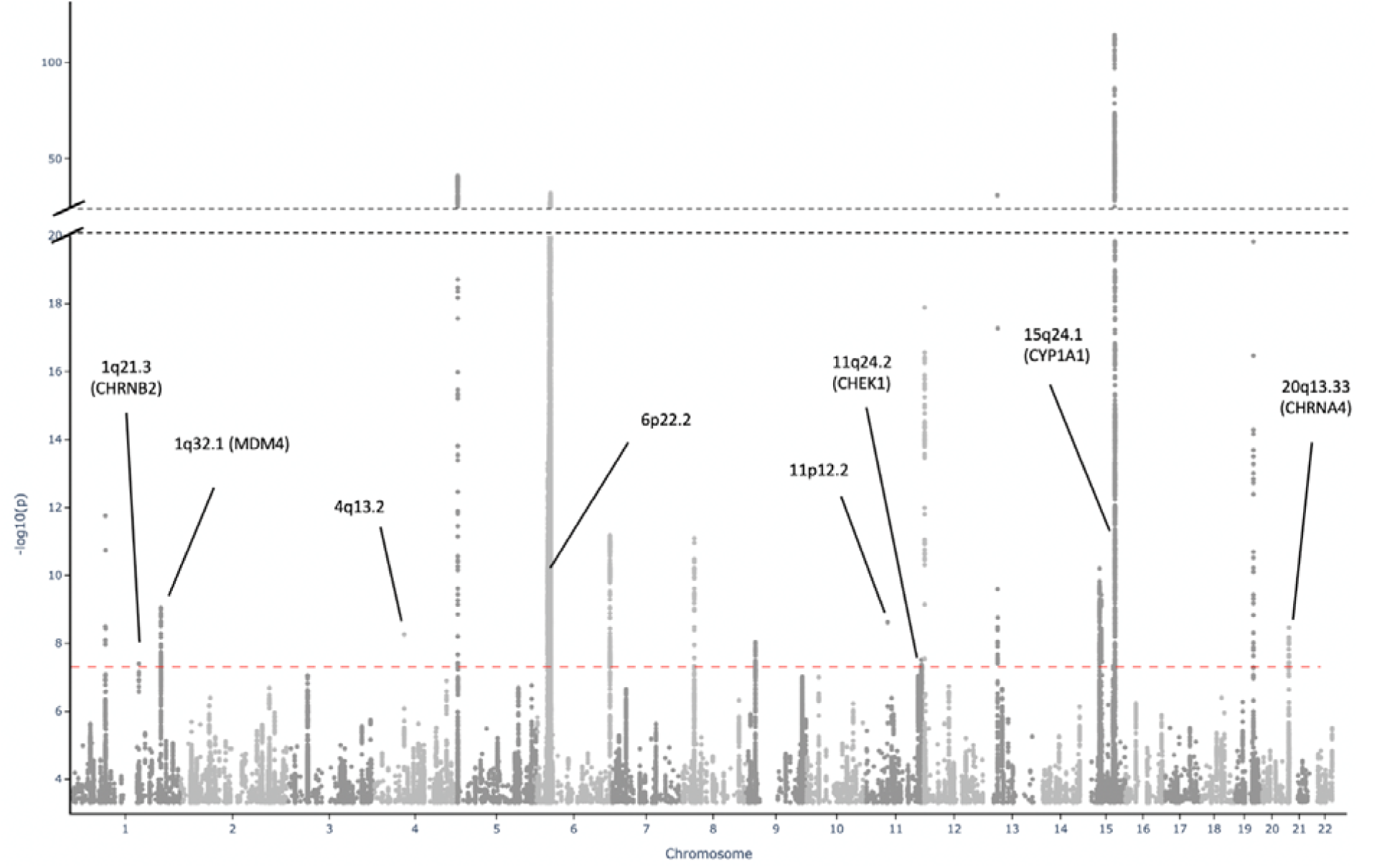
Manhattan plot of the meta-analysis of genome-wide by proxy (GWAx) with genome-wide association study (GWAS) into lung cancer. The Manhattan plot displays the results of the meta-analysis of the GWAx (48,843 proxy cases and 197,029 proxy controls without a family history of any cancer) and the GWAS (29,266 cases and 56,450 controls) with the new novel loci highlighted in black with the likely candidate gene name presented. This meta-analysis discovered 65 novel loci across 21 cytoband regions. The x-axis is the chromosome position across the autosomal chromosomes, with the Y-axis containing the association level displayed as the -log10(P-value), derived by a multivariate logistics regression model. The red dotted line displays the genome-wide significance threshold (5×10^−8^)

Eleven lung cancer susceptibility variants at eight loci have not previously associated with lung cancer at genome wide (GW) significance (Table 1). Of these, the lung cancer susceptibility variants at 1q21.3-rs78062588, 6p22.2-rs7766641 and 20q13.33-rs11697662 were also associated at GW significance with traits related to propensity to smoke tobacco (Supplementary Table 2). The sentinel variants at 1q21.3-rs78062588 and 20q13.33-rs11697662 are eQTLs for the nicotinic acetylcholine receptors (nAChRs) subunits *CHRNB2* and *CHRNA4* (Supplementary Table 2 and Supplementary Table 3). At 6p22.2, LC susceptibility loci were noted (Supplementary Table 2), typified by two sentinel variants, rs6913550 and rs7766641. rs7766641 was also associated with propensity to smoke, whereas curiously rs6913550 was not (Supplementary Table 2).

**Table 1:**
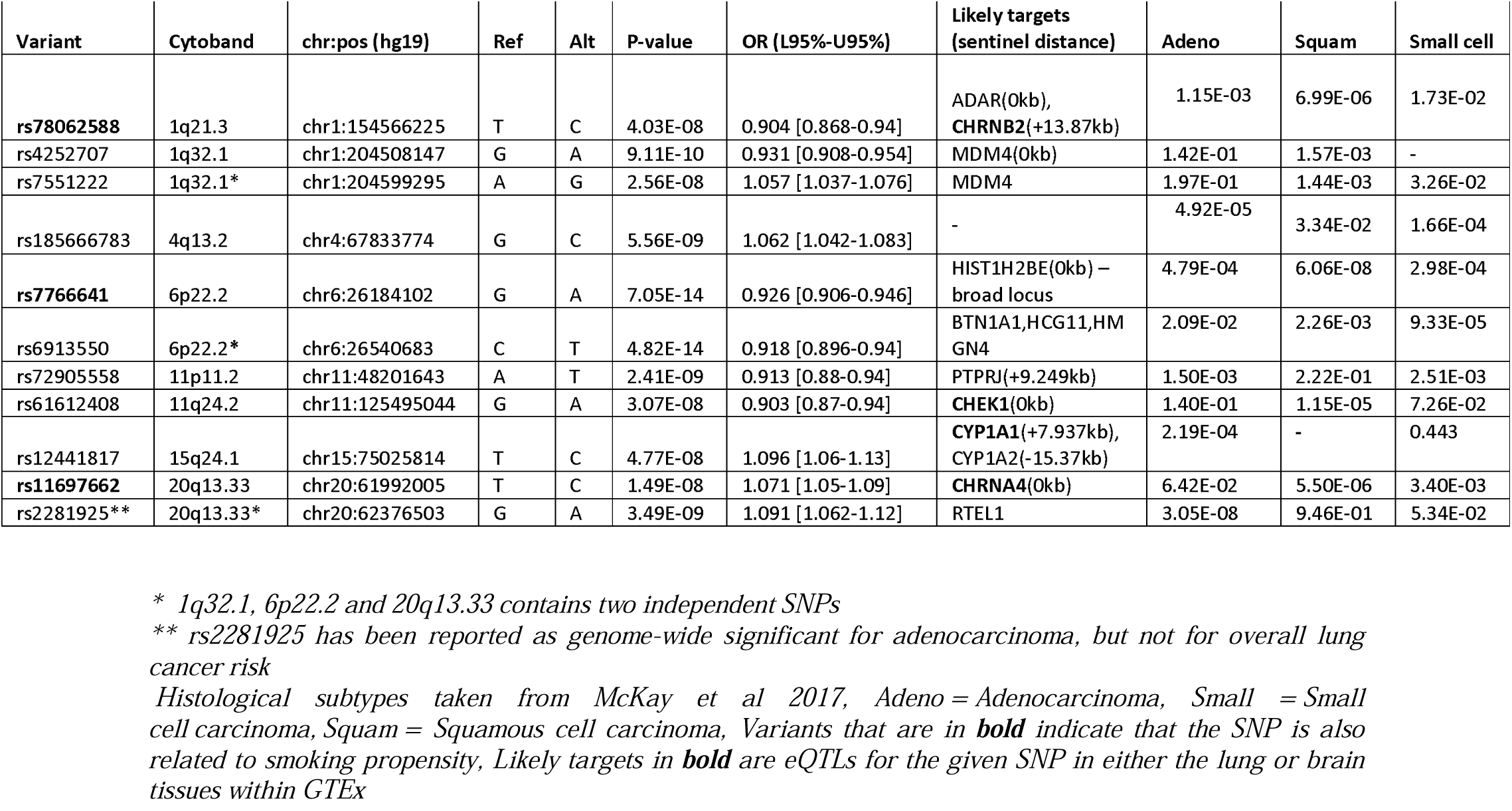
The 8 novel genome-wide significant loci associated with lung cancer risk.

At 1q32.1, 11p11.2, 11q24.2 and 15q24, the sentinel variants (rs4252707, rs72905558, rs61612408, rs12441817, respectively) were not associated with propensity to smoke (Supplementary Table 2). 11q24.2-rs61612408 was associated with the expression of the *CHEK1* gene in multiple tissues including lung epithelia (colocalisation between *CHEK1* lung eQTL and LC: PP4 = 91.1%), with the allele associated with increased expression correlating with decreased risk of lung cancer (Figure 2.C). The association with 11q24.2-rs61612408 appeared to be more prominent in lung squamous cell carcinomas (Table 1). 15q24-rs12441817 is located near the *CYP1A1* and *CYP1A2* enzymatic genes. This locus has been associated with coffee consumption and forced vital capacity (FVC),^28,29^ although there was colocalisation between variants associated with lung cancer only for FVC (colocalisation between coffee consumption and LC: PP4 = 0.0003%, colocalisation between FVC and LC: PP4 = 97.05%) (Supplementary Figure 3). There was evidence that rs12441817 influenced *CYP1A1* expression in the nucleus accumbens (colocalisation PP4 = 70.26%) (Supplementary Figure 4) and an eQTL effect with the processed pseudogene *RP11-10O17*.*1* in lung tissue (colocalisation between eQTL *RP11-10O17*.*1* and LC PP = 95.25%) (Figure 2.D). At 4q13.2-rs185666783 the candidate genes remain ambiguous (AC104806.2 and RNU6-699P) and the association with lung cancer appeared most prominent in lung adenocarcinoma. At 11p11.2, rs72905558 was associated with expression of *C1QTNF4* in lung tissue reported in GTEx but there was no evidence for colocalization between variants related to *C1QTNF4* expression and lung cancer (*C1QTNF4* PP4 = 0.06%).

**Figure 2:**
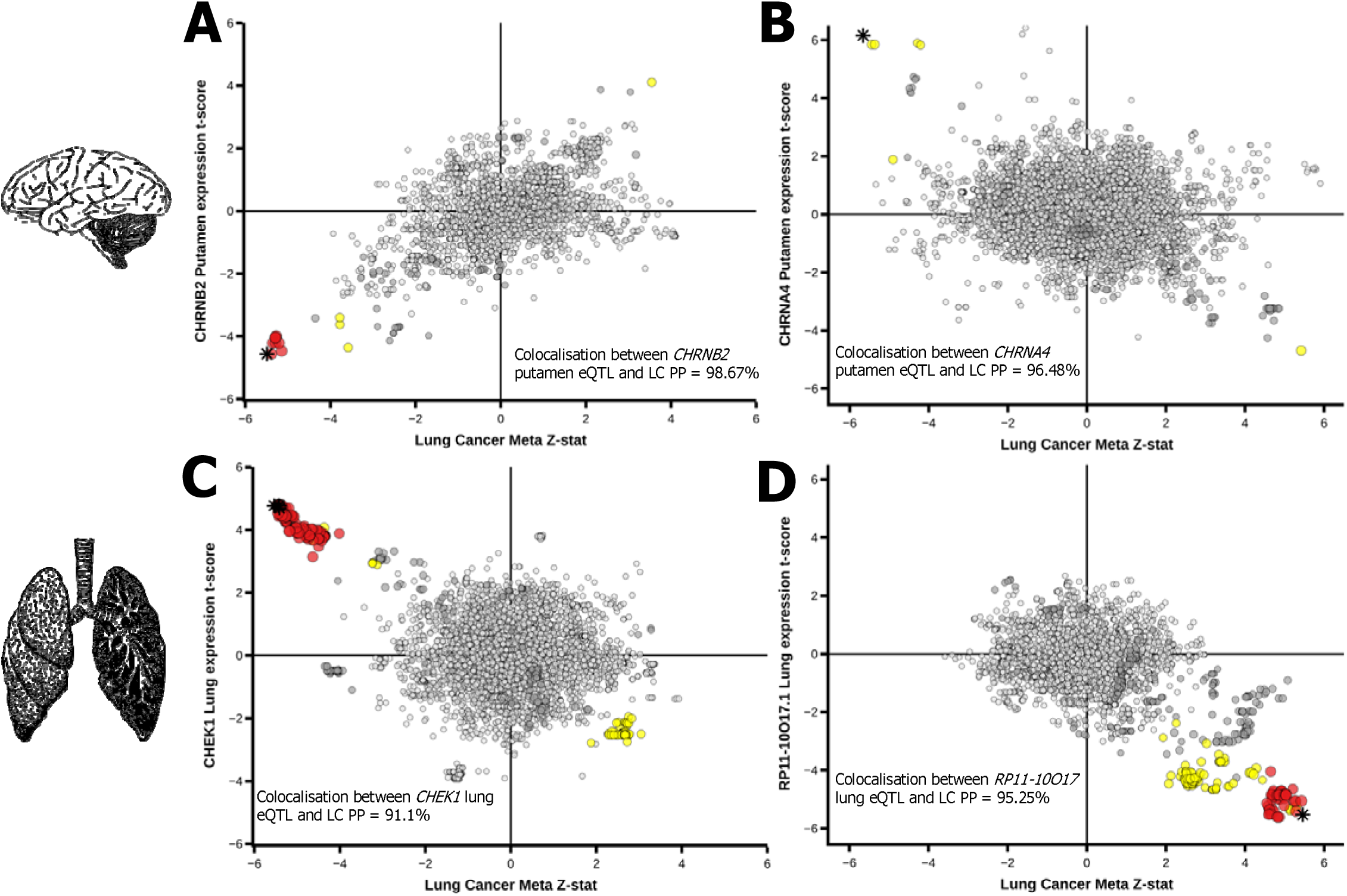
Brain and lung eQTLs discovered within the eight novel loci. Co-localisation between lung cancer (x axis) and *CHRNB2* putamen expression (**A**), *CHRNA4* putamen expression (**B**), *CHEK1* lung expression (**C**) and *RP11-10O17*.*1* lung gene expression (**D**) (y axis). Each variant and eQTL status were compared using COLOC for colocalisation to confirm that the lung cancer SNP was the same SNP driving the eQTL effect in both brain and lung tissues, the Bayesian posterior probability (PP4) of each gene was tested, *CHRNB2* (PP4=98.67%), *CHNRA4* (96.48%), *CHEK1* (91.1%) and *RP11-10O17* (95.25%)

### Exploration of subgenome-wide significant variants and integrative multi-trait polygenic risk score construction

The variants that achieved GW significance also tended to be associated with propensity to smoke and/or an eQTL (Supplementary Table 2). We therefore used partial least squares regression (PLS) to identify subgenome wide significant genetic variants with similar propensity to smoke and/or an eQTL features (represented by PLS components) (see Supplementary Material for details). We constructed bins of variants ranked by these PLS components and represented them as a function of the mean LC association statistic calculated within each bin (Figure 3.A). The bins that were ranked highly by a smoking and/or an eQTL components were observed to have elevated mean LC association statistics relative to most other bins, implying that the variants within these bins are enriched for LC susceptibility alleles (Figure 3.A). Interestingly, this enrichment was more marked for the eQTL PLS component (Figure 3.A). We constructed two polygenic risk scores, smPRS and eQTLPRS, based on the top 100 and 1,000 ranking SNPs from the smoking and eQTLs PLS analyses, respectively (see Methods section and Supplementary Material), with number of variants guided by the degree of enrichment observed (Figure 3.A) and tested them in an independent cohort of 1,666 lung cancer cases and 6,664 matched controls from the UK Biobank. The PRS were robustly associated with lung cancer in this independent series (smPRS: OR per standard deviation = 1.246, 95% CI: 1.176-1.32, P = 8.5 x10^−14^; eQTLPRS: OR = 1.349, 95% CI: 1.27-1.43, P = 7.29 × 10^−17^, combined (both smPRS and eQTLPRS combined after adjusting for variant overlap), OR = 1.366, 95% CI 1.288-1.448, P = 1.44 x10^−25^). These risk estimates were only modestly attenuated when excluding the GWAS significant variants and adjusting for smoking status, again implying that these variants are enriched for LC-susceptibility alleles (Figure 3.B).

**Figure 3:**
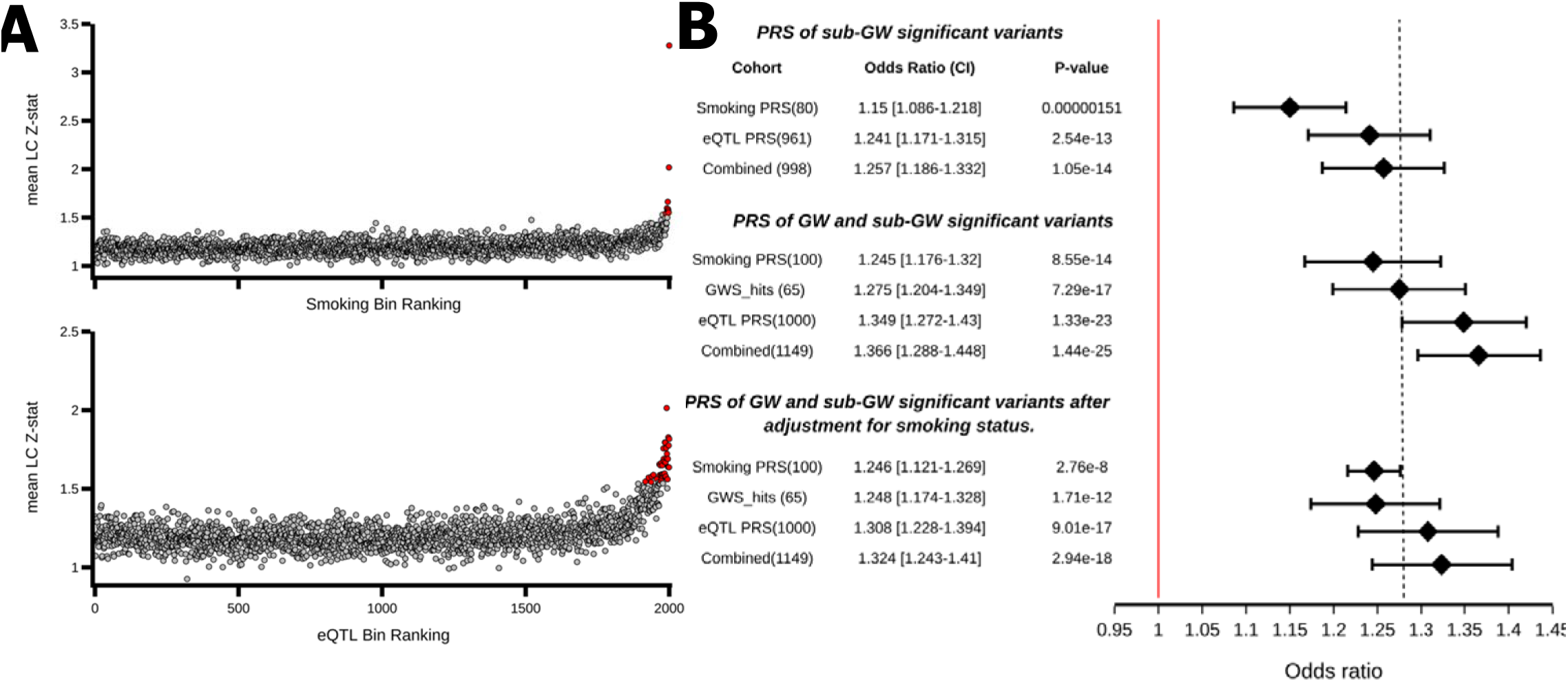
Germline polygenic risk score construction using smoking and eQTL related SNPs and performance testing within the UK Biobank lung cancer cohort. (**A**) The mean lung cancer association statistics calculated by variant bins (100 variants per bin) ranked by component. Variants (clumped on LD based on lung cancer P values) were ranked based on PLS component for smoking propensity (Component1_smoking, top), and eQTLs (Component1_eQTL, bottom) (x axis) and plotted against the mean lung cancer Z statistics calculated across variants in each bin (y axis). Values that exceed 3 SDs from the mean are noted in red (NbinsSmoking =9, NbinsQTL = 37) and are those that have the highest values of the PLS component. (**B**) A Forest plot of the performance for the constructed PRS in comparison to just using the 65 genome-wide significant (GWS) independent loci as a baseline using the model LC ∼ PRS + array + sex + array of recruitment + first 5 PCs. The top panel contains the smoking PRS and the eQTL PRS list without containing any of the 65 GW loci within each list. The middle panel contains the model with smoking status (previous, current, never) added. The bottom panel contains the full lists without adjusting for smoking status. The combined PRS contains all the 65 loci plus both the smoking and eQTL lists.

### PRS germline influences on mutational burden and mutational signatures

We evaluated the association of the smPRS and eQTLPRS with somatic mutational burden in the 736 TCGA lung cancer patients where somatic and germline data overlapped and passed QC metrics (see Methods). There was little evidence for association involving the eQTLPRS and mutation burden (Supplementary Figure 8), however the smPRS was associated with tumour mutational burden (TMB) (P = 1.23×10^−3^, Figure 4.A), with evidence of a trend between increasing polygenic load and somatic mutation burden (Figure 4.A). The smPRS was similarly associated with burden of mutational signatures attributed to tobacco smoke (SBS4 (P = 9.73 × 10^−5^), ID3A (P = 1.78 × 10^−3^), ID3B (P = 3.77 × 10^−2^) and DBS2 (P = 3.05 × 10^−3^)) (Figure 4.A and Supplementary Figure 6). These associations were observed more prominently in patients with LUAD (Figure 4.A). The 15q25 *CHRNA5* lung cancer sentinel variant, rs72740955, had the most striking effect (Supplementary Table 2 and Supplementary Figure 7) but the associations remained significant after excluding genome-wide variants for lung cancer (Figure 4.A). The associations between the smPRS and somatic mutation burden (P = 0.034) and with mutation signatures attributed to tobacco smoking (SBS4: P = 0.023, ID3: p = 0.054, DBS2: P = 0.035, Supplementary Figure 8) were similarly observed in an independent cohort of 61 lung cancer patients whose germline and matched tumour samples have undergone WGS, replicating this finding. We additionally projected the smPRS into other cancer types in TCGA cohorts and the association with TMB was also observed in the esophageal carcinoma (ESCA) cohort (Supplementary Table 6).

**Figure 4:**
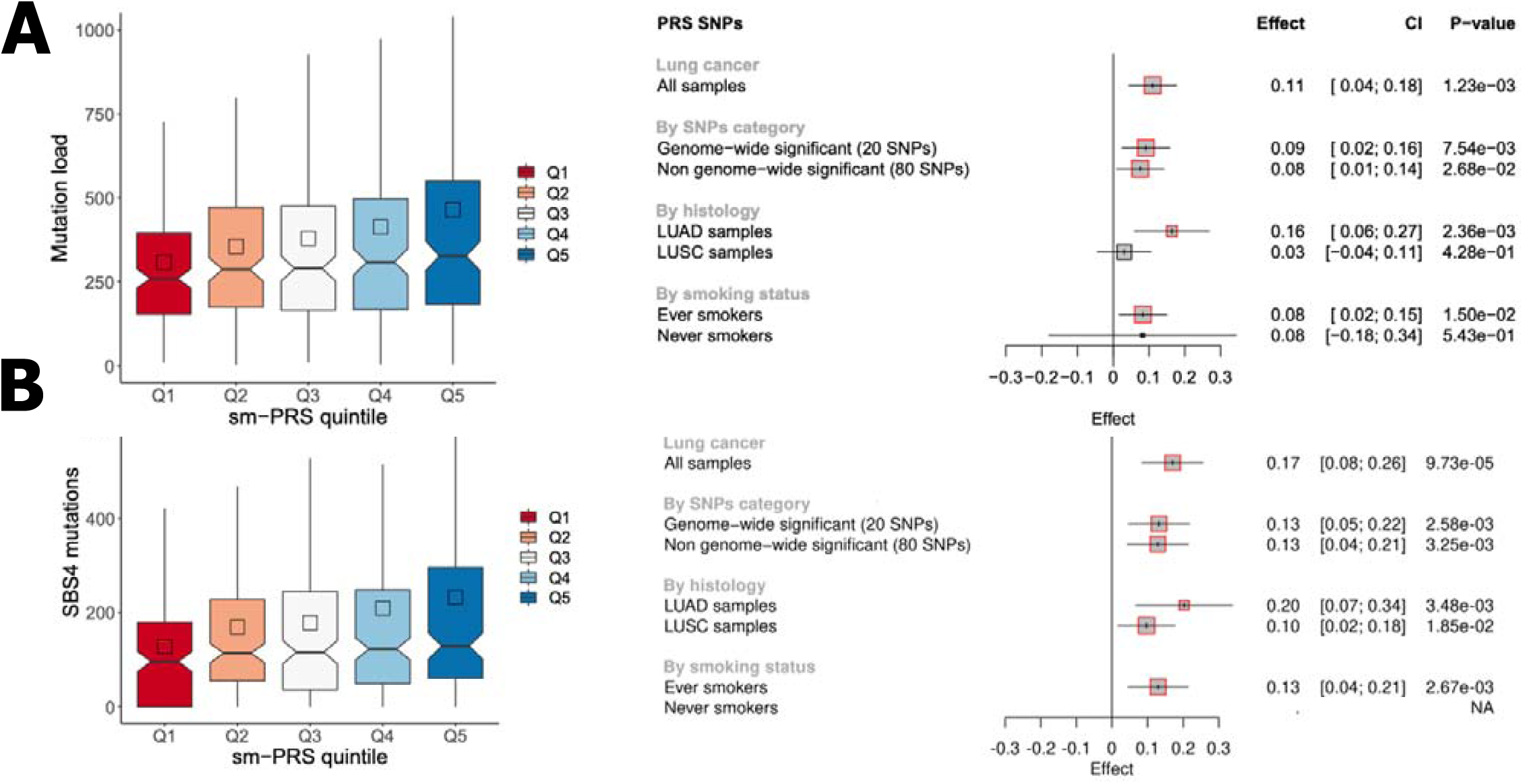
Polygenic risk scores for smoking (smPRS) associations with total number of mutations and mutations attributable to SBS4 in the TCGA cohort. (**A**) Associations with total number of mutations. (**B**) Associations with SBS4 mutations. The left panels represent the distribution of the number of mutations in the sm-PRS quintiles. The right panels correspond, respectively, to the forest plots of sm-PRS associations with total mutational burden (panel A) and SBS4 mutations (panel B). For each PRS, the association was tested: i) in all lung cancer cases when considering all SNPs in the smPRS SNPs selection, in all lung cancer cases when considering different subsets of SNPs in the PRS computation, iii) stratifying by histology, iv) stratifying by smoking status. Gray squares correspond to the estimate resulting from Quasi-Poisson models. The squares are highlighted in red when the associated P-value is below 0.05.

## DISCUSSION

This study identified 21 lung cancer susceptibility loci, including eight novel loci by combining large, genotyped biobank data and traditional genome-wide association studies. Three of eight novel loci were also associated with propensity to smoke. This included brain eQTLs variants for both subunits of the neuronal nAChRs α4β2 receptor. Variants in LD with the α4 subunit (rs2373500) have been described in nicotine dependency and lung cancer risk, albeit not at GW significance for lung cancer,^18^ while the β2 receptor and lung cancer risk has not been described. The neuronal nAChRs α4β2 receptor is the most abundant nAChR subtype within the human brain and important within the dopaminergic signalling pathway. The α4β2 receptor has a key role in nicotine dependence behaviours^30^ and a major target in nicotine addiction intervention.^31,32^ The third novel locus related to lung cancer and propensity to smoke is telomeric to the MHC region, where the target candidate gene(s) is less obvious. The MHC region was among the first susceptibility loci to be associated with lung cancer.^6,33–35^ However, rs7766641, is not in LD with these previously described variants (R^2^< 0.001) and associated with the number of cigarettes smoked per day, implying that these are distinct associations.

This meta-analysis also identified additional lung cancer susceptibility loci that appear to be independent of smoking propensity. This included variants at 15q24 near *CYP1A1, CYP1A2* and *CYP11A1* that participate in the metabolism of many different xenobiotics and some endogenous substrates. Variants at the 15q24 *CYP1A1* / *CYP1A2* locus have been linked with multiple traits, notably other forms of propensity (coffee consumption) and forced vital capacity (FVC).^28,29^ The coffee consumption/FVC variants at this locus appear distinct and colocalisation analysis implicates FVC more in the lung cancer association, which also seems aetiologically more plausible. For tissue expression, rs12441817 colocalised with lung tissue expression of the processed pseudogene *RP11-10O17*.*1* (Figure 2.D) although how this pseudogene relates to lung cancer susceptibility is unclear.

An additional novel lung cancer susceptibility variant, rs61612408, was a lung tissue eQTL for the DNA repair gene *CHEK1* (Figure 2.C). Similar to previously described variants associated near *CHEK2* and *BRCA2*, the association between rs61612408 and lung cancer appears more prominent in lung squamous cell carcinomas^9,10^. We additionally noted the variant impacting the *MDM4* gene, which is an important p53 regulator. This variant was previously associated with non-glioblastoma tumours^36^ and more recently squamous cell carcinomas of the lung and head / neck,^37^ although here we noted weak evidence for association in lung adenocarcinoma (Table 1). At 11p11.2 colocalisation analysis showed little evidence for involvement with genes *C1QTNF4* (lung) and *MTCH2* (brain-cortex), suggesting that these signals are unlikely to explain the lung cancer association, one other candidate is potentially *PTPRO* which is hypermethylated in several cancers including lung.^38,39^ At 4q13.2, the finding remains ambiguous, but from histological subtypes analysis performed from the previous reported GWAS study, it appears that this signal is mostly found in lung adenocarcinoma.

We additionally sought to use the shared genetic aetiology between lung cancer susceptibility and smoking related traits and gene expression annotations (eQTL) to explore variants that did not achieve GW significance. We used the partial least squared (PLS) method to select variants related to these traits for the PRS analysis and demonstrated that such variants are indeed enriched for susceptibility alleles. While the role of these individual variants remains to be confirmed, these sub-GW significant variants were located near relevant candidate genes (smoking traits like *CHNRA6, DBH* and eQTLs for *ERCC2, RAD51C, XRCC3* and *CASP8*). Combining both sub-GW PRS lists (smoking PRS and eQTL PRS) with GW significant results reached an OR of 1.36 per standard deviation unit increase in score improving on previous PRS predictions (OR 1.17 and 1.26),^40,41^ despite the conservative clumping approach (R2 < 0.1) employed. This suggests that integrating functional annotations may be of interest for PRS.

Lastly, the analysis of the smoking PRS demonstrated an association between a person’s genetic risk load and mutation burden, and or burden of tobacco-related somatic mutational signatures, within two independent case cohorts and using different sequencing methods (exome sequencing and whole genome sequencing). These associations appear consistent with the notion that genetic variants influence an individual’s smoking behaviour, which in turn, influences their carcinogenic exposure and consequently, their somatic mutation burden.

In conclusion, this work has increased the number of variants associated with lung cancer susceptibility, with the identification of novel susceptibility loci. PRS analysis highlighted that many additional variants remain to be discovered and provided insights into the carcinogenic mechanisms.

## Data Availability

Data is available on request - PRS panels and weights are within the supplementary data

## Acknowledgements

We would like to acknowledge the TCGA Research Network (https://www.cancer.gov/tcga) and the contribution of specimen donors and research groups involved in this resource. We also would like to acknowledge the GTEx project and the supporting bodies (https://commonfund.nih.gov/GTEx), specimen donors and research groups. Additionally, we would like to acknowledge the work carried about by the Benjamin Neale lab for their work on the UK Biobank (http://www.nealelab.is/uk-biobank/).

The ILLCO consortium is listed in the supplementary text with affiliations.

## Disclaimer

Where authors are identified as personnel of the International Agency for Research on Cancer/World Health Organization, the authors alone are responsible for the views expressed in this article and they do not necessarily represent the decisions, policy, or views of the International Agency for Research on Cancer/World Health Organization.

## Lead contact

Further information and requests for resources and reagents should be directed to and will be fulfilled by the Lead Contact, James McKay

## Data and Code Availability

The code generated during this study are available at GitHub (https://github.com/IARC-genetics/GWAx_lung_cancer). The polygenic risk scores variants used in this study are available within the supplementary tables. Summary statistics from the UK Biobank lung cancer family history summary statistics will be made available on GWAS Catalog. Summary statistics from the meta-analysis (McKay et al. 2017 and the UK Biobank lung cancer family history) are not publicly available due to controlled access of Oncoarray consortium data. Oncoarray data can be accessed by the database of Genotypes and Phenotypes (dbGaP) under accession phs000876.v1.p1

## CRediT author Statement

**Aurélie AG Gabriel:** Conceptualization, Formal analysis, Data curation, Methodology, Investigation, Software, Validation, Writing – Draft, review and editing, Visualization **Joshua R Atkins**: Conceptualization, Formal analysis, Data curation, Methodology, Investigation, Software, Validation, Writing – Draft, review and editing, Visualization **Ricardo CC Penha**: Formal analysis, Data curation, Validation, Investigation, Writing-Review & Editing. **Karl Smith-Byrne**: Formal analysis, Writing-Review & Editing **Valerie Gaborieau**: Formal analysis, Data curation **Catherine Voegele**: Formal analysis, Data curation **Behnoush Abedi-Ardekani**: Formal analysis, Data curation **Maja Milojevic**: Formal analysis, Data curation **Robert Olaso**: Data curation **Vincent Meyer**: Data curation **Anne Boland**: Data curation **Jean François**: Data curation **David Zaridze**: Resources **Anush Mukeriya**: Resources **Beata Swiatkowska**: Resources **Vladimir Janout**: Resources **Miriam Schejbalová**: Resources **Dana Mates**: Resources **Jelena Stojši**c: Resources **Miodrag Ognjanovic**: Resources **the ILCCO consortium**: Resources, Funding acquisition **John S Witte**: Data curation **Sara R Rashkin**: Data curation **Linda Kachuri**: Writing - Review and Editing **Rayjean J Hung**: Writing - Review and Editing **Siddhartha Kar**: Writing - Review and Editing **Paul Brennan**: Resources **Anne-Sophie Sertier**: Data curation **Anthony Ferrari**: Data curation **Alain Viari**: Data curation **Mattias Johansson**: Writing - Review and Editing **Christopher I Amos**: Conceptualization, Writing - Review and Editing, Funding acquisition, Resources **Matthieu Foll**: Conceptualization, Supervision, Methodology, Writing – Draft, review and editing **James D McKay**: Conceptualization, Supervision, Data curation, Methodology, Writing – Draft, review and editing, Funding acquisition

## Supplementary Figures

Supplementary Figure 1: Genomic inflation and quantile–quantile plot across studies that were meta-analysed

Supplementary Figure 2: Visual validation of genome-wide by proxy method by Manhattan plot compared to the lung cancer genome-wide association study.

Supplementary Figure 3: Z-statistic plots for variants associated with traits at 15q24(*CYP1A1*) compared to lung cancer

Supplementary Figure 4: *CYPA1A* expression in the nucleus accumbens

Supplementary Figure 5: Partial least squares of mean z-scores for lung cancer for the polygenic risk scores construction and correlation across smoking traits and eQTLs

Supplementary Figure 6: The smPRS and eQTLPRS associations with mutational signatures related to smoking attributed to tobacco

Supplementary Figure 7: Mutational burden in lung tumours across rs72740955 genotype categories

Supplementary Figure 8: eQTLPRS associations with total number of mutations and mutations attributable to SBS4 in the TCGA cohort

Supplementary Figure 9: Replication analysis for the association of PRS with somatic mutational load in the GeniLuc cohort

## Supplementary Tables within Supplementary Materials

Supplementary Table 1. UK Biobank Sample selection and filtering.

Supplementary Table 2. 65 Genome wide significance variants identified by the GWAx-GWAS meta-analysis.

Supplementary Table 3. eQTL analysis on rs78062588 and rs11697662.

Supplementary Table 4. PRS panels for smPRS and eQTLPRS.

Supplementary Table 5. Association between PRS and lung cancer in the TCGA case cohorts. Association in lung cancer versus all other cancers.

Supplementary Table 6. Association of tobacco-smoking PRS (sm-PRS) with mutation load and SBS4 by TCGA cohort.

