## Supplementary Material for "Genetic analysis of lung cancer reveals novel susceptibility loci and germline impact on somatic mutation burden"

**Page number of sections**

Supplementary Methods – Page 1

Supplementary References – Page 6

Supplementary Tables – Page 7

Supplementary Figures – Page 55

**Supplementary Methods**

**Datasets:**

***Transdisciplinary Research for Cancer in Lung TRICL***

The TRICL cohort is part of the International Lung Cancer Consortium (ILCCO), an international group of lung cancer researchers (more details can be found on the website <https://ilcco.iarc.fr/>). This dataset has been explained in detail in (McKay et al., 2017), with the breakdown of the study populations provided in the supplementary information of that paper. Genotyping was performed on the OncoArray which features additional probes for fine mapping of common cancer susceptibility loci. Imputation was performed using the 1000 Genomes Project Phase 3 panel limiting our meta-analysis discovery to this strata including directly genotyped probes found on the OncoArray. Summary statistics generated by this meta-analysis was utilised in this current paper.

***UK Biobank***

The UK Biobank contains over 500,000 individuals making this resource one of the largest with a goal of enabling new discoveries to improve public health (<https://www.ukbiobank.ac.uk/>). This contains both genetic data conducted on the UK BiLEVE array and the UK Biobank Axiom array. Outside the genetics data, the UK Biobank is rich in phenotypic health data containing information extracted from NHS registries and medical records. Furthermore, individuals recruited to the program were invited for in-person consultation at one of the study research centres were an array of information, biological and imaging assessment were undertaken. All individuals provided informed constant and the UK biobank has ethical approval from the National Research Ethics Service Committee North West-Haydock. More detailed information can be found on the UK Biobank website or by the paper (Sudlow et al., 2015).

***The Cancer Genome Atlas projects (TCGA)***

TCGA is a joint effort in America between the National Cancer Institute and the National Human Genome Research Institute that has characterised over 10,000 primary cancers across 33 cancer types. TCGA access was obtained through TCGA project 2731 via dbGAP. Genotyping was performed for the samples from the 33 TCGA cohorts on the raw intensities CEL files downloaded on the GDC Legacy Archive portal using gdc-client (v1.4.0). Only blood and/or normal tissue samples with DNA analyte were downloaded (in total 11837 files). The CEL files were then filtered out if associated with one of the following TCGA non-rescinded annotations: "Item flagged DNU", "Administrative Compliance", "Item does/may not meet study protocol", "Qualified in error", "BCR Notification", "Normal tissue origin incorrect", "Subject withdrew consent", "Normal class but appears diseased", "Duplicate item", "Tumor tissue origin incorrect", "Tumor type incorrect", "Genotype mismatch", "Permanently missing item or object" and "Qualification metrics changed". Also, when multiple samples were available for a participant, blood samples were selected when available.

10,443 samples remained after filtering on annotations and removing duplicated samples and were used for the genotyping. Prior to genotyping, a quality control function (apt-geno-qc) from Affymetrix Power Tools (APT 2.10.2.2) was applied to the dataset and samples with a contrast QC above 0.4 were selected for genotyping. The largest TCGA cohort, BRCA, was genotyped to define a list of probes with appropriate performance (for example, probe-level genotyping call rates of at least 97%). This probe list was then used with the apt-probeset-genotype program and Birdseed (v2) algorithm to genotype the TCGA samples. Samples with low genotyping call rate of < 97% were removed and a second round of genotyping was performed on each cohort. Sex check (using *plink*) was performed and compared to the clinical data. Inter sample's relatedness (using *plink genome* with min parameter fixed at 0.185) was also tested. Among the 9,855 TCGA samples with reported sex, 46 samples had discordance between reported and imputed sex and 17 pairs of relatives were identified. These samples were not considered in the PRS analyses. Heterozygosity and genotyping missing rates *(--het* and *--missing* options respectively) were also checked in order to select samples with a genotyping missing rate lower than 3% and to ensure that the heterozygosity rate was homogeneous across samples.

Population stratification was performed using admixture (version 1.3) (Alexander et al., 2009) based on HapMap2 data. SNPs were defined by Yu et al., 2008. Among 12,898 SNPs, 11,630 were in common between the HapMap and the TCGA datasets. Around 99% of the samples reported as “WHITE” by the TCGA clinical data were predicted Europeans. Eigenstrat (without outlier removal) was used on the European samples to account for further population stratification; the first 5 principal components (PCs) were kept for downstream analyses.

Considering all TCGA samples, SNPs with genotyping call rate and MAF lower than 97% and 1% respectively were removed. In each ancestry group, we applied the *HRC-1000G-check-bim.pl* script from the McCarthy tools (<https://www.well.ox.ac.uk/%7Ewrayner/tools/#Checking>), removing SNPs with unmatched positions and/or alleles, duplicated SNPs, as well as SNPs with allele frequencies (AF) differing from reported AF in the same population in the 1000 Genome dataset (more than 20% difference). SNPs with a genotyping call rate below 97% and showing strong deviation (P-value < 10^-8^) from the Hardy Weinberg equilibrium (*hwe* plink option) in any of the ancestry groups were excluded. Finally, ambiguous SNPs were excluded prior to imputation. Phasing and imputation were performed on each chromosome by 20Mb chunks. Phasing was performed using Eagle (v2.4.1, flanking regions of 5Mb) (Das et al., 2016; Loh et al., 2016) with the 1000 Genomes phase 3 reference panel. Minimac4 (v1.0.1) (Das et al., 2016) was used to perform the imputation with a window size of 500kb.

***GeniLuc cohort and whole genome sequencing.***

Lung cancer cases and controls were recruited through a multicentric case-control study coordinated by the International Agency for Research on Cancer in Russia, Poland, Serbia, Czech Republic, and Romania from 2005 to 2013. Cases were incident cancer patients collected from general hospitals. Controls were recruited from individuals visiting general hospitals and out-patient clinics for disorders unrelated to lung cancer and/or its associated risk factors, or from the general population. Information on lifestyle risk factors, medical and family history was collected from subjects by interview using a standard questionnaire. All study participants provided written informed consent. Recruitment included collection of normal material in the form of a blood sample and a resection of the patient’s tumour. Subsequent to histopathological review to ensure appropriate tumour purity, DNA was extracted from normal material (blood) and the tumour resection. Whole genome sequencing was undertaken using PCR free whole genome library preparation and DNA sequencing undertaken to a depth of 30X for the paired tumour normal for each patient using an Illumina X five DNA sequencer at the CEA laboratory in Paris France. Ethics for the GeniLuc cohort was reviewed by the IARC ethnics board (IARC IRB 12-05) and approved on 28th of April 2016.

***Quality control performed on the UK Biobank dataset***

Quality control (QC) was based on the support document provided by the UK Biobank and which resulted in similar QC metrics via inhouse practises. Supplementary Table 1 contains the measures performed before association testing was conducted with the numbers of samples that were removed at each step. We also provide the UK Biobank fields for each QC measure performed.

***Processing of raw compressed UK Biobank information into functional data and association testing***

UK Biobank BGEN chunked files were merged using cat-BGEN(Band and Marchini, 2018) into full chromosomes and converted into the PLINK 2.0 PGEN format. This allowed for quick access and processing suited to our high-performance computing cluster. Association testing was performed in plink 2.0 using the –glm function with the hide-covar parameter using 64 GBs of memory and 8 cores for each chromosome. The covariate file contained 14 fields including the sex, array type, age of recruitment, the first 5 PCs taken from the UK Biobank and the total number of siblings. Summary statistics were then concatenated back into a whole dataset.

***Genome-wide association by proxy (GWAx) of lung cancer and validation***

After exclusions based on quality control, we identified 42,124 and 6,719 individuals that reported a parent or a sibling with lung cancer, respectively, as well as 197,029 individuals without a clinical cancer diagnosis who did not report a family history of cancer from the UK Biobank. We undertook a family history GWAS (GWAx) of 48,843 self-reported “family history lung cancer cases” and 197,029 “controls”.

There was limited evidence for any inflation of GWAS test statistics derived from this approach (adjusted genomic control inflation factor (*λ*_GC_) lambda: 1.001; LD score regression (LDSC) intercept: 1.009 (SE=0.0068)) (Supplementary Figure 1). The GWAx was able to five loci (5p15.33, 6p21.32, 12p13.33, 13q13.1 and 15q25.1) that had previously been discovered from the traditional GWAS (Supplementary Table 2 and Supplementary Figure 3). To test whether the GWAx study shared the same genetic overlap with the lung cancer GWAS, we used LDSC based genetic correlation and demonstrated that results from these approaches were strongly genetically correlated (r_g_ =1, se=0.066, p= 4.00 x 10^-52^). Furthermore, as lung cancer shares genetic architecture with smoking propensity traits (Jiang et al., 2019), we obtained summary statistics for age of first cigarette, cigarettes per a day, smoking cessation and smoking initiation from (Liu et al., 2019) to examine the genetic correlation with LC risk using both a previously conducted GWAS of 29,266 LC cases and 56,450 controls (McKay et al., 2017) and our GWAx approach. LDSC indicated that for each comparison there was little difference between the GWAx method and the GWAS approach.

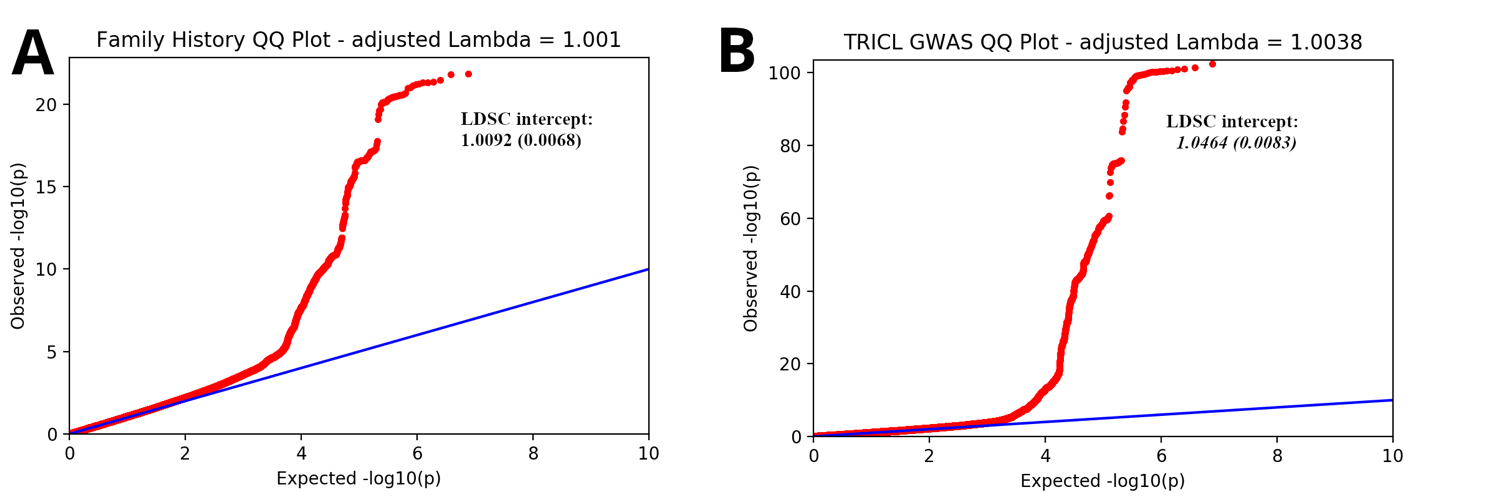

***Supplementary Figure 1: Genomic inflation and quantile–quantile plot across studies that were meta-analysed.*** The family history GWAx study (**A**) and the TRICL GWAS (**B**) both had minimal genomic inflation recorded due to population structure or cryptic relatedness and for these reasons they were meta-analysed. The genomic control inflation factor (*λ*_GC_) was calculated by the median observed $\chi^{2}$test statistic divided by the expected using the percent point function. In python, this is performed by the scipy libraries to convert the p-values distribution into $\chi^{2}$ distribution (scipy.stats.ppf(1-pvalues,1)) then taking the median value and dividing this to the median of the $\chi^{2}$distribution with 1 degree freedom (np.median(chisq)/sci.chi2.ppf(0.5,1)).(Tsepilov et al., 2013) This observed *λ*_GC_ was adjusted to a target sample size of 1,000 cases to 1,000 controls using the formula below.

Adjusted lambda = 1 + $\frac{\left( lambda_{obs}-1 \right)*\left( \frac{1}{n_{cases}} \right)+\left( \frac{1}{n_{controls}} \right)}{\left( \frac{1}{n_{targetcases}} \right)+ \left( \frac{1}{n_{targetcontrols}} \right)}$

Where, n_cases_ = numbers of cases in GWAS, n_controls_ = numbers of controls in GWAS, n_targetcases_ and n_targetcontrols_ are both the target samples size which in this case is 1,000 for both. Due to the nature of the sample size of both datasets and the strength of association of many variants particularly on chr15 and chr6, we also reported the LDSC intercept, with this value being close to 1, we accept that this inflation is mostly due to a true polygenic signal.

**Exploration of sub-genome-wide significant variants and integrative multi-trait polygenic risk score construction.**

The polygenic risk score analysis included genome-wide (GW) significant variants but we additionally included relevant SNPs that did not pass the GW significance threshold. The choice of “relevant” sub-GW significant SNPs was guided by the observation that many of the GW significant variants also tend to be associated with smoking-related traits and/or eQTL’s. As such we sought to also include in the polygenic risk scores sub-GW significant variants that had similar characteristics. We therefore obtained summary statistics of previously published GWAS on smoking-related traits. We choose the traits studied by the GWAS and Sequencing Consortium of Alcohol and Nicotine use (GSCAN) consortium in a large dataset gathering up to 1.2 million samples (Liu et al., 2019). This study explored four smoking traits (cigarettes per day, smoking cessation, smoking initiation and age of initiation) as well as drinking consumption (drinks per week). It has also been shown by Jiang et al. that there is a shared heritability, driven mostly by shared aetiology of tobacco smoking between lung cancer and head and neck cancer (Jiang et al., 2019). As such, the summary statistics of the GWAS on head and neck cancer performed by Lesseur et al. was also considered (Lesseur et al., 2016). Secondly, we gathered eQTLs summary statistics collected across 49 different tissues from GTEx data (version 8) for 7,562,169 variants from the GWAS-GWAx meta-analysis. The eQTL T-values (the top value for each gene) for each tissue were retained for each variant.

We also noted that variants associated with smoking propensity tend to be associated with more than one of these traits, for example a variant associated with cigarettes per day may also be associated with smoking cessation etc. Similarly, many eQTLs are significantly associated with expression levels across multiple tissues (Supplementary Table 2, Supplementary Figure 6). We therefore used a partial least square (PLS) (plsr function from the pls Rpackage) based approach to select variants that tended to have similar smoking and eQTL features, respectively. The PLS model was used considering the Z-statistics from the lung cancer GWAS-GWAx meta-analysis SNPs as the response variable and the association statistics for the smoking propensity traits (smoking PLS) or eQTLs (eQTL PLS) as the explanatory variables. The smoking PLS and eQTL PLS were undertaken separately. The first component of the smoking and eQTL PLS explained 1.13% and 0.53% of the variance in LC association statistics, respectively. The first component of the smoking and eQTL components tended to be correlated with being associated with smoking propensity traits (Component 1_smoking) or eQTL across multiple tissues, including lung epithelia (Component1_eQTLs) (Supplementary Figure 6).

Next, we sought to explore the nature of the correlation between the components defined by the PLS and the lung cancer association statistics. Firstly, we clumped the 7,500,000 SNPs based on LD (R^2^ < 0.1 and 10,000 kb) which resulted in approximately 230,000 independent variants. We then ranked these variants by the value of the PLS component from highest to lowest. Based on this ranking, we then created bins of 100 variants (approximately 2,300 bins). We then represented these as a function of the mean Z statistic for lung cancer within each bin and their order in the PLS component (Figure 3a). The bins that were highly ranked, i.e. having the highest PLS component values, tended also to have higher mean lung cancer association statistics relative to other bins, implying that these variants are enriched for lung cancer susceptibility variants. We observed that the degree of enrichment for lung cancer related SNPs varied between the smoking and eQTL PLS results (Figure 3a).

**Polygenic risk score development.**

Finally, we selected variants for polygenic risk score analysis to be applied in two independent lung cancer data sets (see methods). We first selected variants (clumped as per above) that were in the top 1% variants associated with lung cancer (i.e. above the 99th percentile of LC Z-statistic). We then ranked these variants based on the smoking and eQTLs PLS components and selected the top 100 and top 1,000 variants for smoking and eQTL respectively. We retained a higher number of variants from the eQTLPLS due to the more pronounced enrichment of lung cancer susceptibility for bins of these SNPs (Figure 3a). Based on the smoking PLS and eQTL PLS SNPs selections, two PRS were constructed: smPRS and eQTLPRS. The PRS were weighted using association statistics from the lung cancer GWAx meta-analysis (see equation below) and projected into independent lung cancer cohorts.

$$\sum_{i=1}^{n} \beta*Xd$$

*with Xd defined as the individual’s SNP dosage, and β defined as the effect from the lung cancer meta-analysis, n = 100 SNPs for the smPRS and n = 1000 SNPs for the eQTLPRS.*

**The ILLCO consortium**

Albanes D^1^, Aldrich M.C^2^, Andrew A^3^, Arnold Sm^4^ , Bickeböller H^5^, Bojesen S.E^6,7^, Brunnström H^8^, Caporaso N^1^, Chen C^9^ , Christiani D^10^, Teare M.D^11^, Field JK^12^ , Grankvist K^13^, Hong Y^14^, Kiemeney L.A^15^, Lam S^16^, Landi M^1^, Lazarus P^17^, Liu G^18^, Marchand L.L^19^, Melander O^20,21^ , Johansson MB^22^ , Rennert G^23^, Risch A^24,25,26,27^, Schabath M.B^28^, Shen H^29^, Shete S.S^30^, Tardon A^31,32^, Wichmann H.E^33,34,35^, Yaun J.M^36^, Zienolddiny S^37^

Johansson M, Brennan P, Hung R, Amos C, McKay J affiliations are within the main authors of this study

1. National Cancer Institute, United States
2. Department of Thoracic Surgery, Division of Epidemiology, Vanderbilt University Medical Center, Nashville, Tennessee, United States
3. Norris Cotton Cancer Center, New Hampshire, United States
4. Markey Cancer Center, The University of Kentucky, Lexington, Kentucky, United States
5. University Medical Center Goettingen, Göttingen, Germany
6. Herlev and Gentofte Hospital, Copenhagen, Denmark
7. Faculty of Health and Medical Sciences, University of Copenhagen, Copenhagen, Denmark
8. Lund University, Laboratory Medicine Region Skåne, Department of Clinical Sciences Lund, Pathology, Lund, Sweden
9. Fred Hutchinson Cancer Research Center Public Health Sciences, Seattle, Washington, United States
10. Harvard School of Public Health, Boston, Massachusetts, United States
11. University of Sheffield, Sheffield, United Kingdom
12. The University of Liverpool, Liverpool, United Kingdom
13. Department of Medical Biosciences, Umeå University, 90185 Umeå, Sweden
14. Department of Preventive Medicine, Seoul National University College of Medicine, Seoul, South Korea
15. Radboud University Medical Center, Nijmegen, Netherlands
16. British Columbia Cancer Agency, British Columbia, Canada
17. Washington State University College of Pharmacy and Pharmaceutical Sciences, Spokane, Washington, United States
18. Princess Margaret Cancer Center, Toronto, Ontario, Canada
19. Cancer Epidemiology Program, University of Hawaiʻi Cancer Center, Hawaiʻi , United States
20. Department of Clinical Sciences Malmö, Lund University, Sweden
21. Department of Internal Medicine, Skåne University Hospital, Malmö, Sweden
22. Department of Radiation Sciences, Umeå University, 90185 Umeå, Sweden
23. Carmel Medical Center, Haifa, Israel
24. University of Salzburg and Cancer Cluster Salzburg, The University of Salzburg, Salzburg, Austria
25. Translational Lung Research Center, Heidelberg, Germany
26. The German Center for Lung Research, Germany
27. German Cancer Research Center, Heidelberg, Germany
28. Department of Cancer Epidemiology, H. Lee Moffitt Cancer Center and Research Institute, Tampa, Florida, United States
29. Department of Epidemiology and Biostatistics, Jiangsu Key Lab of Cancer Biomarkers, Prevention and Treatment, Collaborative Innovation Center for Cancer Personalized Medicine, School of Public Health, Nanjing Medical University, Nanjing, P.R. China
30. The University of Texas MD Anderson Cancer Center, Houston, Texas
31. The University Institute of Oncology of Asturias – Cajastur Social Programme- IUOPA, The University of Oviedo, Oviedo, Asturias, Spain
32. The Epidemiology and Public Health Networking Biomedical Research Centre, Planta, Madrid, Spain
33. Institute of Medical Informatics, Biometry and Epidemiology, Ludwig Maximilians University, Munich, Germany
34. Institute of Epidemiology, Helmholtz Center Munich, Neuherberg, Oberschleißheim, Germany
35. Institute of Medical Statistics and Epidemiology, Technical University Munich, Munich, Germany
36. University of Pittsburgh Cancer Institute, Pittsburgh, Pennsylvania, United States
37. National Institute of Occupational Health, Oslo, Norway

**Supplementary Tables**

**Supplementary Table 1 -UK Biobank Sample selection and filtering**

| **Exclusion field** | **Number of Samples removed** | **UK Biobank data field** |
| --- | --- | --- |
| Not white British Ancestry | 78422 | 2001 |
| Sex discrepancy | 308 | 22001 |
| Aneuploidy | 407 | 22019 |
| 2nd degree relative | 32674 | 22021 |
| Excluded from the kinship inference procedure | 2 | 22021 |
| Excess relatives | 151 |  |
| **Total samples for analysis count** | **Number of Samples remaining** |  |
| Total parent history count after filtering | 29935 | 20107 |
| Total maternal history count after filtering | 13908 | 20110 |
| Total Sibling history count after filtering | 6719 | 20111 |
| Total number of controls used after filtering | 197029 | !==20107,20110,20111,40006, 2453 |
| **Total samples after in-house QC including large missingness, withdrawn consent etc** |  |  |
| Total FH proxy cases | 48843 |  |
| Total FH proxy controls | 195387 |  |
| UK Biobank Lung cancer cohort after filtering | 1666 | 2453,40006 |
| Independent controls used with lung cancer cohort (UK Biobank) | 6664 | !==20107,20110,20111,40006,2453 |

**Supplementary Table 2 - 65 Genome wide significance variants identified by the GWAx-GWAS meta-analysis.**

| **Variant** | **Cytoband** | **chr:pos(hg19)** | **Ref** | **Alt** | **P-value** | **OR(L95%-U95%)** | **Allele Frequency** | **Likely targets with 50KB** | **Smoking related (P-value) *** | **GTEx Lung eQTL** | **GTEx Brain eQTL** | **Adeno** | **Squam** | Small cell | **LC_overall_P** | **GWAx_P** |
| --- | --- | --- | --- | --- | --- | --- | --- | --- | --- | --- | --- | --- | --- | --- | --- | --- |
| rs71658797 | 1p31.1 | chr1:77967507 | T | A | 1.77E-12 | 1.107 [1.079-1.136] | 0.09 | AK5, FUBP1 | - | FUBP1 | FUBP1 | 3.12E-10 | 7.28E-05 | 1.28E-02 | 3.25E-11 | 0.00171522 |
| rs78062588 | 1q21.3 | chr1:154566225 | T | C | 4.03E-08 | 0.904 [0.868-0.94] | 0.07 | CHRNB2 | CPD (1.65e-07) | - | CHRNB2 | 1.15E-03 | 6.99E-06 | 1.73E-02 | 4.60E-07 | 0.00944037 |
| rs4252707 | 1q32.1 | chr1:204508147 | G | A | 9.11E-10 | 0.931 [0.908-0.954] | 0.22 | MDM4 | - | - | - | 1.42E-01 | 1.57E-03 | - | 2.34475E-05 | 6.35E-06 |
| rs7551222 | 1q32.1 | chr1:204599295 | A | G | 2.56E-08 | 1.057 [1.037-1.076] | 0.32 | MDM4 | - | - | - | 1.97E-01 | 1.44E-03 | 3.26E-02 | 0.002385926 | 3.65E-07 |
| rs185666783 | 4q13.2 | chr4:67833774 | G | C | 5.56E-09 | 1.062 [1.042-1.083] | 0.46 | - | - | - | - | 4.92E-05 | 3.34E-02 | 1.66E-04 | 9.20E-08 | 0.00373339 |
| rs35812074 | 5p15.33 | chr5:1267881 | C | G | 4.12E-08 | 1.227 [1.154-1.3] | 0.02 | CLPTM1L,TERT | - | - | - | - | 1.05E-01 | 6.82E-01 | 6.50E-06 | 0.00139271 |
| rs13156167 | 5p15.33 | chr5:1275857 | T | C | 4.08E-14 | 1.121 [1.091-1.151] | 0.15 | CLPTM1L,TERT | - | - | - | 3.10E-23 | 5.13E-01 | 4.79E-01 | 1.02E-10 | 3.68E-05 |
| rs2853677 | 5p15.33 | chr5:1287194 | A | G | 2.87E-29 | 1.114 [1.095-1.133] | 0.41 | CLPTM1L,TERT | - | - | - | 6.90E-32 | 5.42E-01 | 1.74E-01 | 2.66E-18 | 1.44E-12 |
| rs2735947 | 5p15.33 | chr5:1299392 | G | A | 3.48E-13 | 0.904 [0.877-0.931] | 0.14 | CLPTM1L,TERT | - | - | - | 4.01E-16 | 1.08E-01 | 3.77E-02 | 4.32E-11 | 0.000432589 |
| rs2735845 | 5p15.33 | chr5:1300584 | C | G | 3.39E-19 | 1.111 [1.088-1.134] | 0.20 | CLPTM1L,TERT | - | - | - | - | 4.90E-02 | 5.70E-02 | 2.59E-12 | 2.28E-08 |
| rs380286 | 5p15.33 | chr5:1320247 | G | A | 4.93E-42 | 0.882 [0.864-0.9] | 0.44 | CLPTM1L,TERT | - | - | - | 9.89E-22 | 5.81E-20 | 1.33E-03 | 1.51E-32 | 4.51E-12 |
| rs34152723 | 5p15.33 | chr5:1360534 | T | C | 7.26E-10 | 0.893 [0.857-0.929] | 0.07 | CLPTM1L,TERT | - | - | - | 4.00E-06 | 3.42E-04 | 2.23E-01 | 1.68E-08 | 0.0032345 |
| rs7766641 | 6p22.2 | chr6:26184102 | G | A | 7.05E-14 | 0.926 [0.906-0.946] | 0.28 | HIST1H1E,HIST1H2AD, HIST1H2AE,HIST1H2BD, HIST1H2BE,HIST1H2BF, HIST1H2BG,HIST1H3D ,HIST1H3E,HIST1H4D, HIST1H4E | CPD (3.2e-08) | - | - | 4.79E-04 | 6.06E-08 | 2.98E-04 | 1.33E-09 | 1.09E-05 |
| rs6913550 | 6p22.2 | chr6:26540683 | C | T | 4.82E-14 | 0.918 [0.896-0.94] | 0.24 | BTN1A1,HCG11,HMGN4 | - | ZNF322,GUSBP2,HMGN4,ABT1 | ZNF322,HIST1H3E, | 2.09E-02 | 2.26E-03 | 9.33E-05 | 1.70E-07 | 3.16E-08 |
| rs192804591 | 6p22.1 - MHC | chr6:27729706 | T | A | 1.31E-11 | 1.135 [1.098-1.171] | 0.06 | HIST1H2AI,HIST1H2BL, HIST1H3H,LOC100131289 | - | - | - | 6.39E-02 | 8.03E-08 | 4.13E-03 | 2.81E-09 | 0.000482153 |
| rs2531814 | 6p22.1 - MHC | chr6:28425755 | A | G | 1.59E-08 | 0.928 [0.903-0.954] | 0.15 | GPX6,ZSCAN23 | - | ZNF603P,ZSCAN23 | Many | 1.01E-03 | 9.55E-02 | 1.80E-01 | 6.00628E-05 | 5.00E-05 |
| rs116591906 | 6p22.1 - MHC | chr6:28583392 | A | G | 7.43E-11 | 1.113 [1.08-1.145] | 0.12 | ZBED9 | - | - | - | - | 2.35E-06 | 8.04E-03 | 2.09E-09 | 0.0039538 |
| rs9258381 | 6p22.1 - MHC | chr6:29753348 | G | A | 3.06E-20 | 1.112 [1.09-1.135] | 0.14 | HCG4,HLA-F-AS1,HLA-G, IFITM4P,LOC554223 | - | Many | Many | 5.51E-05 | 1.44E-07 | - | 5.99E-14 | 6.42E-08 |
| rs3129813 | 6p22.1 - MHC | chr6:30338247 | G | A | 3.08E-08 | 1.054 [1.035-1.073] | 0.37 | HCG17,HCG18, RPP21,TRIM39, TRIM39-RPP21 | - | HCP5B,HLA-F-AS1 | - | 2.41E-02 | 1.82E-05 | 4.26E-02 | 3.16E-07 | 0.0115202 |
| rs6457312 | 6p21.33 - MHC | chr6:31012941 | A | G | 3.18E-13 | 1.079 [1.058-1.099] | 0.40 | HCG22,MUC22 | - | - | - | 8.10E-01 | 3.19E-08 | 1.96E-03 | 1.61E-06 | 1.11E-08 |
| rs9257054 | 6p21.33 - MHC | chr6:31072949 | C | T | 3.73E-11 | 1.063 [1.045-1.081] | 0.42 | C6orf15,CCHCR1, CDSN,HCG22, PSORS1C1,PSORS1C2 | - | Many | Many | 1.72E-02 | 1.34E-07 | 4.89E-02 | 7.64E-08 | 0.000109655 |
| rs9468936 | 6p21.33 - MHC | chr6:31269884 | T | G | 9.41E-12 | 0.937 [0.919-0.956] | 0.42 | HLA-C | - | Many | Many | 2.29E-03 | 2.43E-05 | 1.01E-02 | 2.47E-07 | 8.14E-06 |
| rs4143333 | 6p21.33 - MHC | chr6:31348519 | A | G | 5.63E-25 | 1.163 [1.134-1.191] | 0.08 | HLA-B,MICA | - | Many | Many | 8.05E-05 | 1.11E-15 | 3.51E-05 | 3.03E-17 | 2.69E-09 |
| rs2516470 | 6p21.33 - MHC | chr6:31407331 | C | G | 2.09E-13 | 0.93 [0.911-0.95] | 0.36 | HCG26,HCP5,MICA | - | Many | Many | - | 5.79E-08 | 3.55E-03 | 4.14E-06 | 2.68E-09 |
| rs3096703 | 6p21.32 - MHC | chr6:32184094 | G | A | 6.18E-09 | 1.055 [1.037-1.073] | 0.43 | AGER,AGPAT1, EGFL8,GPSM3, MIR6721,MIR6833, NOTCH4,PBX2, PPT2-EGFL8,RNF5 ,RNF5P1 | - | Many | Many | 5.54E-01 | 4.84E-07 | 3.97E-03 | 2.39E-06 | 0.000650025 |
| rs3130302 | 6p21.32 - MHC | chr6:32204642 | T | C | 4.05E-09 | 0.941 [0.92-0.961] | 0.27 | GPSM3,NOTCH4,PBX2 | - | - | - | 1.21E-02 | 8.38E-04 | 0.350218516 | 2.13073E-05 | 3.91E-05 |
| rs7753228 | 6p21.32 - MHC | chr6:32503794 | C | T | 3.41E-10 | 1.154 [1.109-1.199] | 0.02 | HLA-DRB1,HLA-DRB5, HLA-DRB6 | - | Many | Many | 4.01E-01 | 2.06E-03 | 1.80E-01 | 0.002309737 | 4.48E-11 |
| rs6904562 | 6p21.32 - MHC | chr6:32556007 | A | T | 1.25E-09 | 1.2 [1.141-1.259] | 0.06 | HLA-DQA1,HLA-DRB1, HLA-DRB6 | - | - | - | 1.59E-02 | 1.14E-03 | 5.34E-02 | 0.000421129 | 3.94E-07 |
| rs62406303 | 6p21.32 - MHC | chr6:32585294 | T | G | 2.28E-09 | 0.926 [0.9-0.951] | 0.21 | HLA-DQA1,HLA-DQB1, HLA-DRB1 | - | - | - | 9.96E-05 | 9.24E-04 | - | 8.44E-08 | 0.00156486 |
| rs28383322 | 6p21.32 - MHC | chr6:32592796 | C | T | 2.21E-10 | 0.929 [0.906-0.952] | 0.23 | HLA-DQA1,HLA-DQB1 ,HLA-DRB1 | - | Many | STK19B,HLA-DRB6 | 7.94E-03 | 1.47E-09 | 1.61E-01 | 1.53E-08 | 0.00174767 |
| rs9272306 | 6p21.32 - MHC | chr6:32603880 | C | T | 2.85E-12 | 1.1 [1.073-1.126] | 0.05 | HLA-DQA1,HLA-DQB1 ,HLA-DRB1 | - | Many | Many |  | 1.25E-08 | 2.94E-03 | 2.61E-08 | 9.82E-06 |
| rs9272327 | 6p21.32 - MHC | chr6:32604188 | G | A | 4.13E-08 | 1.074 [1.048-1.099] | 0.08 | HLA-DQA1,HLA-DQB1, HLA-DRB1 | - | Many | Many | 9.20E-02 | 3.67E-08 | 2.72E-03 | 6.44E-08 | 0.099087 |
| rs1794514 | 6p21.32 - MHC | chr6:32667473 | G | C | 7.55E-33 | 1.147 [1.124-1.169] | 0.17 | HLA-DQA2,HLA-DQB1 | - | Many | Many | 5.19E-02 | 2.45E-15 | 1.78E-04 | 3.61E-13 | 3.55E-22 |
| rs2621403 | 6p21.32 - MHC | chr6:32749795 | A | T | 1.60E-09 | 0.944 [0.925-0.963] | 0.37 | HLA-DOB,HLA-DQA2 ,HLA-DQB2,TAP2 | - | HLA-DOB,HLA-DQB1,HLA-DRB5 | HLA-DOB,HLA-DQB1,C4A | 1.34E-01 | 6.79E-08 | 3.47E-03 | 1.65E-06 | 0.000243657 |
| rs239933 | 6q27 | chr6:167413230 | A | G | 6.70E-12 | 0.939 [0.921-0.957] | 0.44 | FGFR1OP,MIR3939 ,RNASET2 | - | RNASET2, FGFR1OP | RNASET2 | 1.24E-05 | 1.15E-02 | 3.89E-02 | 1.56E-07 | 9.41E-06 |
| rs11778371 | 8p21.2 | chr8:27319905 | C | T | 4.36E-10 | 0.881 [0.841-0.921] | 0.07 | CHRNA2,EPHX2, MIR6842,PTK2B | AgeofIn(3.12e-09) smkIn (1.49e-07) | - | CHRNA2, TRIM35 | 3.00E-05 | 8.53E-03 | 3.68E-02 | 1.14E-07 | 0.000722484 |
| rs10091679 | 8p21.2 | chr8:27351738 | G | A | 4.75E-08 | 1.087 [1.057-1.117] | 0.09 | CHRNA2,EPHX2, PTK2B | - | - | - | 5.86E-04 | 5.29E-04 | 9.32E-02 | 5.02E-07 | 0.0121681 |
| rs6558008 | 8p21.1 | chr8:27438306 | A | C | 8.15E-12 | 0.928 [0.906-0.949] | 0.23 | CLU,EPHX2,MIR6843 | smkIn(8.76e-11) | - | - | 2.67E-04 | 3.36E-03 | - | 8.48E-07 | 1.75E-06 |
| rs1333047 | 9p21.3 | chr9:22124504 | A | T | 9.57E-09 | 0.949 [0.93-0.967] | 0.48 | CDKN2B-AS1 | - | - | - | 2.72E-03 | 7.51E-05 | 6.78E-01 | 0.000415693 | 2.13E-06 |
| rs72905558 | 11p11.2 | chr11:48201643 | A | T | 2.41E-09 | 0.913 [0.883-0.943] | 0.14 | OR4B1,PTPRJ | - | C1QTNF4 | MTCH2 | 1.50E-03 | 2.22E-01 | 2.51E-03 | 2.30247E-05 | 2.50E-05 |
| rs61612408 | 11q24.2 | chr11:125495044 | G | A | 3.07E-08 | 0.903 [0.867-0.939] | 0.07 | ACRV1,CHEK1, EI24,STT3A | - | CHEK1 | CHEK1 | 1.40E-01 | 1.15E-05 | 7.26E-02 | 0.00018486 | 2.74E-05 |
| rs7953330 | 12p13.33 | chr12:998819 | G | C | 1.30E-18 | 0.916 [0.897-0.936] | 0.29 | RAD52,WNK1 | - | RAD52 | RAD52 | 3.70E-02 | 7.26E-13 | - | 6.10E-12 | 3.80E-08 |
| rs7487683 | 12p13.33 | chr12:1036042 | C | T | 4.98E-11 | 0.844 [0.793-0.894] | 0.03 | RAD52,WNK1 | - | - | - | 8.00E-03 | 8.48E-05 | 2.61E-05 | 6.58E-08 | 0.000126012 |
| rs11571734 | 13q13.1 | chr13:32940888 | C | T | 9.24E-09 | 0.943 [0.923-0.963] | 0.28 | BRCA2,N4BP2L1 | - | BRCA2 | - | 4.12E-04 | - | - | 1.78E-06 | 0.00123155 |
| rs11571815 | 13q13.1 | chr13:32968550 | G | A | 6.98E-32 | 1.695 [1.607-1.783] | 0.01 | BRCA2,N4BP2L1, N4BP2L2 | - | - | - | 1.93E-03 | 1.10E-15 | - | 6.69E-16 | 4.37E-18 |
| rs506120 | 15q15.3 | chr15:43802024 | C | T | 6.37E-11 | 0.936 [0.916-0.956] | 0.30 | MAP1A,PPIP5K1, RNU6-28P,TP53BP1 | - | Many | Many | 4.28E-02 | 1.67E-03 | 4.34E-01 | 0.000149895 | 1.24E-08 |
| rs66759488 | 15q21.1 | chr15:47577451 | G | A | 3.72E-10 | 1.061 [1.042-1.08] | 0.36 | SEMA6D | - | - | - | 1.95E-03 | 1.36E-03 | 4.98E-04 | 2.83E-08 | 0.00185167 |
| rs77468143 | 15q21.1 | chr15:49376624 | T | G | 3.46E-08 | 0.943 [0.923-0.964] | 0.28 | COPS2,SECISBP2L | - | SECISBP2L,FAM227B,RP11-295H24.3 | - | 1.69E-16 | 6.79E-01 | 3.60E-01 | 1.00E-09 | 0.235845 |
| rs12441817 | 15q24.1 | chr15:75025814 | T | C | 4.77E-08 | 1.096 [1.063-1.129] | 0.08 | CSK,CYP1A1, CYP1A2,EDC3 | - | RP11-10O17.1 | CYP1A1,SEMA7A | 2.19E-04 | - | 0.4430523075 | 5.28E-06 | 0.00216923 |
| rs16969892 | 15q25.1 | chr15:78774737 | A | G | 3.71E-08 | 0.835 [0.771-0.899] | 0.03 | HYKK,IREB2 | CPD (6.52e-09) | - | - | 5.76E-02 | 3.44E-05 | 5.47E-03 | 6.23E-07 | 0.00888396 |
| rs117131212 | 15q25.1 | chr15:78819478 | A | G | 3.01E-08 | 1.138 [1.092-1.183] | 0.06 | CHRNA5,HYKK,IREB2,PSMA4 | - | - | - | 1.24E-02 | 5.02E-04 | 3.05E-03 | 7.42E-07 | 0.0086308 |
| rs55781567 | 15q25.1 | chr15:78857986 | C | G | 4.59E-115 | 1.241 [1.222-1.259] | 0.37 | CHRNA3,CHRNA5, HYKK,PSMA4 | CPD (1.03e-206) Cess (1.93e-08) | CHRNA5 | CHRNA5,CHRNA3,PSMA4 | 2.83E-48 | 4.61E-41 | 9.13E-21 | 3.08E-103 | 2.69E-21 |
| rs2229961 | 15q25.1 | chr15:78880752 | G | A | 9.50E-16 | 1.325 [1.257-1.394] | 0.01 | CHRNA3,CHRNA5, CHRNB4,PSMA4 | CPD (1.17e-18) | - | - | 1.33E-09 | 7.89E-09 | 3.74E-02 | 5.01E-14 | 0.000237387 |
| rs76474922 | 15q25.1 | chr15:78884553 | A | C | 1.79E-23 | 0.838 [0.803-0.873] | 0.09 | CHRNA3,CHRNA5 ,CHRNB4,PSMA4 | CPD (5.86e-60) | CHRNA5 | - | 3.47E-08 | 5.68E-15 | 1.04E-04 | 6.13E-21 | 6.35E-06 |
| rs12443170 | 15q25.1 | chr15:78907736 | G | A | 3.65E-40 | 0.828 [0.8-0.856] | 0.13 | CHRNA3,CHRNA5, CHRNB4 | CPD (1.71e-48) | - | CHRNA3 | 2.49E-13 | 9.31E-14 | 6.53E-09 | 9.17E-34 | 4.48E-10 |
| rs8192479 | 15q25.1 | chr15:78909398 | C | T | 3.78E-08 | 1.202 [1.136-1.268] | 0.02 | CHRNA3,CHRNA5, CHRNB4 | CPD (6.83e-10) | - | CTSH | 2.03E-05 | 5.40E-07 | 1.98E-02 | 2.29E-08 | 0.0498391 |
| rs151118057 | 15q25.1 | chr15:78988620 | G | A | 1.04E-11 | 0.772 [0.697-0.847] | 0.01 | . | - | - | - | 1.13E-03 | 1.60E-03 | 1.44E-03 | 9.56E-09 | 0.000149456 |
| rs28406853 | 15q25.1 | chr15:79034874 | A | G | 4.17E-08 | 1.128 [1.085-1.172] | 0.04 | ADAMTS7,LOC646938 | - | - | - | 9.86E-05 | 2.01E-05 | 4.33E-01 | 4.24E-06 | 0.00246041 |
| rs7177201 | 15q25.1 | chr15:79065380 | C | T | 3.80E-24 | 0.892 [0.87-0.914] | 0.25 | ADAMTS7,LOC646938 | - | - | - | 2.86E-09 | - | 3.95E-03 | 1.52E-16 | 2.57E-09 |
| rs11638020 | 15q25.1 | chr15:79107724 | G | A | 1.37E-16 | 1.08 [1.062-1.099] | 0.39 | ADAMTS7 | CPD (7.35e-35) | - | ADAMTS7 | 1.49E-09 | 3.18E-07 | 5.26E-02 | 3.22E-14 | 0.000165819 |
| rs3865453 | 19q13.2 | chr19:41338556 | C | T | 9.84E-14 | 0.868 [0.831-0.905] | 0.08 | CYP2A6,CYP2A7 | CPD (3.56e-23) Cess (5.26e-10) | CYP2G1P | - | 3.06E-05 | 2.13E-05 | 7.61E-03 | 3.46E-09 | 5.57E-06 |
| rs56113850 | 19q13.2 | chr19:41353107 | C | T | 1.50E-20 | 0.911 [0.891-0.93] | 0.41 | CYP2A6,CYP2A7 | CPD (1.1e-81) Cess (2.52e-26) | CYP2A7,CYP2A6,CTC-490E21.11 | CYP2A7 | 8.64E-10 | 7.18E-11 | 1.14E-03 | 5.02E-19 | 4.50E-05 |
| rs1801272 | 19q13.2 | chr19:41354533 | A | T | 3.77E-10 | 0.825 [0.765-0.885] | 0.03 | CYP2A6,CYP2A7 | CPD (2.05e-38) Cess (1.92e-10) | - | - | 1.52E-02 | 4.18E-05 | 1.72E-02 | 2.37E-07 | 0.000332828 |
| rs11697662 | 20q13.33 | chr20:61992005 | T | C | 1.49E-08 | 1.071 [1.047-1.094] | 0.20 | CHRNA4 | CPD (1.33e-15) Cess (9.82e-15) | - | CHRNA4 | 6.42E-02 | 5.50E-06 | 3.40E-03 | 1.58E-06 | 0.00201967 |
| rs2281925 | 20q13.33 | chr20:62376503 | G | A | 3.49E-09 | 1.091 [1.062-1.12] | 0.10 | RTEL1 | - | - | - | 3.05E-08 | 9.46E-01 | 5.34E-02 | 1.52319E-05 | 5.65E-05 |

Smoking related SNP, Genome-wide significant in GSCAN, cessation (Cess), cigarettes per day (CPD), age of imitation. GTEx analysis was perform on the GTEx variant page for single-tissue eQTLs  with P-value < 0.05, Adeno = Adenocarcinoma, Small = Small cell carcinoma, Squam = Squamous cell carcinoma

**Supplementary Table 3 - eQTL analysis on rs78062588 and rs11697662**

| **rsID** | **Gene Symbol** | **GTEx Variant Id** | **P-Value** | **NES** | **Tissue** |
| --- | --- | --- | --- | --- | --- |
| rs78062588 | CHRNB2 | chr1_154593749_T_C_b38 | 0.0000092 | -0.27 | Brain - Cerebellum |
| **rs78062588** | **CHRNB2** | **chr1_154593749_T_C_b38** | **0.000012** | **-0.21** | **Brain - Putamen (basal ganglia)** |
| rs78062588 | CHRNB2 | chr1_154593749_T_C_b38 | 0.000014 | -0.29 | Brain - Cerebellar Hemisphere |
| rs78062588 | CHRNB2 | chr1_154593749_T_C_b38 | 0.000053 | -0.18 | Brain - Caudate (basal ganglia) |
| rs11697662 | CHRNA4 | chr20_63360653_C_T_b38 | 2.50E-10 | 0.44 | Brain - Nucleus accumbens (basal ganglia) |
| rs11697662 | CHRNA4 | chr20_63360653_C_T_b38 | 3.60E-09 | -0.57 | Liver |
| **rs11697662** | **CHRNA4** | **chr20_63360653_C_T_b38** | **8.40E-09** | **0.5** | **Brain - Putamen (basal ganglia)** |
| rs11697662 | CHRNA4 | chr20_63360653_C_T_b38 | 1.00E-07 | 0.2 | Testis |
| rs11697662 | CHRNA4 | chr20_63360653_C_T_b38 | 0.0000012 | 0.53 | Brain - Spinal cord (cervical c-1) |

**Supplementary Table 4 - PRS panels for smPRS and eQTLPRS**

| **SNP** | **CHROM** | **POS** | **REF** | **ALT** | **beta_LC** | **P_LC** | **SE_LC** | **Z_LC** | **MAF** | **PRS_PANEL** |
| --- | --- | --- | --- | --- | --- | --- | --- | --- | --- | --- |
| rs72740955 | 15 | 78849779 | C | T | 0.215883 | 6.471E-115 | 0.00947481 | 22.7849424 | 0.3712 | smPRS |
| rs12443170 | 15 | 78907736 | G | A | -0.188852 | 3.6517E-40 | 0.014236 | -13.265805 | 0.1278 | smPRS |
| rs76474922 | 15 | 78884553 | A | C | -0.176936 | 1.7938E-23 | 0.0177223 | -9.9838057 | 0.09407 | smPRS |
| rs2002854 | 15 | 79053014 | G | A | -0.108097 | 1.192E-21 | 0.0113087 | -9.5587468 | 0.2526 | smPRS |
| rs56113850 | 19 | 41353107 | C | T | -0.0934494 | 1.4969E-20 | 0.0100557 | -9.293177 | 0.407 | smPRS |
| rs7164479 | 15 | 79123054 | T | C | 0.0789763 | 1.541E-17 | 0.00926514 | 8.52402662 | 0.4192 | smPRS |
| rs2229961 | 15 | 78880752 | G | A | 0.281776 | 9.5014E-16 | 0.0350767 | 8.03313881 | 0.01125 | smPRS |
| rs7766641 | 6 | 26184102 | G | A | -0.07698 | 7.0504E-14 | 0.0102819 | -7.4869431 | 0.2843 | smPRS |
| rs3865453 | 19 | 41338556 | C | T | -0.141392 | 9.8445E-14 | 0.0189967 | -7.4429769 | 0.08384 | smPRS |
| rs6558008 | 8 | 27438306 | A | C | -0.074914 | 8.1472E-12 | 0.0109588 | -6.8359674 | 0.2331 | smPRS |
| rs34517439 | 1 | 78450517 | C | A | 0.0995982 | 1.8351E-11 | 0.0148243 | 6.71857693 | 0.091 | smPRS |
| rs3734523 | 6 | 25925987 | G | A | 0.0958541 | 7.7956E-11 | 0.0147366 | 6.50449222 | 0.08691 | smPRS |
| rs11778371 | 8 | 27319905 | C | T | -0.126536 | 4.3552E-10 | 0.0202758 | -6.2407402 | 0.06748 | smPRS |
| rs35255262 | 15 | 47569263 | A | C | 0.0580925 | 5.2917E-10 | 0.00935436 | 6.21020572 | 0.3609 | smPRS |
| rs11671669 | 19 | 41324392 | G | A | -0.169533 | 6.7881E-10 | 0.0274727 | -6.1709624 | 0.03988 | smPRS |
| rs146722791 | 15 | 79140191 | C | T | 0.110197 | 1.2859E-08 | 0.0193738 | 5.68793938 | 0.07362 | smPRS |
| rs11697662 | 20 | 61992005 | T | C | 0.0683118 | 1.488E-08 | 0.012063 | 5.66291967 | 0.2004 | smPRS |
| rs16969892 | 15 | 78774737 | A | G | -0.180319 | 3.7085E-08 | 0.0327603 | -5.5041926 | 0.02863 | smPRS |
| rs8192479 | 15 | 78909398 | C | T | 0.183958 | 3.7777E-08 | 0.0334411 | 5.50095541 | 0.02249 | smPRS |
| rs78062588 | 1 | 154566225 | T | C | -0.101165 | 4.0285E-08 | 0.0184285 | -5.4895949 | 0.06851 | smPRS |
| rs13296519 | 9 | 128471924 | G | T | 0.0502667 | 9.7312E-08 | 0.00942794 | 5.33167373 | 0.3937 | smPRS |
| rs705698 | 12 | 56384687 | T | C | -0.0509268 | 1.8988E-07 | 0.00977672 | -5.2089862 | 0.32 | smPRS |
| rs215622 | 7 | 32357659 | T | C | 0.0494946 | 2.2925E-07 | 0.0095662 | 5.17390395 | 0.32 | smPRS |
| rs4774495 | 15 | 47651362 | T | C | 0.0703162 | 2.9419E-07 | 0.0137145 | 5.12714281 | 0.1207 | smPRS |
| rs77802411 | 15 | 78925414 | T | C | 0.144154 | 5.4322E-07 | 0.0287711 | 5.01037499 | 0.03272 | smPRS |
| rs113864525 | 9 | 136473053 | C | A | 0.070263 | 1.8093E-06 | 0.014719 | 4.77362593 | 0.091 | smPRS |
| rs3025383 | 9 | 136502369 | T | C | -0.056208 | 2.2056E-06 | 0.0118742 | -4.7336242 | 0.181 | smPRS |
| rs11162504 | 1 | 78991197 | A | G | 0.0492613 | 2.2232E-06 | 0.0104102 | 4.73202244 | 0.2485 | smPRS |
| rs2152113 | 1 | 43983569 | C | T | -0.0445074 | 2.4101E-06 | 0.00943835 | -4.7155912 | 0.3558 | smPRS |
| rs79368540 | 2 | 45189737 | C | T | 0.0662498 | 2.4757E-06 | 0.0140654 | 4.71012556 | 0.1043 | smPRS |
| rs6704753 | 2 | 186031626 | T | C | 0.0547779 | 2.5413E-06 | 0.011643 | 4.70479258 | 0.1861 | smPRS |
| rs17087844 | 4 | 67863736 | T | C | 0.0426672 | 3.0932E-06 | 0.00914715 | 4.66453486 | 0.4591 | smPRS |
| rs2002403 | 15 | 78681002 | G | A | -0.175704 | 4.7042E-06 | 0.0383837 | -4.5775681 | 0.01431 | smPRS |
| rs74024464 | 15 | 79125155 | G | A | -0.0953825 | 5.2432E-06 | 0.020941 | -4.5548207 | 0.0501 | smPRS |
| rs6603950 | 1 | 77938087 | G | A | -0.0406734 | 8.2406E-06 | 0.00912198 | -4.4588346 | 0.4714 | smPRS |
| rs2728125 | 4 | 89001893 | A | G | -0.0684567 | 1.2656E-05 | 0.0156796 | -4.3659723 | 0.0726 | smPRS |
| rs1541965 | 15 | 79205126 | G | A | 0.0903501 | 1.439E-05 | 0.0208285 | 4.33781117 | 0.04601 | smPRS |
| rs9328534 | 9 | 134874805 | T | C | 0.0419043 | 1.6513E-05 | 0.00972827 | 4.30747707 | 0.319 | smPRS |
| rs4737074 | 8 | 42663781 | C | T | -0.0474014 | 1.7196E-05 | 0.0110274 | -4.298511 | 0.2311 | smPRS |
| rs187618975 | 15 | 78818072 | C | T | 0.178965 | 1.9871E-05 | 0.0419481 | 4.26634341 | 0.01636 | smPRS |
| rs13412382 | 2 | 146027193 | G | A | 0.0395135 | 1.9978E-05 | 0.00926431 | 4.26513146 | 0.4162 | smPRS |
| rs182364018 | 19 | 41447851 | T | C | -0.264052 | 2.0582E-05 | 0.0620061 | -4.2584842 | 0.01022 | smPRS |
| rs2519150 | 9 | 136491172 | T | C | -0.0388444 | 2.1205E-05 | 0.00913599 | -4.2517998 | 0.4939 | smPRS |
| rs329122 | 5 | 133864599 | G | A | -0.0386908 | 2.5287E-05 | 0.00918535 | -4.2122293 | 0.4294 | smPRS |
| rs910928 | 20 | 61720769 | C | T | -0.0382785 | 3.3593E-05 | 0.009229 | -4.1476325 | 0.4376 | smPRS |
| rs1123893 | 7 | 32446930 | T | C | -0.0470571 | 3.402E-05 | 0.0113535 | -4.1447219 | 0.2035 | smPRS |
| rs2604909 | 19 | 41297615 | C | T | 0.0453975 | 3.983E-05 | 0.0110497 | 4.10848258 | 0.2209 | smPRS |
| rs7169340 | 15 | 47614645 | T | C | 0.0471704 | 4.2227E-05 | 0.0115192 | 4.09493715 | 0.1789 | smPRS |
| rs912779 | 13 | 67336287 | C | T | -0.0420798 | 4.4261E-05 | 0.0103035 | -4.0840297 | 0.2464 | smPRS |
| rs55731474 | 14 | 103878481 | G | A | 0.0387202 | 4.6768E-05 | 0.00951072 | 4.07121648 | 0.3517 | smPRS |
| rs1459066 | 5 | 167044666 | T | C | 0.0403557 | 4.8676E-05 | 0.00993519 | 4.06189514 | 0.2883 | smPRS |
| rs3130160 | 6 | 33124972 | A | G | 0.0405025 | 5.4598E-05 | 0.0100377 | 4.03503791 | 0.271 | smPRS |
| rs57339473 | 19 | 41452921 | C | T | -0.0570477 | 5.7127E-05 | 0.0141755 | -4.0243871 | 0.1411 | smPRS |
| rs72745113 | 15 | 79008966 | G | A | 0.122895 | 6.54E-05 | 0.0307821 | 3.99241767 | 0.03579 | smPRS |
| rs176415 | 2 | 32649778 | G | A | 0.0381724 | 7.7663E-05 | 0.00966023 | 3.95150012 | 0.3211 | smPRS |
| rs7689452 | 4 | 147945733 | A | G | -0.0387424 | 7.8189E-05 | 0.00980848 | -3.9498883 | 0.2945 | smPRS |
| rs79183029 | 19 | 41298228 | C | T | 0.10803 | 8.3645E-05 | 0.0274625 | 3.93372781 | 0.0184 | smPRS |
| rs12802244 | 11 | 47932666 | G | A | -0.0376109 | 0.00010468 | 0.00969481 | -3.8794881 | 0.3865 | smPRS |
| rs13107325 | 4 | 103188709 | C | T | 0.0672244 | 0.00012177 | 0.0174948 | 3.84253607 | 0.07669 | smPRS |
| rs10266703 | 7 | 1918941 | C | A | 0.035212 | 0.00013393 | 0.00921991 | 3.81912622 | 0.4366 | smPRS |
| rs12971445 | 19 | 41484602 | G | T | -0.0362472 | 0.00016003 | 0.00960198 | -3.7749714 | 0.3528 | smPRS |
| rs828097 | 9 | 127959449 | C | T | -0.0379361 | 0.00016443 | 0.0100674 | -3.7682122 | 0.2996 | smPRS |
| rs10468280 | 16 | 53827479 | A | G | -0.0349903 | 0.00016601 | 0.0092916 | -3.7657992 | 0.4131 | smPRS |
| rs2540034 | 16 | 4022694 | T | C | -0.0353348 | 0.00018781 | 0.00946077 | -3.7348757 | 0.4325 | smPRS |
| rs10123941 | 9 | 120518162 | T | C | 0.0378936 | 0.0002137 | 0.0102353 | 3.70224615 | 0.271 | smPRS |
| rs58164846 | 19 | 41140115 | G | A | -0.0422373 | 0.00021984 | 0.0114307 | -3.6950755 | 0.228 | smPRS |
| rs748283 | 8 | 27317195 | C | A | -0.0336755 | 0.00023672 | 0.00916038 | -3.6762121 | 0.4765 | smPRS |
| rs12643257 | 4 | 106076749 | A | G | 0.0344643 | 0.00024067 | 0.00938573 | 3.67198929 |  | smPRS |
| rs117527479 | 11 | 48705469 | G | A | 0.109266 | 0.00024925 | 0.0298295 | 3.66301815 | 0.03681 | smPRS |
| rs11136000 | 8 | 27464519 | C | T | 0.0338558 | 0.00025758 | 0.00926387 | 3.65460655 | 0.3896 | smPRS |
| rs7638643 | 3 | 49565842 | C | T | -0.0485497 | 0.00033413 | 0.0135338 | -3.5872926 | 0.1462 | smPRS |
| rs11125927 | 2 | 62752975 | A | G | -0.0518088 | 0.00034851 | 0.0144868 | -3.5762763 | 0.1278 | smPRS |
| rs673345 | 9 | 120716221 | A | G | 0.033259 | 0.00034886 | 0.00930055 | 3.57602507 | 0.3896 | smPRS |
| rs35967356 | 16 | 69581354 | C | T | -0.0329464 | 0.00036995 | 0.00925292 | -3.560649 | 0.4192 | smPRS |
| rs3801788 | 7 | 111378299 | G | A | -0.0344521 | 0.00039181 | 0.00971703 | -3.5455381 | 0.317 | smPRS |
| rs60799361 | 8 | 27558931 | T | C | -0.0477369 | 0.00046929 | 0.0136481 | -3.4976956 | 0.1258 | smPRS |
| rs68136852 | 17 | 27382061 | C | A | 0.0446874 | 0.00047386 | 0.0127857 | 3.49510782 | 0.1575 | smPRS |
| rs12693971 | 2 | 203415977 | T | C | 0.0363506 | 0.00049393 | 0.0104335 | 3.48402741 | 0.2607 | smPRS |
| rs7656078 | 4 | 24489441 | G | T | -0.0318125 | 0.00050985 | 0.00915328 | -3.4755301 | 0.4448 | smPRS |
| rs117683060 | 19 | 41394102 | A | G | -0.115907 | 0.0005441 | 0.0335182 | -3.4580318 | 0.02147 | smPRS |
| rs17398598 | 1 | 52024908 | A | C | -0.0362265 | 0.00057857 | 0.0105265 | -3.4414573 | 0.2413 | smPRS |
| rs6684973 | 1 | 73801632 | G | A | 0.0324075 | 0.00059537 | 0.00943804 | 3.43371081 | 0.3937 | smPRS |
| rs10939621 | 4 | 15578068 | C | T | 0.0314223 | 0.00060478 | 0.00916246 | 3.42946108 | 0.4305 | smPRS |
| rs150208378 | 4 | 100850288 | C | T | -0.163175 | 0.00063615 | 0.0477719 | -3.4157109 | 0.005112 | smPRS |
| rs117826664 | 8 | 88684691 | C | T | 0.0813009 | 0.00063845 | 0.0238089 | 3.41472727 | 0.0317 | smPRS |
| rs115552537 | 1 | 91190856 | C | A | -0.0382499 | 0.00065957 | 0.0112306 | -3.4058643 | 0.1984 | smPRS |
| rs7678161 | 4 | 2901600 | C | T | -0.0334674 | 0.00066993 | 0.00983872 | -3.401601 | 0.3395 | smPRS |
| rs78997851 | 2 | 148977496 | T | G | 0.0591774 | 0.00075053 | 0.0175578 | 3.37043365 | 0.0726 | smPRS |
| rs6741157 | 2 | 104421850 | C | T | 0.0305737 | 0.00079822 | 0.0091172 | 3.35340894 | 0.4928 | smPRS |
| rs140895602 | 6 | 31024244 | A | G | -0.105085 | 0.00081432 | 0.0313885 | -3.3478822 | 0.03374 | smPRS |
| rs719319 | 7 | 125712737 | T | C | 0.0314185 | 0.00086336 | 0.00943035 | 3.33163668 | 0.3793 | smPRS |
| rs7507400 | 19 | 41330179 | G | T | -0.0406817 | 0.00091609 | 0.0122716 | -3.3151097 | 0.316 | smPRS |
| rs739190 | 22 | 37758272 | C | T | -0.0765114 | 0.00093698 | 0.0231236 | -3.3088014 | 0.03783 | smPRS |
| rs28411352 | 1 | 38278579 | C | T | -0.0356502 | 0.00100515 | 0.0108389 | -3.2890976 | 0.2546 | smPRS |
| rs9870893 | 3 | 16863267 | T | G | 0.0307041 | 0.00111431 | 0.00941856 | 3.25995694 | 0.3589 | smPRS |
| rs3133877 | 11 | 132148460 | A | G | 0.0338505 | 0.00114037 | 0.0104047 | 3.25338549 | 0.2883 | smPRS |
| rs150353 | 15 | 89928189 | T | G | -0.0297439 | 0.00115586 | 0.00915322 | -3.2495559 | 0.4325 | smPRS |
| rs12738089 | 1 | 154604171 | A | G | 0.0347654 | 0.0011698 | 0.0107098 | 3.24612971 | 0.271 | smPRS |
| rs10832569 | 11 | 16249453 | T | G | -0.0297915 | 0.00119604 | 0.00919541 | -3.2398229 | 0.4724 | smPRS |
| rs4939031 | 11 | 56056452 | C | A | -0.0393164 | 0.00120558 | 0.0121438 | -3.2375698 | 0.1636 | smPRS |
| rs72740955 | 15 | 78849779 | C | T | 0.215883 | 6.471E-115 | 0.00947481 | 22.7849424 | 0.3712 | eqtlPRS |
| rs380286 | 5 | 1320247 | G | A | -0.125426 | 4.9336E-42 | 0.00923285 | -13.584754 | 0.4366 | eqtlPRS |
| rs12443170 | 15 | 78907736 | G | A | -0.188852 | 3.6517E-40 | 0.014236 | -13.265805 | 0.1278 | eqtlPRS |
| rs2853677 | 5 | 1287194 | A | G | 0.107761 | 2.8684E-29 | 0.00959484 | 11.2311409 | 0.411 | eqtlPRS |
| rs76474922 | 15 | 78884553 | A | C | -0.176936 | 1.7938E-23 | 0.0177223 | -9.9838057 | 0.09407 | eqtlPRS |
| rs2002854 | 15 | 79053014 | G | A | -0.108097 | 1.192E-21 | 0.0113087 | -9.5587468 | 0.2526 | eqtlPRS |
| rs56113850 | 19 | 41353107 | C | T | -0.0934494 | 1.4969E-20 | 0.0100557 | -9.293177 | 0.407 | eqtlPRS |
| rs7164479 | 15 | 79123054 | T | C | 0.0789763 | 1.541E-17 | 0.00926514 | 8.52402662 | 0.4192 | eqtlPRS |
| rs147560086 | 12 | 1054956 | C | T | -0.0995753 | 2.7538E-17 | 0.0117749 | -8.4565729 | 0.1851 | eqtlPRS |
| rs2229961 | 15 | 78880752 | G | A | 0.281776 | 9.5014E-16 | 0.0350767 | 8.03313881 | 0.01125 | eqtlPRS |
| rs6913550 | 6 | 26540683 | C | T | -0.0853457 | 4.8238E-14 | 0.0113242 | -7.5365765 | 0.2393 | eqtlPRS |
| rs7766641 | 6 | 26184102 | G | A | -0.07698 | 7.0504E-14 | 0.0102819 | -7.4869431 | 0.2843 | eqtlPRS |
| rs3865453 | 19 | 41338556 | C | T | -0.141392 | 9.8445E-14 | 0.0189967 | -7.4429769 | 0.08384 | eqtlPRS |
| rs2735947 | 5 | 1299392 | G | A | -0.100852 | 3.476E-13 | 0.0138637 | -7.2745371 | 0.136 | eqtlPRS |
| rs27919 | 5 | 1361888 | C | T | 0.105569 | 3.5588E-12 | 0.0151818 | 6.95365503 | 0.1166 | eqtlPRS |
| rs239933 | 6 | 167413230 | A | G | -0.0628235 | 6.6986E-12 | 0.00915268 | -6.8639459 | 0.4417 | eqtlPRS |
| rs6558008 | 8 | 27438306 | A | C | -0.074914 | 8.1472E-12 | 0.0109588 | -6.8359674 | 0.2331 | eqtlPRS |
| rs151118057 | 15 | 78988620 | G | A | -0.258753 | 1.0369E-11 | 0.0380446 | -6.8013069 | 0.01431 | eqtlPRS |
| rs34517439 | 1 | 78450517 | C | A | 0.0995982 | 1.8351E-11 | 0.0148243 | 6.71857693 | 0.091 | eqtlPRS |
| rs7734992 | 5 | 1280128 | T | C | 0.0650181 | 3.6452E-11 | 0.00982469 | 6.61782713 | 0.4427 | eqtlPRS |
| rs506120 | 15 | 43802024 | C | T | -0.0659654 | 6.3655E-11 | 0.0100943 | -6.5349157 | 0.2975 | eqtlPRS |
| rs3734523 | 6 | 25925987 | G | A | 0.0958541 | 7.7956E-11 | 0.0147366 | 6.50449222 | 0.08691 | eqtlPRS |
| rs28383322 | 6 | 32592796 | C | T | -0.0736974 | 2.2109E-10 | 0.0116133 | -6.3459482 | 0.2321 | eqtlPRS |
| rs11778371 | 8 | 27319905 | C | T | -0.126536 | 4.3552E-10 | 0.0202758 | -6.2407402 | 0.06748 | eqtlPRS |
| rs11671669 | 19 | 41324392 | G | A | -0.169533 | 6.7881E-10 | 0.0274727 | -6.1709624 | 0.03988 | eqtlPRS |
| rs34152723 | 5 | 1360534 | T | C | -0.113054 | 7.2575E-10 | 0.0183518 | -6.1603766 | 0.06748 | eqtlPRS |
| rs4252707 | 1 | 204508147 | G | A | -0.0719435 | 9.1057E-10 | 0.0117471 | -6.1243626 | 0.2209 | eqtlPRS |
| rs2281925 | 20 | 62376503 | G | A | 0.086965 | 3.4894E-09 | 0.014723 | 5.90674455 | 0.09918 | eqtlPRS |
| rs146722791 | 15 | 79140191 | C | T | 0.110197 | 1.2859E-08 | 0.0193738 | 5.68793938 | 0.07362 | eqtlPRS |
| rs11697662 | 20 | 61992005 | T | C | 0.0683118 | 1.488E-08 | 0.012063 | 5.66291967 | 0.2004 | eqtlPRS |
| rs2531814 | 6 | 28425755 | A | G | -0.0742335 | 1.5882E-08 | 0.0131346 | -5.6517519 | 0.1544 | eqtlPRS |
| rs7551222 | 1 | 204599295 | A | G | 0.0552433 | 2.5569E-08 | 0.00991916 | 5.56935265 | 0.319 | eqtlPRS |
| rs61612408 | 11 | 125495044 | G | A | -0.101955 | 3.0747E-08 | 0.0184129 | -5.5371506 | 0.07362 | eqtlPRS |
| rs77468143 | 15 | 49376624 | T | G | -0.0583901 | 3.4552E-08 | 0.0105843 | -5.5166709 | 0.2832 | eqtlPRS |
| rs16969892 | 15 | 78774737 | A | G | -0.180319 | 3.7085E-08 | 0.0327603 | -5.5041926 | 0.02863 | eqtlPRS |
| rs8192479 | 15 | 78909398 | C | T | 0.183958 | 3.7777E-08 | 0.0334411 | 5.50095541 | 0.02249 | eqtlPRS |
| rs78062588 | 1 | 154566225 | T | C | -0.101165 | 4.0285E-08 | 0.0184285 | -5.4895949 | 0.06851 | eqtlPRS |
| rs10091679 | 8 | 27351738 | G | A | 0.0834697 | 4.7546E-08 | 0.0152868 | 5.46024675 | 0.091 | eqtlPRS |
| rs12441817 | 15 | 75025814 | T | C | 0.0920115 | 4.7685E-08 | 0.0168527 | 5.45974829 | 0.08384 | eqtlPRS |
| rs10061756 | 5 | 1630142 | G | A | -0.0599337 | 6.01E-08 | 0.0110609 | -5.4185193 | 0.2352 | eqtlPRS |
| rs9311380 | 3 | 45996047 | C | T | 0.048509 | 9.0874E-08 | 0.00907714 | 5.34408415 | 0.4734 | eqtlPRS |
| rs1629083 | 11 | 118126576 | T | C | 0.0488717 | 9.2198E-08 | 0.0091495 | 5.34146128 | 0.499 | eqtlPRS |
| rs35640778 | 20 | 62321128 | G | A | -0.189 | 9.3575E-08 | 0.0354013 | -5.338787 | 0.01227 | eqtlPRS |
| rs13296519 | 9 | 128471924 | G | T | 0.0502667 | 9.7312E-08 | 0.00942794 | 5.33167373 | 0.3937 | eqtlPRS |
| rs10764672 | 10 | 27392541 | A | C | -0.0631781 | 9.8923E-08 | 0.0118562 | -5.3286972 | 0.1779 | eqtlPRS |
| rs2292115 | 15 | 78790189 | A | G | -0.154714 | 1.4178E-07 | 0.0293971 | -5.2629001 | 0.03272 | eqtlPRS |
| rs705698 | 12 | 56384687 | T | C | -0.0509268 | 1.8988E-07 | 0.00977672 | -5.2089862 | 0.32 | eqtlPRS |
| rs2434576 | 5 | 138917674 | A | G | 0.0525831 | 2.0904E-07 | 0.0101295 | 5.19108544 | 0.3108 | eqtlPRS |
| rs2239701 | 6 | 32805049 | T | C | 0.0472411 | 2.5346E-07 | 0.0091639 | 5.15513046 | 0.4448 | eqtlPRS |
| rs1055263 | 4 | 164045822 | G | A | -0.0599345 | 2.6374E-07 | 0.011643 | -5.1476853 | 0.2076 | eqtlPRS |
| rs7133567 | 12 | 2993074 | G | A | 0.0638442 | 3.2596E-07 | 0.0124994 | 5.10778117 | 0.1595 | eqtlPRS |
| rs10197246 | 2 | 202204741 | C | T | 0.0517313 | 3.3864E-07 | 0.0101423 | 5.10054919 | 0.2955 | eqtlPRS |
| rs78853063 | 11 | 57250026 | C | T | -0.0910743 | 4.1503E-07 | 0.017992 | -5.0619331 | 0.07362 | eqtlPRS |
| rs77802411 | 15 | 78925414 | T | C | 0.144154 | 5.4322E-07 | 0.0287711 | 5.01037499 | 0.03272 | eqtlPRS |
| rs12327712 | 19 | 17424947 | T | G | 0.0465392 | 5.4544E-07 | 0.00929002 | 5.00959094 | 0.3855 | eqtlPRS |
| rs2106457 | 16 | 23539132 | C | A | 0.0637146 | 6.0073E-07 | 0.0127659 | 4.99099946 | 0.1513 | eqtlPRS |
| rs34547036 | 10 | 105637026 | C | T | 0.0729992 | 6.1121E-07 | 0.014636 | 4.9876469 | 0.09611 | eqtlPRS |
| rs117202602 | 15 | 63744034 | G | A | 0.114277 | 6.5997E-07 | 0.0229806 | 4.97275963 | 0.04192 | eqtlPRS |
| rs72822431 | 2 | 65521816 | T | G | 0.0628331 | 6.8692E-07 | 0.0126551 | 4.96504176 | 0.1483 | eqtlPRS |
| rs72909574 | 11 | 48325454 | G | A | -0.116844 | 7.3882E-07 | 0.0236006 | -4.9508911 | 0.05828 | eqtlPRS |
| rs2294309 | 6 | 27483816 | T | G | -0.0467271 | 7.6357E-07 | 0.0094504 | -4.9444574 | 0.365 | eqtlPRS |
| rs2523690 | 6 | 31420713 | C | T | -0.0572509 | 1.0241E-06 | 0.011715 | -4.886974 | 0.2444 | eqtlPRS |
| rs2197025 | 2 | 214015305 | A | G | -0.0457937 | 1.1151E-06 | 0.00940291 | -4.8701625 | 0.4458 | eqtlPRS |
| rs3824988 | 11 | 57377968 | A | G | 0.0491463 | 1.3646E-06 | 0.010175 | 4.83010319 | 0.2658 | eqtlPRS |
| rs743211 | 14 | 104366179 | T | C | -0.0464107 | 1.4423E-06 | 0.00963063 | -4.8190721 | 0.363 | eqtlPRS |
| rs2072113 | 11 | 61604967 | C | T | -0.067094 | 1.4783E-06 | 0.0139368 | -4.8141611 | 0.1483 | eqtlPRS |
| rs114322499 | 6 | 29801180 | C | T | -0.116803 | 1.5658E-06 | 0.0243205 | -4.8026562 | 0.06748 | eqtlPRS |
| rs113864525 | 9 | 136473053 | C | A | 0.070263 | 1.8093E-06 | 0.014719 | 4.77362593 | 0.091 | eqtlPRS |
| rs174455 | 11 | 61656117 | A | G | -0.04539 | 1.87E-06 | 0.00952172 | -4.7669959 | 0.3885 | eqtlPRS |
| rs2289393 | 2 | 24928579 | T | C | 0.0487881 | 2.084E-06 | 0.0102818 | 4.74509327 | 0.2699 | eqtlPRS |
| rs3025383 | 9 | 136502369 | T | C | -0.056208 | 2.2056E-06 | 0.0118742 | -4.7336242 | 0.181 | eqtlPRS |
| rs11162504 | 1 | 78991197 | A | G | 0.0492613 | 2.2232E-06 | 0.0104102 | 4.73202244 | 0.2485 | eqtlPRS |
| rs978689 | 12 | 46102356 | C | T | 0.0433079 | 2.3405E-06 | 0.00917239 | 4.72155022 | 0.4632 | eqtlPRS |
| rs62482238 | 7 | 100214762 | G | A | -0.0552686 | 2.4088E-06 | 0.0117201 | -4.7157106 | 0.1861 | eqtlPRS |
| rs86715 | 6 | 33480435 | A | G | -0.0653394 | 2.4488E-06 | 0.0138656 | -4.7123384 | 0.135 | eqtlPRS |
| rs4338560 | 11 | 118143368 | C | T | 0.0518212 | 2.7574E-06 | 0.0110537 | 4.68813158 | 0.2076 | eqtlPRS |
| rs7621631 | 3 | 169512145 | C | A | -0.0498031 | 2.8443E-06 | 0.0106377 | -4.6817545 | 0.2464 | eqtlPRS |
| rs2276907 | 4 | 140950160 | G | A | -0.0428764 | 3.1043E-06 | 0.00919346 | -4.6637936 | 0.4254 | eqtlPRS |
| rs7221205 | 17 | 56485799 | G | A | -0.054028 | 3.1634E-06 | 0.0115942 | -4.6599162 | 0.2055 | eqtlPRS |
| rs2273256 | 22 | 50873561 | C | T | 0.049554 | 3.1745E-06 | 0.0106357 | 4.65921378 | 0.2403 | eqtlPRS |
| rs73040690 | 12 | 10791540 | A | C | -0.0935252 | 3.2801E-06 | 0.0201023 | -4.6524627 | 0.06442 | eqtlPRS |
| rs10064596 | 5 | 178983748 | G | A | 0.0438057 | 3.6175E-06 | 0.00945673 | 4.63222488 | 0.3456 | eqtlPRS |
| rs12802931 | 11 | 1236164 | A | G | 0.0637141 | 3.8101E-06 | 0.0137865 | 4.62148479 | 0.182 | eqtlPRS |
| rs78154696 | 5 | 1000156 | G | A | 0.146893 | 4.3871E-06 | 0.0319878 | 4.59215701 | 0.02352 | eqtlPRS |
| rs77045810 | 1 | 168505017 | A | C | -0.0669467 | 4.5792E-06 | 0.014607 | -4.583193 | 0.137 | eqtlPRS |
| rs79778395 | 20 | 62740544 | C | T | 0.0935522 | 4.7325E-06 | 0.0204427 | 4.5763133 | 0.06237 | eqtlPRS |
| rs4140594 | 6 | 36564240 | A | G | 0.217302 | 5.038E-06 | 0.0476205 | 4.56320282 | 0.01125 | eqtlPRS |
| rs74024464 | 15 | 79125155 | G | A | -0.0953825 | 5.2432E-06 | 0.020941 | -4.5548207 | 0.0501 | eqtlPRS |
| rs35561057 | 5 | 1320704 | C | T | -0.1118 | 5.3156E-06 | 0.024561 | -4.5519319 | 0.03579 | eqtlPRS |
| rs7781386 | 7 | 16142180 | C | T | 0.0585068 | 5.3229E-06 | 0.012854 | 4.55164151 | 0.1074 | eqtlPRS |
| rs76994854 | 13 | 111492286 | C | T | 0.151974 | 5.6227E-06 | 0.0334736 | 4.5401152 | 0.03476 | eqtlPRS |
| rs10119 | 19 | 45406673 | G | A | -0.0470401 | 6.0711E-06 | 0.0103981 | -4.523913 | 0.2904 | eqtlPRS |
| rs2160241 | 2 | 202451984 | A | G | -0.0923953 | 6.1463E-06 | 0.0204356 | -4.5212913 | 0.04397 | eqtlPRS |
| rs12461874 | 19 | 17180358 | C | A | -0.0460081 | 6.2372E-06 | 0.0101829 | -4.5181726 | 0.2638 | eqtlPRS |
| rs72836408 | 6 | 26254995 | G | T | -0.155077 | 6.2867E-06 | 0.0343355 | -4.5165208 | 0.0184 | eqtlPRS |
| rs139236208 | 12 | 94880742 | G | A | 0.0743335 | 6.3291E-06 | 0.0164634 | 4.51507587 | 0.08384 | eqtlPRS |
| rs788226 | 10 | 27554985 | C | T | -0.129374 | 6.9259E-06 | 0.0287756 | -4.4959619 | 0.02556 | eqtlPRS |
| rs73032630 | 19 | 39032525 | C | T | 0.0773243 | 7.6774E-06 | 0.0172831 | 4.47398326 | 0.08487 | eqtlPRS |
| rs4939578 | 18 | 46910973 | G | A | 0.0459006 | 7.8186E-06 | 0.0102684 | 4.47008297 | 0.2781 | eqtlPRS |
| rs3917290 | 2 | 102782079 | C | T | 0.0417462 | 7.8305E-06 | 0.00933969 | 4.46976292 | 0.3569 | eqtlPRS |
| rs77003790 | 15 | 91397771 | C | T | 0.0785509 | 8.2508E-06 | 0.017618 | 4.45855943 | 0.08691 | eqtlPRS |
| rs1025526 | 1 | 231750641 | T | C | 0.0429872 | 8.7609E-06 | 0.00966941 | 4.44569007 | 0.3262 | eqtlPRS |
| rs71645854 | 1 | 47654458 | T | G | -0.0654 | 8.8448E-06 | 0.0147177 | -4.4436291 | 0.1125 | eqtlPRS |
| rs13344413 | 19 | 39143289 | G | A | 0.0408045 | 8.9131E-06 | 0.0091861 | 4.441983 | 0.411 | eqtlPRS |
| rs1250532 | 10 | 81017157 | C | T | 0.0443096 | 8.9237E-06 | 0.00997574 | 4.44173565 | 0.3037 | eqtlPRS |
| rs113659074 | 4 | 100252308 | G | T | -0.0803618 | 8.9957E-06 | 0.0180995 | -4.4400011 | 0.07771 | eqtlPRS |
| rs12476193 | 2 | 18097936 | G | A | 0.041156 | 9.1131E-06 | 0.0092752 | 4.4372089 | 0.4458 | eqtlPRS |
| rs7631551 | 3 | 46186310 | C | A | -0.071888 | 9.8807E-06 | 0.0162651 | -4.4197699 | 0.09918 | eqtlPRS |
| rs75484351 | 15 | 78594423 | C | T | 0.0881091 | 1.0061E-05 | 0.0199528 | 4.41587647 | 0.05317 | eqtlPRS |
| rs7674827 | 4 | 113033568 | G | A | 0.0420532 | 1.0118E-05 | 0.00952585 | 4.41464016 | 0.3415 | eqtlPRS |
| rs2178313 | 3 | 121346600 | C | T | 0.0868619 | 1.0278E-05 | 0.019691 | 4.41124879 | 0.06544 | eqtlPRS |
| rs653178 | 12 | 112007756 | T | C | -0.0405156 | 1.0334E-05 | 0.00918705 | -4.4100772 | 0.4683 | eqtlPRS |
| rs4970484 | 1 | 27092312 | G | A | -0.062919 | 1.0575E-05 | 0.0142833 | -4.4050745 | 0.1728 | eqtlPRS |
| rs745215 | 10 | 91429271 | C | T | -0.040311 | 1.1396E-05 | 0.00918489 | -4.3888386 | 0.4622 | eqtlPRS |
| rs17599629 | 1 | 150658287 | A | G | 0.0483572 | 1.1728E-05 | 0.0110339 | 4.38260271 | 0.1984 | eqtlPRS |
| rs139097404 | 15 | 43933941 | T | C | -0.141153 | 1.2159E-05 | 0.0322656 | -4.3747211 | 0.02147 | eqtlPRS |
| rs2728125 | 4 | 89001893 | A | G | -0.0684567 | 1.2656E-05 | 0.0156796 | -4.3659723 | 0.0726 | eqtlPRS |
| rs62332583 | 5 | 1286037 | C | T | 0.178179 | 1.2774E-05 | 0.0408298 | 4.36394496 | 0.01329 | eqtlPRS |
| rs11162417 | 1 | 78619038 | T | C | -0.0412603 | 1.2892E-05 | 0.00945916 | -4.3619412 | 0.4284 | eqtlPRS |
| rs112761610 | 11 | 661697 | C | T | 0.129176 | 1.3129E-05 | 0.0296415 | 4.3579441 | 0.02863 | eqtlPRS |
| rs2783386 | 1 | 179236579 | G | A | -0.055803 | 1.3229E-05 | 0.0128098 | -4.3562741 | 0.1769 | eqtlPRS |
| rs75100087 | 20 | 62636139 | C | T | -0.0574404 | 1.3252E-05 | 0.0131868 | -4.3559014 | 0.1299 | eqtlPRS |
| rs113016973 | 9 | 136113324 | C | T | 0.103759 | 1.3746E-05 | 0.0238643 | 4.34787528 | 0.01943 | eqtlPRS |
| rs1541965 | 15 | 79205126 | G | A | 0.0903501 | 1.439E-05 | 0.0208285 | 4.33781117 | 0.04601 | eqtlPRS |
| rs35762452 | 19 | 5252175 | G | A | 0.0757974 | 1.5002E-05 | 0.0175106 | 4.32865807 | 0.07362 | eqtlPRS |
| rs58280122 | 2 | 234260231 | C | A | -0.0697839 | 1.5398E-05 | 0.0161428 | -4.3229118 | 0.08793 | eqtlPRS |
| rs699245 | 1 | 183521283 | C | T | 0.0393664 | 1.6167E-05 | 0.00912915 | 4.31216488 | 0.4673 | eqtlPRS |
| rs11775733 | 8 | 27641026 | C | T | 0.146824 | 1.6169E-05 | 0.034049 | 4.31213839 | 0.01534 | eqtlPRS |
| rs1054866 | 20 | 61571208 | A | C | -0.0426353 | 1.6273E-05 | 0.00989054 | -4.3107151 | 0.2873 | eqtlPRS |
| rs9328534 | 9 | 134874805 | T | C | 0.0419043 | 1.6513E-05 | 0.00972827 | 4.30747707 | 0.319 | eqtlPRS |
| rs74829122 | 5 | 1241565 | A | G | -0.0946955 | 1.6846E-05 | 0.0220066 | -4.30305 | 0.07566 | eqtlPRS |
| rs4737074 | 8 | 42663781 | C | T | -0.0474014 | 1.7196E-05 | 0.0110274 | -4.298511 | 0.2311 | eqtlPRS |
| rs4756168 | 11 | 34764919 | C | A | 0.0529716 | 1.722E-05 | 0.0123242 | 4.29817757 | 0.1626 | eqtlPRS |
| rs72793280 | 5 | 131562900 | C | T | 0.0397432 | 1.726E-05 | 0.0092476 | 4.29767724 | 0.4202 | eqtlPRS |
| rs2302212 | 4 | 17818885 | C | T | 0.0395597 | 1.7553E-05 | 0.00921293 | 4.29393255 | 0.4008 | eqtlPRS |
| rs72951731 | 6 | 117308758 | T | C | 0.0927781 | 1.7627E-05 | 0.0216115 | 4.29299678 | 0.05215 | eqtlPRS |
| rs10868787 | 9 | 91101651 | A | G | -0.0580749 | 1.8088E-05 | 0.0135459 | -4.2872677 | 0.1268 | eqtlPRS |
| rs1368442 | 19 | 39835169 | T | G | 0.0399938 | 1.8147E-05 | 0.00933007 | 4.28654876 | 0.4785 | eqtlPRS |
| rs62396224 | 6 | 26292926 | A | G | -0.0922849 | 1.8234E-05 | 0.0215343 | -4.2854841 | 0.0593 | eqtlPRS |
| rs77235729 | 14 | 101535711 | G | T | 0.128487 | 1.8746E-05 | 0.0300252 | 4.27930538 | 0.03067 | eqtlPRS |
| rs2286604 | 12 | 2926543 | A | G | 0.051629 | 1.8758E-05 | 0.0120651 | 4.279202 | 0.1667 | eqtlPRS |
| rs8140581 | 22 | 19294533 | C | T | 0.0506791 | 1.8779E-05 | 0.0118439 | 4.27891995 | 0.18 | eqtlPRS |
| rs1208083 | 2 | 202532447 | G | A | -0.0477106 | 1.921E-05 | 0.0111633 | -4.2738796 | 0.1973 | eqtlPRS |
| rs12378118 | 9 | 6033278 | G | A | 0.0486146 | 1.9563E-05 | 0.0113856 | 4.26983207 | 0.1646 | eqtlPRS |
| rs6546541 | 2 | 69908621 | C | T | 0.0390221 | 1.9895E-05 | 0.0091471 | 4.26606247 | 0.4888 | eqtlPRS |
| rs932826 | 20 | 62380516 | A | C | -0.0400012 | 2.0225E-05 | 0.00938467 | -4.2623981 | 0.407 | eqtlPRS |
| rs2519150 | 9 | 136491172 | T | C | -0.0388444 | 2.1205E-05 | 0.00913599 | -4.2517998 | 0.4939 | eqtlPRS |
| rs115402147 | 1 | 44516962 | G | A | -0.0981229 | 2.1318E-05 | 0.0230844 | -4.2506151 | 0.03579 | eqtlPRS |
| rs8079078 | 17 | 65821166 | A | C | 0.0418355 | 2.1776E-05 | 0.00985326 | 4.24585366 | 0.319 | eqtlPRS |
| rs4975662 | 5 | 1090368 | T | C | 0.0389182 | 2.1815E-05 | 0.00916702 | 4.24545818 | 0.4571 | eqtlPRS |
| rs12419373 | 11 | 63691892 | A | G | -0.0737259 | 2.2181E-05 | 0.0173811 | -4.2417281 | 0.07975 | eqtlPRS |
| rs57570776 | 15 | 101072736 | C | T | -0.0901066 | 2.2729E-05 | 0.0212704 | -4.2362438 | 0.04703 | eqtlPRS |
| rs4924457 | 15 | 40653435 | A | G | -0.0409424 | 2.3204E-05 | 0.0096754 | -4.2315977 | 0.3701 | eqtlPRS |
| rs3091309 | 3 | 46303184 | G | A | 0.0489757 | 2.3374E-05 | 0.0115783 | 4.22995604 | 0.1697 | eqtlPRS |
| rs79004324 | 20 | 4639380 | G | A | -0.0476644 | 2.3556E-05 | 0.011273 | -4.2281913 | 0.2188 | eqtlPRS |
| rs75099792 | 15 | 78956846 | C | T | -0.179224 | 2.3898E-05 | 0.0424203 | -4.2249583 | 0.01738 | eqtlPRS |
| rs1267693 | 3 | 120199467 | C | T | -0.0520597 | 2.5181E-05 | 0.0123564 | -4.213177 | 0.1605 | eqtlPRS |
| rs114806656 | 5 | 138337651 | G | A | -0.134009 | 2.5274E-05 | 0.0318134 | -4.2123445 | 0.02658 | eqtlPRS |
| rs329122 | 5 | 133864599 | G | A | -0.0386908 | 2.5287E-05 | 0.00918535 | -4.2122293 | 0.4294 | eqtlPRS |
| rs116082446 | 11 | 73495606 | C | T | 0.116618 | 2.58E-05 | 0.0277155 | 4.20768162 | 0.02863 | eqtlPRS |
| rs848629 | 2 | 36798415 | C | T | 0.0395591 | 2.5895E-05 | 0.00940349 | 4.20685299 | 0.3998 | eqtlPRS |
| rs6724281 | 2 | 88529531 | G | T | 0.0418945 | 2.6301E-05 | 0.00996697 | 4.20333361 | 0.2802 | eqtlPRS |
| rs8112072 | 19 | 51741143 | A | G | 0.118455 | 2.6965E-05 | 0.028219 | 4.19770367 | 0.02761 | eqtlPRS |
| rs4802509 | 19 | 49307617 | A | G | 0.0987528 | 2.7191E-05 | 0.0235361 | 4.19580134 | 0.03885 | eqtlPRS |
| rs10902436 | 12 | 132178537 | A | G | -0.0406914 | 0.00002745 | 0.0097031 | -4.1936495 | 0.4448 | eqtlPRS |
| rs61743921 | 1 | 201184878 | C | A | -0.0803939 | 2.8238E-05 | 0.0191998 | -4.1872259 | 0.04908 | eqtlPRS |
| rs11023094 | 11 | 2527361 | G | A | 0.0516023 | 2.9074E-05 | 0.0123433 | 4.1805919 | 0.2014 | eqtlPRS |
| rs62109707 | 19 | 36464626 | C | T | -0.127511 | 2.9213E-05 | 0.0305084 | -4.1795374 | 0.03885 | eqtlPRS |
| rs2527895 | 7 | 99538594 | G | A | 0.0383278 | 2.9239E-05 | 0.00917083 | 4.17931638 | 0.453 | eqtlPRS |
| rs118162691 | 8 | 21767809 | C | A | -0.100975 | 2.9525E-05 | 0.0241734 | -4.177112 | 0.0409 | eqtlPRS |
| rs17389502 | 1 | 51468754 | G | A | 0.0426866 | 3.0374E-05 | 0.010235 | 4.17064973 | 0.271 | eqtlPRS |
| rs2208590 | 20 | 47411456 | C | T | -0.0386632 | 3.0501E-05 | 0.00927243 | -4.1696945 | 0.3661 | eqtlPRS |
| rs11670094 | 19 | 48681930 | T | C | 0.0894151 | 3.0774E-05 | 0.0214545 | 4.1676618 | 0.0726 | eqtlPRS |
| rs6059298 | 20 | 31984124 | G | A | -0.0511638 | 3.11E-05 | 0.0122835 | -4.1652461 | 0.1544 | eqtlPRS |
| rs73161745 | 7 | 99893765 | C | T | 0.0427836 | 3.1532E-05 | 0.0102793 | 4.16211221 | 0.2863 | eqtlPRS |
| rs76185923 | 19 | 7076791 | C | T | -0.121816 | 3.2484E-05 | 0.0293156 | -4.1553303 | 0.03067 | eqtlPRS |
| rs1410138 | 1 | 230756497 | T | C | -0.121256 | 3.3023E-05 | 0.0292074 | -4.1515506 | 0.03783 | eqtlPRS |
| rs910928 | 20 | 61720769 | C | T | -0.0382785 | 3.3593E-05 | 0.009229 | -4.1476325 | 0.4376 | eqtlPRS |
| rs6824221 | 4 | 174842251 | C | T | -0.040356 | 3.3614E-05 | 0.00973021 | -4.1474953 | 0.3252 | eqtlPRS |
| rs17103378 | 12 | 67503844 | T | C | -0.0555991 | 3.3732E-05 | 0.0134081 | -4.14668 | 0.1309 | eqtlPRS |
| rs146305655 | 6 | 32268953 | G | A | -0.116513 | 3.4601E-05 | 0.0281375 | -4.1408441 | 0.02965 | eqtlPRS |
| rs1860229 | 5 | 142078069 | G | A | -0.0901564 | 3.4854E-05 | 0.0217812 | -4.1391843 | 0.04397 | eqtlPRS |
| rs62290089 | 3 | 184399214 | C | T | -0.0955936 | 3.5049E-05 | 0.0231019 | -4.1379107 | 0.05419 | eqtlPRS |
| rs238418 | 19 | 45855262 | G | T | 0.038879 | 3.5086E-05 | 0.00939638 | 4.13765727 | 0.3783 | eqtlPRS |
| rs2682675 | 12 | 31533318 | C | T | 0.104 | 0.00003549 | 0.0251511 | 4.13500801 | 0.03783 | eqtlPRS |
| rs871524 | 1 | 38411445 | G | A | -0.0420301 | 3.6512E-05 | 0.0101804 | -4.1285313 | 0.32 | eqtlPRS |
| rs62395827 | 6 | 31786730 | C | T | -0.065754 | 3.6653E-05 | 0.0159302 | -4.1276318 | 0.08078 | eqtlPRS |
| rs4845553 | 1 | 153480173 | A | G | -0.055947 | 3.7659E-05 | 0.0135748 | -4.1213867 | 0.1339 | eqtlPRS |
| rs73544230 | 9 | 123145634 | T | C | -0.0466695 | 3.79E-05 | 0.0113278 | -4.1199085 | 0.2025 | eqtlPRS |
| rs12437675 | 15 | 84185007 | C | T | 0.0719193 | 3.9225E-05 | 0.0174901 | 4.1120005 | 0.07362 | eqtlPRS |
| rs2604909 | 19 | 41297615 | C | T | 0.0453975 | 3.983E-05 | 0.0110497 | 4.10848258 | 0.2209 | eqtlPRS |
| rs114564790 | 6 | 26589046 | G | A | 0.0993555 | 3.9831E-05 | 0.0241832 | 4.10845132 | 0.0501 | eqtlPRS |
| rs56112907 | 1 | 155070862 | A | C | -0.0374839 | 4.0939E-05 | 0.00913769 | -4.1021199 | 0.4918 | eqtlPRS |
| rs78035834 | 7 | 63592275 | T | C | -0.112269 | 4.0963E-05 | 0.0273695 | -4.1019748 | 0.02761 | eqtlPRS |
| rs10884942 | 10 | 112016926 | A | G | -0.0392198 | 4.1922E-05 | 0.00957369 | -4.0966231 | 0.3558 | eqtlPRS |
| rs7200434 | 16 | 686398 | A | G | 0.154566 | 4.4129E-05 | 0.0378399 | 4.08473595 | 0.01636 | eqtlPRS |
| rs114269138 | 15 | 84116815 | C | A | -0.0525003 | 4.4841E-05 | 0.0128646 | -4.0809897 | 0.1431 | eqtlPRS |
| rs77881327 | 6 | 125600052 | G | A | 0.106847 | 4.5817E-05 | 0.0262136 | 4.07601398 | 0.02761 | eqtlPRS |
| rs67313812 | 5 | 40967444 | G | A | -0.0435983 | 4.6455E-05 | 0.0107048 | -4.0727804 | 0.2239 | eqtlPRS |
| rs62132157 | 19 | 4133055 | G | T | -0.0607836 | 4.6533E-05 | 0.0149258 | -4.0723847 | 0.1258 | eqtlPRS |
| rs55731474 | 14 | 103878481 | G | A | 0.0387202 | 4.6768E-05 | 0.00951072 | 4.07121648 | 0.3517 | eqtlPRS |
| rs11243104 | 6 | 6566619 | C | A | 0.0379845 | 4.6792E-05 | 0.00933029 | 4.07109533 | 0.4049 | eqtlPRS |
| rs11722701 | 4 | 76171211 | A | G | -0.0413841 | 4.784E-05 | 0.0101782 | -4.0659547 | 0.2986 | eqtlPRS |
| rs11170164 | 12 | 52913668 | C | T | 0.068386 | 4.8283E-05 | 0.0168282 | 4.0637739 | 0.06544 | eqtlPRS |
| rs6911745 | 6 | 33677661 | G | A | -0.123696 | 4.9062E-05 | 0.0304666 | -4.0600526 | 0.03681 | eqtlPRS |
| rs3847711 | 12 | 58834035 | A | G | 0.0557415 | 5.0437E-05 | 0.0137511 | 4.05360298 | 0.1391 | eqtlPRS |
| rs13268919 | 8 | 23096594 | G | A | -0.0383634 | 5.1218E-05 | 0.00947244 | -4.0500019 | 0.3742 | eqtlPRS |
| rs144704378 | 5 | 1259489 | C | T | 0.0921028 | 5.1939E-05 | 0.0227598 | 4.04673152 | 0.03681 | eqtlPRS |
| rs1402001 | 3 | 183642751 | A | G | 0.036806 | 5.2543E-05 | 0.00910135 | 4.04401545 | 0.4755 | eqtlPRS |
| rs1456244 | 11 | 93171107 | T | C | -0.0709591 | 5.4133E-05 | 0.0175771 | -4.0370198 | 0.07464 | eqtlPRS |
| rs4982832 | 14 | 24461235 | C | T | 0.0652463 | 5.452E-05 | 0.0161687 | 4.03534607 | 0.1166 | eqtlPRS |
| rs1998424 | 14 | 24643782 | T | C | 0.0378248 | 5.5797E-05 | 0.00938599 | 4.02992119 | 0.4141 | eqtlPRS |
| rs77322113 | 5 | 76192017 | C | A | 0.087139 | 5.5888E-05 | 0.0216251 | 4.0295305 | 0.04294 | eqtlPRS |
| rs11871280 | 17 | 45559843 | G | A | -0.140539 | 5.5898E-05 | 0.0348776 | -4.0294917 | 0.02249 | eqtlPRS |
| rs3852860 | 19 | 45382966 | C | T | 0.0377777 | 5.6868E-05 | 0.00938472 | 4.02544775 | 0.4141 | eqtlPRS |
| rs115974489 | 5 | 1214606 | C | T | 0.126321 | 5.7075E-05 | 0.0313872 | 4.02460239 | 0.02454 | eqtlPRS |
| rs57339473 | 19 | 41452921 | C | T | -0.0570477 | 5.7127E-05 | 0.0141755 | -4.0243871 | 0.1411 | eqtlPRS |
| rs7785866 | 7 | 36792564 | G | A | 0.0471327 | 5.7137E-05 | 0.0117119 | 4.02434276 | 0.2198 | eqtlPRS |
| rs12571579 | 10 | 27455153 | A | G | 0.0807198 | 6.011E-05 | 0.0201177 | 4.01237716 | 0.06442 | eqtlPRS |
| rs10835995 | 11 | 4413026 | T | C | 0.0443481 | 6.0331E-05 | 0.0110552 | 4.01151494 | 0.2413 | eqtlPRS |
| rs12081674 | 1 | 90337025 | A | G | 0.0477332 | 6.0658E-05 | 0.0119028 | 4.01024969 | 0.1718 | eqtlPRS |
| rs568878 | 11 | 95578694 | G | T | -0.0367807 | 6.1724E-05 | 0.00918113 | -4.0061191 |  | eqtlPRS |
| rs198470 | 11 | 61528306 | C | A | 0.0386596 | 6.1811E-05 | 0.00965092 | 4.00579427 | 0.3722 | eqtlPRS |
| rs405509 | 19 | 45408836 | G | T | -0.0366596 | 6.2841E-05 | 0.00916059 | -4.001882 | 0.4847 | eqtlPRS |
| rs60816576 | 4 | 89097087 | G | T | 0.0497248 | 6.3102E-05 | 0.0124284 | 4.00090116 | 0.1861 | eqtlPRS |
| rs404613 | 20 | 62173925 | T | G | 0.0798293 | 6.3628E-05 | 0.0199626 | 3.99894302 | 0.06646 | eqtlPRS |
| rs17506431 | 8 | 38582476 | C | T | 0.0483274 | 6.3688E-05 | 0.0120857 | 3.99872577 | 0.1994 | eqtlPRS |
| rs72745113 | 15 | 79008966 | G | A | 0.122895 | 6.54E-05 | 0.0307821 | 3.99241767 | 0.03579 | eqtlPRS |
| rs1279293 | 11 | 74050173 | G | A | -0.0393168 | 6.6245E-05 | 0.00985537 | -3.9893784 | 0.318 | eqtlPRS |
| rs1079145 | 6 | 167360724 | G | A | 0.0677549 | 6.6444E-05 | 0.0169868 | 3.98867945 | 0.09816 | eqtlPRS |
| rs11064584 | 12 | 996671 | G | A | 0.0423164 | 6.6448E-05 | 0.0106092 | 3.98865136 | 0.2474 | eqtlPRS |
| rs4671067 | 2 | 63798167 | T | C | -0.0400824 | 6.7209E-05 | 0.0100559 | -3.9859585 | 0.2945 | eqtlPRS |
| rs2967 | 6 | 33689534 | G | A | 0.0404203 | 6.7582E-05 | 0.010144 | 3.98465103 | 0.2618 | eqtlPRS |
| rs148513508 | 4 | 100403028 | A | G | 0.108834 | 6.8348E-05 | 0.0273318 | 3.98195509 | 0.03374 | eqtlPRS |
| rs155428 | 5 | 72109700 | C | T | -0.0390756 | 6.8696E-05 | 0.00981613 | -3.9807541 | 0.3231 | eqtlPRS |
| rs76022354 | 10 | 94306385 | T | C | -0.0909526 | 6.8729E-05 | 0.0228487 | -3.9806466 | 0.03374 | eqtlPRS |
| rs28887188 | 2 | 231425650 | G | A | -0.0594458 | 6.901E-05 | 0.0149374 | -3.9796618 | 0.1115 | eqtlPRS |
| rs113794582 | 17 | 20340342 | T | C | 0.0864534 | 7.1851E-05 | 0.0217763 | 3.97006838 | 0.04703 | eqtlPRS |
| rs4897759 | 10 | 133509442 | A | G | 0.0454561 | 7.2095E-05 | 0.011452 | 3.96927174 | 0.2096 | eqtlPRS |
| rs8034099 | 15 | 80725930 | G | A | -0.0479775 | 7.2259E-05 | 0.0120889 | -3.9687234 | 0.1697 | eqtlPRS |
| rs28580505 | 5 | 96101220 | A | G | -0.0388563 | 7.3122E-05 | 0.00979762 | -3.9658917 | 0.3354 | eqtlPRS |
| rs5753170 | 22 | 30896093 | G | A | 0.0743245 | 7.3559E-05 | 0.0187477 | 3.96445964 | 0.06953 | eqtlPRS |
| rs34818580 | 19 | 45894138 | G | A | 0.0877056 | 7.5073E-05 | 0.0221501 | 3.95960289 | 0.05726 | eqtlPRS |
| rs113447691 | 11 | 119848376 | C | T | -0.0670066 | 7.5373E-05 | 0.0169266 | -3.9586568 | 0.08078 | eqtlPRS |
| rs139829601 | 2 | 55051858 | T | C | 0.155553 | 7.5526E-05 | 0.0392991 | 3.95818225 | 0.01738 | eqtlPRS |
| rs4821526 | 22 | 37184203 | T | G | 0.0396538 | 7.563E-05 | 0.0100191 | 3.95782056 | 0.3354 | eqtlPRS |
| rs59036121 | 12 | 133416125 | C | T | 0.0394188 | 7.6919E-05 | 0.00996985 | 3.95380071 | 0.2761 | eqtlPRS |
| rs176415 | 2 | 32649778 | G | A | 0.0381724 | 7.7663E-05 | 0.00966023 | 3.95150012 | 0.3211 | eqtlPRS |
| rs2127900 | 15 | 79007561 | A | G | -0.11207 | 7.7962E-05 | 0.0283679 | -3.950592 | 0.03579 | eqtlPRS |
| rs4801925 | 19 | 53165057 | A | C | 0.0391125 | 7.8084E-05 | 0.00990139 | 3.95020295 | 0.3517 | eqtlPRS |
| rs2075061 | 17 | 36934124 | T | C | -0.0391995 | 7.8811E-05 | 0.00992898 | -3.9479886 | 0.4356 | eqtlPRS |
| rs9620235 | 22 | 23628161 | A | G | 0.0413251 | 7.8921E-05 | 0.0104683 | 3.94764193 | 0.2812 | eqtlPRS |
| rs873972 | 7 | 75444549 | C | T | -0.0362166 | 7.8942E-05 | 0.00917436 | -3.9475887 | 0.4908 | eqtlPRS |
| rs142310629 | 4 | 57358996 | C | T | -0.167816 | 7.9826E-05 | 0.0425399 | -3.9449082 | 0.01534 | eqtlPRS |
| rs11079346 | 17 | 56383744 | T | C | 0.0393092 | 8.1631E-05 | 0.00997806 | 3.9395634 | 0.3221 | eqtlPRS |
| rs55699579 | 7 | 151131929 | T | C | -0.0568689 | 8.2942E-05 | 0.0144494 | -3.9357274 | 0.1176 | eqtlPRS |
| rs10745340 | 1 | 114436970 | T | C | -0.0373334 | 8.3108E-05 | 0.0094869 | -3.9352581 | 0.3783 | eqtlPRS |
| rs7438777 | 4 | 151083488 | T | G | -0.042762 | 8.3309E-05 | 0.010868 | -3.9346706 | 0.2249 | eqtlPRS |
| rs79183029 | 19 | 41298228 | C | T | 0.10803 | 8.3645E-05 | 0.0274625 | 3.93372781 | 0.0184 | eqtlPRS |
| rs3862619 | 11 | 125610338 | G | T | 0.0363756 | 8.3661E-05 | 0.00924726 | 3.93366251 | 0.411 | eqtlPRS |
| rs601310 | 11 | 65040436 | T | C | -0.0424678 | 8.4173E-05 | 0.0108 | -3.9322037 | 0.2515 | eqtlPRS |
| rs145713288 | 19 | 52052302 | T | C | 0.135978 | 8.5335E-05 | 0.0346098 | 3.92888719 | 0.01636 | eqtlPRS |
| rs61397463 | 7 | 43533850 | T | C | 0.0857617 | 8.6944E-05 | 0.0218534 | 3.92440993 | 0.04499 | eqtlPRS |
| rs2888977 | 12 | 109270863 | C | T | -0.0359843 | 8.9904E-05 | 0.00918824 | -3.9163431 | 0.4734 | eqtlPRS |
| rs3110098 | 17 | 28607211 | C | T | 0.131649 | 9.0426E-05 | 0.0336272 | 3.91495575 | 0.03783 | eqtlPRS |
| rs75394182 | 12 | 3849973 | C | T | -0.0751518 | 9.1066E-05 | 0.0192045 | -3.9132391 | 0.05726 | eqtlPRS |
| rs4542208 | 1 | 223633739 | G | A | 0.0508557 | 9.1938E-05 | 0.0130035 | 3.91092398 | 0.1646 | eqtlPRS |
| rs141950045 | 19 | 42146707 | C | T | 0.145046 | 9.3744E-05 | 0.037132 | 3.90622644 | 0.01431 | eqtlPRS |
| rs3219005 | 3 | 9794520 | A | C | -0.0938814 | 9.3899E-05 | 0.0240362 | -3.9058337 | 0.03885 | eqtlPRS |
| rs137864244 | 11 | 57209884 | A | C | 0.133482 | 9.3929E-05 | 0.0341756 | 3.90576903 | 0.03272 | eqtlPRS |
| rs72636505 | 1 | 16405951 | T | C | -0.0478794 | 9.469E-05 | 0.0122648 | -3.903806 | 0.2485 | eqtlPRS |
| rs139060080 | 11 | 65301603 | G | A | 0.0894088 | 9.6347E-05 | 0.0229276 | 3.89961444 | 0.03579 | eqtlPRS |
| rs149485511 | 8 | 22242185 | G | T | -0.143254 | 9.7E-05 | 0.0367508 | -3.8979832 | 0.01738 | eqtlPRS |
| rs149477354 | 15 | 68285163 | C | T | -0.12606 | 9.7184E-05 | 0.0323437 | -3.8975133 | 0.02761 | eqtlPRS |
| rs33954292 | 12 | 962461 | G | T | -0.0466336 | 9.7638E-05 | 0.0119684 | -3.8963938 | 0.184 | eqtlPRS |
| rs7042263 | 9 | 127039017 | T | C | -0.0360733 | 9.8869E-05 | 0.00926536 | -3.8933511 | 0.4356 | eqtlPRS |
| rs9737518 | 11 | 60773327 | C | T | -0.0609228 | 0.0001001 | 0.01566 | -3.8903448 | 0.1094 | eqtlPRS |
| rs2505298 | 10 | 27691009 | A | G | 0.0367624 | 0.00010312 | 0.00946722 | 3.88312514 | 0.3436 | eqtlPRS |
| rs12802244 | 11 | 47932666 | G | A | -0.0376109 | 0.00010468 | 0.00969481 | -3.8794881 | 0.3865 | eqtlPRS |
| rs4965843 | 15 | 101933226 | A | G | 0.0499939 | 0.00010561 | 0.0128939 | 3.87732959 | 0.1431 | eqtlPRS |
| rs7219555 | 17 | 59740122 | A | C | 0.0385102 | 0.00010656 | 0.00993773 | 3.87515056 | 0.2945 | eqtlPRS |
| rs10786615 | 10 | 102777461 | C | T | -0.037177 | 0.0001084 | 0.00960402 | -3.8709832 | 0.4305 | eqtlPRS |
| rs17145750 | 7 | 73026378 | C | T | 0.0490658 | 0.00010937 | 0.0126824 | 3.86881032 | 0.1472 | eqtlPRS |
| rs11589855 | 1 | 154755843 | C | T | -0.0366549 | 0.00010982 | 0.00947691 | -3.8678113 | 0.4581 | eqtlPRS |
| rs12218075 | 10 | 101900190 | T | C | 0.0609129 | 0.00011107 | 0.01576 | 3.86503173 | 0.07873 | eqtlPRS |
| rs1565773 | 12 | 104156715 | G | A | -0.0399653 | 0.00011111 | 0.0103405 | -3.8649292 | 0.2689 | eqtlPRS |
| rs7601 | 15 | 91509592 | T | C | -0.0389605 | 0.00011216 | 0.0100864 | -3.8626765 | 0.2986 | eqtlPRS |
| rs73890196 | 3 | 194383168 | T | C | 0.0751103 | 0.00011358 | 0.0194608 | 3.85956898 | 0.06339 | eqtlPRS |
| rs6090249 | 20 | 61746786 | G | A | -0.0768441 | 0.00011393 | 0.0199138 | -3.8588366 | 0.05419 | eqtlPRS |
| rs2259797 | 20 | 62272248 | T | C | -0.0655775 | 0.00011407 | 0.0169955 | -3.8585214 | 0.1135 | eqtlPRS |
| rs12624447 | 20 | 62031046 | A | G | -0.0370772 | 0.00011448 | 0.00961134 | -3.8576515 | 0.4366 | eqtlPRS |
| rs79127996 | 20 | 2885482 | G | A | -0.0607784 | 0.00011448 | 0.0157553 | -3.8576479 | 0.1135 | eqtlPRS |
| rs55715017 | 19 | 44549793 | T | C | -0.0794441 | 0.00011469 | 0.0205963 | -3.8572025 | 0.06033 | eqtlPRS |
| rs10015976 | 4 | 56030293 | A | G | -0.0413403 | 0.00011576 | 0.010724 | -3.8549329 | 0.3701 | eqtlPRS |
| rs140279297 | 17 | 15293114 | C | T | 0.130938 | 0.00011607 | 0.0339722 | 3.85426908 | 0.01738 | eqtlPRS |
| rs76182238 | 6 | 150750625 | C | T | -0.0852022 | 0.00011637 | 0.0221095 | -3.8536466 | 0.04499 | eqtlPRS |
| rs34407109 | 7 | 43639000 | T | C | 0.0396582 | 0.000117 | 0.0102946 | 3.85233035 | 0.2894 | eqtlPRS |
| rs2153552 | 14 | 50819369 | A | G | 0.0368194 | 0.00011765 | 0.00956105 | 3.85097871 | 0.3814 | eqtlPRS |
| rs4583740 | 4 | 8299162 | T | C | -0.0425457 | 0.00012109 | 0.0110684 | -3.8438889 | 0.228 | eqtlPRS |
| rs72802985 | 5 | 140350139 | G | A | -0.0753334 | 0.00012412 | 0.0196291 | -3.8378428 | 0.08589 | eqtlPRS |
| rs56071450 | 15 | 52591352 | A | G | 0.0408789 | 0.00012625 | 0.0106632 | 3.83364281 | 0.2648 | eqtlPRS |
| rs112370969 | 9 | 93977315 | G | A | -0.147223 | 0.00012716 | 0.0384204 | -3.8318966 | 0.02045 | eqtlPRS |
| rs12403454 | 1 | 45000436 | G | A | -0.04423 | 0.00012795 | 0.0115472 | -3.8303658 | 0.226 | eqtlPRS |
| rs56281723 | 14 | 31621633 | T | C | 0.0353041 | 0.00012817 | 0.00921792 | 3.82994211 | 0.4908 | eqtlPRS |
| rs921934 | 19 | 50667450 | G | T | 0.0695506 | 0.00012863 | 0.0181638 | 3.82907762 | 0.07873 | eqtlPRS |
| rs7599772 | 2 | 47521570 | A | C | 0.0357013 | 0.00013093 | 0.00933438 | 3.82471037 | 0.4601 | eqtlPRS |
| rs4406174 | 5 | 1519079 | C | T | -0.0584376 | 0.00013103 | 0.0152797 | -3.8245254 | 0.1074 | eqtlPRS |
| rs2509751 | 18 | 13085129 | G | A | 0.0354517 | 0.00013133 | 0.00927095 | 3.82395547 | 0.411 | eqtlPRS |
| rs445754 | 14 | 23863802 | G | T | 0.0417852 | 0.00013172 | 0.0109293 | 3.82322747 | 0.2229 | eqtlPRS |
| rs1729412 | 2 | 201083598 | C | T | 0.0351359 | 0.00013287 | 0.00919528 | 3.82107995 | 0.4213 | eqtlPRS |
| rs151183751 | 20 | 14187792 | C | T | 0.109568 | 0.00013366 | 0.0286856 | 3.81961681 | 0.01431 | eqtlPRS |
| rs56216220 | 17 | 28209182 | G | A | 0.0348853 | 0.00013389 | 0.00913424 | 3.81917926 | 0.4622 | eqtlPRS |
| rs10266703 | 7 | 1918941 | C | A | 0.035212 | 0.00013393 | 0.00921991 | 3.81912622 | 0.4366 | eqtlPRS |
| rs79304413 | 22 | 46882317 | C | T | -0.0706538 | 0.00013568 | 0.0185156 | -3.8159066 | 0.08282 | eqtlPRS |
| rs60609289 | 10 | 71966417 | C | T | -0.0775099 | 0.00013755 | 0.0203303 | -3.812531 | 0.06135 | eqtlPRS |
| rs2473336 | 1 | 15711487 | T | C | 0.0395775 | 0.00013989 | 0.0103922 | 3.80838514 | 0.2587 | eqtlPRS |
| rs4900557 | 14 | 103252262 | G | A | -0.0379367 | 0.00014282 | 0.00997487 | -3.8032275 | 0.2965 | eqtlPRS |
| rs73008217 | 6 | 144370331 | G | A | -0.0669007 | 0.00014285 | 0.0175907 | -3.8031858 | 0.0726 | eqtlPRS |
| rs9379818 | 6 | 26023206 | G | T | -0.0349379 | 0.00014371 | 0.0091901 | -3.8016888 | 0.4233 | eqtlPRS |
| rs13204527 | 6 | 133035855 | C | T | -0.0476644 | 0.00014383 | 0.0125384 | -3.8014739 | 0.1912 | eqtlPRS |
| rs1091917 | 3 | 9763748 | C | T | 0.0407731 | 0.00014395 | 0.0107262 | 3.80126233 | 0.3415 | eqtlPRS |
| rs12024920 | 1 | 84919467 | C | T | -0.0351624 | 0.00014718 | 0.00926355 | -3.7957802 | 0.4315 | eqtlPRS |
| rs167479 | 19 | 11526765 | G | T | 0.0417583 | 0.00014746 | 0.0110026 | 3.79531202 | 0.4611 | eqtlPRS |
| rs935972 | 4 | 1045171 | C | T | 0.0419292 | 0.00014978 | 0.0110589 | 3.791444 | 0.2485 | eqtlPRS |
| rs113470029 | 1 | 25234489 | T | C | 0.0353683 | 0.00015124 | 0.00933443 | 3.7890155 | 0.4356 | eqtlPRS |
| rs343084 | 7 | 35572165 | A | G | -0.0346936 | 0.00015313 | 0.00916378 | -3.7859486 | 0.4969 | eqtlPRS |
| rs639436 | 19 | 56032711 | G | A | -0.0446736 | 0.00015446 | 0.0118066 | -3.783782 | 0.2198 | eqtlPRS |
| rs694893 | 11 | 77563344 | C | T | 0.0361918 | 0.00015507 | 0.00956746 | 3.78280129 | 0.3466 | eqtlPRS |
| rs111811202 | 20 | 34441768 | C | T | 0.085913 | 0.00015618 | 0.0227221 | 3.78103256 | 0.0501 | eqtlPRS |
| rs4722409 | 7 | 947236 | T | C | -0.035526 | 0.00015882 | 0.00940626 | -3.7768465 | 0.4315 | eqtlPRS |
| rs10782504 | 14 | 20960462 | G | A | -0.036404 | 0.00015918 | 0.00964014 | -3.7762937 | 0.4826 | eqtlPRS |
| rs62204564 | 20 | 43300770 | A | G | 0.0551808 | 0.00015922 | 0.0146127 | 3.77622205 | 0.1575 | eqtlPRS |
| rs4246991 | 14 | 21915516 | A | G | -0.0609454 | 0.00015987 | 0.0161435 | -3.7752284 | 0.09305 | eqtlPRS |
| rs12971445 | 19 | 41484602 | G | T | -0.0362472 | 0.00016003 | 0.00960198 | -3.7749714 | 0.3528 | eqtlPRS |
| rs1002414 | 2 | 201616442 | G | T | 0.0385243 | 0.00016031 | 0.0102064 | 3.77452383 | 0.2863 | eqtlPRS |
| rs2997884 | 6 | 169902309 | C | T | -0.0604511 | 0.00016191 | 0.0160261 | -3.7720406 | 0.1329 | eqtlPRS |
| rs56913458 | 15 | 66702345 | C | A | 0.0406746 | 0.0001629 | 0.0107875 | 3.77053071 | 0.2331 | eqtlPRS |
| rs75972644 | 5 | 171818431 | G | A | 0.093985 | 0.00016318 | 0.0249291 | 3.77009198 | 0.03579 | eqtlPRS |
| rs215329 | 12 | 47726577 | A | G | 0.0346193 | 0.00016421 | 0.00918643 | 3.76852597 | 0.454 | eqtlPRS |
| rs828097 | 9 | 127959449 | C | T | -0.0379361 | 0.00016443 | 0.0100674 | -3.7682122 | 0.2996 | eqtlPRS |
| rs55893771 | 7 | 2096255 | C | T | -0.0478826 | 0.00016751 | 0.0127227 | -3.7635565 | 0.1595 | eqtlPRS |
| rs6093717 | 20 | 35584836 | T | C | 0.0458632 | 0.00016855 | 0.0121912 | 3.76199226 | 0.1339 | eqtlPRS |
| rs714006 | 22 | 49847719 | G | A | 0.0428075 | 0.00016913 | 0.0113815 | 3.76114748 | 0.2301 | eqtlPRS |
| rs1708403 | 7 | 134152364 | A | G | -0.0348813 | 0.00016953 | 0.00927554 | -3.7605681 | 0.4888 | eqtlPRS |
| rs408156 | 14 | 68581256 | G | A | 0.0407356 | 0.0001697 | 0.010833 | 3.76032493 | 0.2423 | eqtlPRS |
| rs61856558 | 1 | 248001319 | C | T | 0.0445657 | 0.00017123 | 0.0118587 | 3.75805948 | 0.1943 | eqtlPRS |
| rs36028781 | 15 | 59104413 | G | A | -0.171932 | 0.00017124 | 0.0457502 | -3.7580601 | 0.01022 | eqtlPRS |
| rs2677727 | 15 | 91390454 | A | G | -0.0421268 | 0.00017291 | 0.011217 | -3.7556209 | 0.3742 | eqtlPRS |
| rs10788542 | 10 | 88962741 | A | G | 0.0352366 | 0.00017508 | 0.00939016 | 3.75250262 | 0.4468 | eqtlPRS |
| rs30480 | 19 | 39807213 | G | A | 0.0574797 | 0.00017523 | 0.0153186 | 3.75228154 | 0.1104 | eqtlPRS |
| rs56688852 | 11 | 72315126 | T | C | -0.143677 | 0.00017649 | 0.0383088 | -3.750496 | 0.0184 | eqtlPRS |
| rs1952183 | 14 | 89844111 | A | G | -0.0345372 | 0.00017666 | 0.00920931 | -3.7502484 | 0.4816 | eqtlPRS |
| rs11781613 | 8 | 11502448 | T | G | 0.0397672 | 0.00017762 | 0.0106077 | 3.74889938 | 0.2362 | eqtlPRS |
| rs112614158 | 12 | 47751189 | C | T | 0.0744544 | 0.00017768 | 0.0198608 | 3.74881173 | 0.06646 | eqtlPRS |
| rs141823737 | 1 | 52785413 | G | A | 0.0751084 | 0.00017782 | 0.0200364 | 3.74859755 | 0.05521 | eqtlPRS |
| rs3856330 | 1 | 228107643 | C | T | -0.0376661 | 0.00017939 | 0.0100539 | -3.7464168 | 0.319 | eqtlPRS |
| rs673526 | 11 | 111734143 | T | C | 0.0455729 | 0.00018068 | 0.0121703 | 3.74459956 | 0.1677 | eqtlPRS |
| rs4807505 | 19 | 3750869 | G | A | -0.0531916 | 0.00018115 | 0.0142073 | -3.7439626 | 0.1411 | eqtlPRS |
| rs68084031 | 14 | 73621576 | A | G | 0.0441596 | 0.00018121 | 0.0117952 | 3.74386191 | 0.182 | eqtlPRS |
| rs10162419 | 14 | 35252097 | A | G | -0.0357581 | 0.0001824 | 0.0095553 | -3.7422268 | 0.3599 | eqtlPRS |
| rs6691734 | 1 | 6482601 | G | A | 0.0427695 | 0.00018353 | 0.0114336 | 3.74068535 | 0.1861 | eqtlPRS |
| rs739441 | 9 | 135770115 | T | C | 0.045514 | 0.00018381 | 0.0121686 | 3.74028237 | 0.1728 | eqtlPRS |
| rs17720590 | 3 | 186907100 | A | G | -0.139838 | 0.00018384 | 0.0373875 | -3.740234 | 0.01125 | eqtlPRS |
| rs12990411 | 2 | 220713825 | G | A | 0.0451572 | 0.00018438 | 0.0120757 | 3.73950992 | 0.4591 | eqtlPRS |
| rs74952329 | 8 | 143525501 | T | C | -0.0601565 | 0.00018611 | 0.0160968 | -3.7371714 | 0.09509 | eqtlPRS |
| rs2934429 | 15 | 57899577 | G | A | -0.0396194 | 0.00018633 | 0.0106023 | -3.7368684 | 0.2331 | eqtlPRS |
| rs2540034 | 16 | 4022694 | T | C | -0.0353348 | 0.00018781 | 0.00946077 | -3.7348757 | 0.4325 | eqtlPRS |
| rs4143632 | 15 | 61952231 | T | C | -0.0381376 | 0.0001889 | 0.0102152 | -3.7334169 | 0.2556 | eqtlPRS |
| rs984616 | 5 | 130891101 | T | C | -0.0402672 | 0.00018912 | 0.0107864 | -3.7331454 | 0.2699 | eqtlPRS |
| rs11666203 | 19 | 48345941 | G | T | -0.0345019 | 0.00019025 | 0.00924579 | -3.7316335 | 0.3916 | eqtlPRS |
| rs61156500 | 19 | 40873310 | C | T | -0.108292 | 0.00019116 | 0.0290295 | -3.7304122 | 0.03476 | eqtlPRS |
| rs6774451 | 3 | 196186948 | C | T | 0.0474701 | 0.00019123 | 0.0127254 | 3.73034246 | 0.1759 | eqtlPRS |
| rs6560667 | 10 | 134792456 | C | T | 0.0351916 | 0.00019261 | 0.0094385 | 3.72851618 | 0.3865 | eqtlPRS |
| rs3740888 | 11 | 130278210 | T | C | 0.0354732 | 0.00019566 | 0.00952414 | 3.72455676 | 0.3436 | eqtlPRS |
| rs8059420 | 16 | 10824380 | G | A | -0.0371799 | 0.00019915 | 0.00999437 | -3.7200844 | 0.3006 | eqtlPRS |
| rs666982 | 6 | 79259712 | C | T | -0.0343902 | 0.0001997 | 0.00924619 | -3.7193914 | 0.4284 | eqtlPRS |
| rs6694580 | 1 | 156524221 | G | T | 0.036489 | 0.00020075 | 0.00981396 | 3.71807099 | 0.3047 | eqtlPRS |
| rs11032362 | 11 | 33759092 | G | A | -0.058147 | 0.00020115 | 0.0156411 | -3.7175774 | 0.09305 | eqtlPRS |
| rs116837283 | 1 | 225515225 | A | G | -0.154691 | 0.00020291 | 0.0416355 | -3.7153631 | 0.02045 | eqtlPRS |
| rs13038055 | 20 | 36069515 | T | G | 0.0402939 | 0.00020508 | 0.0108531 | 3.71266274 | 0.2873 | eqtlPRS |
| rs12625240 | 20 | 42342316 | C | T | -0.0730086 | 0.00020662 | 0.0196747 | -3.7107859 | 0.06033 | eqtlPRS |
| rs2283865 | 22 | 30750907 | G | A | 0.0376064 | 0.00020785 | 0.0101384 | 3.70930324 | 0.2924 | eqtlPRS |
| rs73239429 | 8 | 27079082 | G | A | 0.039977 | 0.00020951 | 0.0107834 | 3.70727229 | 0.227 | eqtlPRS |
| rs2621326 | 6 | 32783896 | G | A | -0.0342985 | 0.00021133 | 0.00925715 | -3.705082 | 0.4407 | eqtlPRS |
| rs72936309 | 2 | 204040296 | G | T | -0.0512235 | 0.00021374 | 0.013836 | -3.7021899 | 0.1278 | eqtlPRS |
| rs4079029 | 15 | 83727830 | C | T | -0.0355551 | 0.00021739 | 0.00961494 | -3.6979014 | 0.3517 | eqtlPRS |
| rs58164846 | 19 | 41140115 | G | A | -0.0422373 | 0.00021984 | 0.0114307 | -3.6950755 | 0.228 | eqtlPRS |
| rs3737487 | 4 | 89608225 | C | T | -0.0459503 | 0.00021984 | 0.0124356 | -3.695061 | 0.1585 | eqtlPRS |
| rs76617690 | 19 | 10519141 | C | T | 0.109524 | 0.0002201 | 0.0296432 | 3.69474281 | 0.01943 | eqtlPRS |
| rs28485907 | 22 | 46273006 | C | T | -0.0460397 | 0.00022016 | 0.0124611 | -3.6946738 | 0.1912 | eqtlPRS |
| rs4433920 | 19 | 44571154 | C | T | 0.0362695 | 0.00022093 | 0.00981902 | 3.6938004 | 0.318 | eqtlPRS |
| rs75717807 | 6 | 111830362 | A | G | 0.124929 | 0.00022181 | 0.0338305 | 3.69279201 | 0.02352 | eqtlPRS |
| rs2269193 | 21 | 38106543 | C | A | -0.0454844 | 0.00022268 | 0.0123204 | -3.6917957 | 0.181 | eqtlPRS |
| rs61766261 | 1 | 84868876 | G | A | 0.0898645 | 0.00022484 | 0.0243579 | 3.68933693 | 0.04703 | eqtlPRS |
| rs137897580 | 16 | 11499998 | G | A | -0.119403 | 0.00022505 | 0.0323665 | -3.6890921 | 0.02658 | eqtlPRS |
| rs4400555 | 1 | 3779151 | T | C | -0.0795386 | 0.00022618 | 0.0215679 | -3.6878231 | 0.06033 | eqtlPRS |
| rs9905946 | 17 | 55748622 | C | T | 0.0338097 | 0.00022854 | 0.0091745 | 3.68518175 | 0.4928 | eqtlPRS |
| rs71362689 | 19 | 44907899 | G | A | 0.12678 | 0.00022955 | 0.0344131 | 3.68406217 | 0.02658 | eqtlPRS |
| rs12978319 | 19 | 17134159 | T | G | 0.0364853 | 0.00022979 | 0.00990428 | 3.68379125 | 0.3272 | eqtlPRS |
| rs6002503 | 22 | 42208672 | A | G | -0.0351397 | 0.00023069 | 0.00954157 | -3.6828006 | 0.3374 | eqtlPRS |
| rs2469205 | 15 | 77255029 | C | T | -0.0403924 | 0.00023106 | 0.0109691 | -3.6823805 | 0.2076 | eqtlPRS |
| rs12443371 | 15 | 68154234 | T | C | -0.0375108 | 0.00023275 | 0.0101917 | -3.6805243 | 0.2955 | eqtlPRS |
| rs117945402 | 8 | 38703278 | T | C | -0.0641003 | 0.0002333 | 0.0174189 | -3.6799281 | 0.07362 | eqtlPRS |
| rs748283 | 8 | 27317195 | C | A | -0.0336755 | 0.00023672 | 0.00916038 | -3.6762121 | 0.4765 | eqtlPRS |
| rs79257569 | 19 | 6965739 | G | A | 0.0531503 | 0.0002389 | 0.0144671 | 3.67387382 | 0.08384 | eqtlPRS |
| rs7100997 | 10 | 133111975 | G | A | 0.0349999 | 0.0002413 | 0.00953334 | 3.67131561 | 0.453 | eqtlPRS |
| rs11707923 | 3 | 58553669 | A | C | -0.039591 | 0.00024207 | 0.0107863 | -3.6704894 | 0.3139 | eqtlPRS |
| rs142577159 | 7 | 148693849 | A | G | 0.103613 | 0.00024264 | 0.0282332 | 3.66989927 | 0.03783 | eqtlPRS |
| rs4925276 | 20 | 60300480 | G | A | -0.0661614 | 0.0002429 | 0.0180295 | -3.6696192 | 0.07669 | eqtlPRS |
| rs393521 | 16 | 337678 | T | G | -0.0403692 | 0.00024387 | 0.0110039 | -3.6686266 | 0.2147 | eqtlPRS |
| rs77159830 | 16 | 11545894 | T | C | -0.108694 | 0.00024476 | 0.0296357 | -3.6676711 | 0.03476 | eqtlPRS |
| rs80243152 | 16 | 3630786 | T | C | -0.0673554 | 0.00024477 | 0.0183646 | -3.6676759 | 0.06953 | eqtlPRS |
| rs1174190 | 12 | 58770346 | G | T | -0.0354702 | 0.00024519 | 0.00967219 | -3.6672357 | 0.3374 | eqtlPRS |
| rs12199171 | 6 | 170827329 | C | T | -0.0847646 | 0.00024578 | 0.0231179 | -3.666622 | 0.04601 | eqtlPRS |
| rs34072093 | 2 | 112725747 | G | A | 0.160503 | 0.00024737 | 0.0437938 | 3.66497084 | 0.01534 | eqtlPRS |
| rs73033498 | 7 | 2911827 | G | A | 0.0489657 | 0.0002474 | 0.0133606 | 3.66493271 | 0.1186 | eqtlPRS |
| rs2614053 | 8 | 28150727 | A | G | 0.0355164 | 0.00024897 | 0.00969513 | 3.66332375 | 0.364 | eqtlPRS |
| rs1256358 | 1 | 201745951 | G | T | -0.0336424 | 0.00025235 | 0.00919226 | -3.6598617 | 0.4366 | eqtlPRS |
| rs10146820 | 14 | 22764676 | A | G | 0.0341138 | 0.00025327 | 0.00932343 | 3.65893239 | 0.4867 | eqtlPRS |
| rs6537045 | 4 | 142274255 | T | C | 0.0335813 | 0.00025481 | 0.00918179 | 3.65738053 | 0.4499 | eqtlPRS |
| rs7790633 | 7 | 63596962 | T | C | -0.035317 | 0.00025594 | 0.00965937 | -3.6562426 | 0.3303 | eqtlPRS |
| rs11136000 | 8 | 27464519 | C | T | 0.0338558 | 0.00025758 | 0.00926387 | 3.65460655 | 0.3896 | eqtlPRS |
| rs11610322 | 12 | 6390413 | A | G | 0.0469597 | 0.00025789 | 0.0128505 | 3.65430917 | 0.184 | eqtlPRS |
| rs2672603 | 10 | 124216834 | A | C | 0.0462405 | 0.00025938 | 0.0126589 | 3.65280554 | 0.1677 | eqtlPRS |
| rs4755308 | 11 | 45294152 | G | A | 0.033453 | 0.0002594 | 0.0091582 | 3.65279203 | 0.4632 | eqtlPRS |
| rs72902414 | 6 | 82957954 | C | T | 0.0514577 | 0.00025987 | 0.014089 | 3.65233161 | 0.1227 | eqtlPRS |
| rs62442504 | 7 | 2273301 | A | G | -0.0403785 | 0.0002625 | 0.0110634 | -3.649737 | 0.2321 | eqtlPRS |
| rs117768034 | 19 | 1960789 | G | A | -0.131499 | 0.00026284 | 0.0360329 | -3.6494148 | 0.02658 | eqtlPRS |
| rs17184822 | 14 | 24967452 | G | T | 0.0546999 | 0.000265 | 0.0149973 | 3.64731652 | 0.09816 | eqtlPRS |
| rs60486252 | 19 | 3768880 | G | A | -0.0418096 | 0.00026519 | 0.0114637 | -3.6471296 | 0.2117 | eqtlPRS |
| rs13288531 | 9 | 116861654 | C | T | 0.0772413 | 0.00026562 | 0.0211811 | 3.64670862 | 0.05726 | eqtlPRS |
| rs2595375 | 2 | 187561806 | A | G | 0.0340516 | 0.00026566 | 0.00933774 | 3.64666397 | 0.3701 | eqtlPRS |
| rs142531591 | 17 | 1148682 | T | C | 0.0888867 | 0.00026571 | 0.0243751 | 3.64661889 | 0.05521 | eqtlPRS |
| rs17109608 | 10 | 95739318 | A | G | 0.040428 | 0.00026589 | 0.0110869 | 3.64646565 | 0.18 | eqtlPRS |
| rs35673065 | 2 | 33003419 | A | G | 0.152301 | 0.00026663 | 0.0417753 | 3.64571888 | 0.02147 | eqtlPRS |
| rs17529477 | 11 | 113317067 | G | A | -0.0366182 | 0.00026783 | 0.0100473 | -3.6445811 | 0.3272 | eqtlPRS |
| rs76384858 | 4 | 103996276 | A | C | 0.0811916 | 0.00026902 | 0.0222844 | 3.64342769 | 0.04908 | eqtlPRS |
| rs142435690 | 20 | 62518690 | C | T | -0.145959 | 0.00027008 | 0.040072 | -3.6424186 | 0.01227 | eqtlPRS |
| rs11078256 | 17 | 1285765 | T | C | -0.0334126 | 0.00027161 | 0.00917683 | -3.6409741 | 0.4949 | eqtlPRS |
| rs8070971 | 17 | 18913742 | T | C | -0.0574174 | 0.00027195 | 0.0157712 | -3.6406488 | 0.1227 | eqtlPRS |
| rs17120523 | 11 | 117094591 | A | G | 0.0703964 | 0.00027289 | 0.0193409 | 3.63976857 | 0.06339 | eqtlPRS |
| rs73039485 | 19 | 33645524 | G | A | -0.0501092 | 0.00027358 | 0.0137696 | -3.6391181 | 0.1922 | eqtlPRS |
| rs3732073 | 2 | 36670011 | T | C | -0.0351099 | 0.00027611 | 0.00965422 | -3.6367412 | 0.3466 | eqtlPRS |
| rs1151676 | 1 | 247803638 | G | A | -0.0349939 | 0.00027639 | 0.00962301 | -3.6364817 | 0.407 | eqtlPRS |
| rs230661 | 11 | 77884782 | T | C | -0.0340414 | 0.00027643 | 0.00936118 | -3.6364433 | 0.4734 | eqtlPRS |
| rs62019405 | 15 | 91077146 | C | A | -0.0659719 | 0.00027741 | 0.0181464 | -3.6355365 | 0.07669 | eqtlPRS |
| rs6863305 | 5 | 66281007 | A | G | 0.0337356 | 0.00028322 | 0.00929308 | 3.63018504 | 0.4366 | eqtlPRS |
| rs7787619 | 7 | 100538028 | A | C | 0.0338723 | 0.00028347 | 0.00933132 | 3.62995803 | 0.4376 | eqtlPRS |
| rs45481494 | 20 | 61599540 | G | A | -0.0604949 | 0.00028512 | 0.0166724 | -3.6284458 | 0.09305 | eqtlPRS |
| rs12250629 | 10 | 15080584 | C | T | 0.0332809 | 0.00028548 | 0.00917302 | 3.62812901 | 0.4387 | eqtlPRS |
| rs6036860 | 20 | 2517377 | G | A | -0.0331783 | 0.0002875 | 0.00914934 | -3.6263053 | 0.456 | eqtlPRS |
| rs861823 | 22 | 22046738 | T | C | 0.0434511 | 0.00028799 | 0.0119836 | 3.62588037 | 0.2086 | eqtlPRS |
| rs3202848 | 17 | 8076655 | C | T | 0.0349278 | 0.00028804 | 0.00963304 | 3.62583359 | 0.3732 | eqtlPRS |
| rs7751410 | 6 | 122661338 | A | G | 0.032767 | 0.00028906 | 0.0090394 | 3.62490873 | 0.4744 | eqtlPRS |
| rs115074909 | 20 | 3739868 | G | A | 0.0899108 | 0.00029278 | 0.0248262 | 3.62160943 | 0.0409 | eqtlPRS |
| rs34493171 | 8 | 38685274 | T | G | 0.0448808 | 0.00029371 | 0.0123953 | 3.62079175 | 0.1708 | eqtlPRS |
| rs12988536 | 2 | 113293622 | G | A | 0.0470512 | 0.0002948 | 0.0129982 | 3.61982428 | 0.1247 | eqtlPRS |
| rs9894365 | 17 | 45284787 | A | G | 0.0340691 | 0.00029775 | 0.0094185 | 3.61725328 | 0.3609 | eqtlPRS |
| rs4393126 | 1 | 66952788 | A | G | -0.0333621 | 0.00029851 | 0.00922474 | -3.6165897 | 0.3998 | eqtlPRS |
| rs12464387 | 2 | 75445544 | G | A | 0.033351 | 0.00030026 | 0.00922553 | 3.61507686 | 0.4867 | eqtlPRS |
| rs2728108 | 4 | 88961736 | A | C | -0.0328684 | 0.00030077 | 0.00909315 | -3.614633 | 0.4734 | eqtlPRS |
| rs73001197 | 18 | 76855236 | G | A | 0.0582699 | 0.00030149 | 0.0161233 | 3.61401822 | 0.1012 | eqtlPRS |
| rs2079300 | 12 | 1117683 | T | C | -0.0340843 | 0.00030206 | 0.00943241 | -3.6135304 | 0.3906 | eqtlPRS |
| rs7976285 | 12 | 124852461 | T | C | 0.0432092 | 0.00030225 | 0.0119581 | 3.6133834 | 0.2188 | eqtlPRS |
| rs112069278 | 19 | 7094082 | C | T | 0.105877 | 0.00030435 | 0.029316 | 3.6115773 | 0.02249 | eqtlPRS |
| rs1610305 | 20 | 2152057 | G | A | 0.0337762 | 0.00030597 | 0.00935579 | 3.61019219 | 0.4182 | eqtlPRS |
| rs9613667 | 22 | 29127412 | A | C | 0.0347755 | 0.00030612 | 0.00963293 | 3.61006464 | 0.3262 | eqtlPRS |
| rs35243077 | 7 | 2604816 | C | T | -0.0482379 | 0.00031305 | 0.0133836 | -3.6042545 | 0.137 | eqtlPRS |
| rs59862264 | 2 | 68644910 | G | A | -0.0371305 | 0.00031352 | 0.010303 | -3.6038532 | 0.2914 | eqtlPRS |
| rs3761705 | 2 | 111903527 | A | G | -0.0358913 | 0.00031376 | 0.00995967 | -3.6036636 | 0.2986 | eqtlPRS |
| rs113441957 | 7 | 1562519 | C | A | 0.0731473 | 0.00031448 | 0.0203014 | 3.60306678 | 0.06135 | eqtlPRS |
| rs10447098 | 5 | 60831133 | C | T | 0.0403881 | 0.00031478 | 0.0112101 | 3.60283138 | 0.2065 | eqtlPRS |
| rs1104801 | 2 | 135436601 | A | G | -0.0352503 | 0.00031697 | 0.00978898 | -3.6010187 | 0.4325 | eqtlPRS |
| rs61349915 | 3 | 10615602 | C | T | -0.0473483 | 0.00031778 | 0.013151 | -3.6003574 | 0.1421 | eqtlPRS |
| rs9985005 | 21 | 40356274 | T | C | 0.0527274 | 0.00031842 | 0.0146472 | 3.59982795 | 0.08793 | eqtlPRS |
| rs142047940 | 17 | 80539630 | C | T | 0.0399183 | 0.0003197 | 0.0110921 | 3.59880455 | 0.2014 | eqtlPRS |
| rs76622989 | 6 | 56906187 | T | C | 0.139795 | 0.00032085 | 0.038855 | 3.59786385 | 0.01329 | eqtlPRS |
| rs191195 | 7 | 95310999 | G | A | 0.0335288 | 0.00032117 | 0.00931978 | 3.59759565 | 0.409 | eqtlPRS |
| rs72877189 | 4 | 88784746 | C | T | 0.0671991 | 0.00032473 | 0.0186938 | 3.59472659 | 0.06033 | eqtlPRS |
| rs2229333 | 18 | 3457607 | C | T | -0.0772957 | 0.00032566 | 0.021507 | -3.5939787 | 0.06033 | eqtlPRS |
| rs7106851 | 11 | 118554259 | G | A | 0.0354919 | 0.00032853 | 0.00988165 | 3.59169774 | 0.3088 | eqtlPRS |
| rs9561901 | 13 | 96206462 | A | G | -0.0402899 | 0.00032877 | 0.0112181 | -3.5915084 | 0.2311 | eqtlPRS |
| rs118114904 | 14 | 104630750 | C | T | 0.151512 | 0.00032922 | 0.0421904 | 3.5911487 | 0.01636 | eqtlPRS |
| rs73926782 | 2 | 47468102 | C | A | 0.161964 | 0.00033265 | 0.0451348 | 3.5884506 | 0.01329 | eqtlPRS |
| rs7638643 | 3 | 49565842 | C | T | -0.0485497 | 0.00033413 | 0.0135338 | -3.5872926 | 0.1462 | eqtlPRS |
| rs1531488 | 17 | 25690556 | T | G | -0.0330874 | 0.00033842 | 0.00923205 | -3.5839711 | 0.4366 | eqtlPRS |
| rs6007049 | 22 | 45858887 | A | C | 0.04842 | 0.00033858 | 0.0135107 | 3.58382615 | 0.1534 | eqtlPRS |
| rs34894704 | 15 | 74195522 | G | A | -0.0389804 | 0.00034052 | 0.0108812 | -3.5823622 | 0.2331 | eqtlPRS |
| rs35906296 | 2 | 128822001 | A | G | 0.0588176 | 0.00034166 | 0.0164227 | 3.58148173 | 0.07975 | eqtlPRS |
| rs76403931 | 5 | 1181213 | G | A | -0.0832661 | 0.00034177 | 0.0232497 | -3.5813838 | 0.03885 | eqtlPRS |
| rs56270936 | 18 | 13480890 | T | C | -0.0331673 | 0.00034353 | 0.00926447 | -3.5800537 | 0.4857 | eqtlPRS |
| rs34357052 | 5 | 141195546 | T | C | 0.0356438 | 0.00034787 | 0.00996535 | 3.57677352 | 0.3047 | eqtlPRS |
| rs1338571 | 1 | 100130644 | A | G | 0.0326612 | 0.00034842 | 0.00913254 | 3.57635444 | 0.4796 | eqtlPRS |
| rs2351012 | 5 | 96191747 | G | A | 0.0469964 | 0.00034849 | 0.013141 | 3.57631839 | 0.1503 | eqtlPRS |
| rs11125927 | 2 | 62752975 | A | G | -0.0518088 | 0.00034851 | 0.0144868 | -3.5762763 | 0.1278 | eqtlPRS |
| rs74089557 | 12 | 49930366 | G | A | -0.0664871 | 0.00035108 | 0.0186011 | -3.5743639 | 0.06339 | eqtlPRS |
| rs17481705 | 5 | 87582787 | G | A | -0.0706281 | 0.0003529 | 0.0197671 | -3.5730127 | 0.07873 | eqtlPRS |
| rs153317 | 5 | 76004246 | A | G | -0.0328311 | 0.00035308 | 0.00918897 | -3.5728814 | 0.4407 | eqtlPRS |
| rs45459391 | 1 | 26112130 | C | T | -0.0695372 | 0.0003534 | 0.0194638 | -3.5726425 | 0.08691 | eqtlPRS |
| rs78688545 | 5 | 402165 | C | A | 0.0693188 | 0.00035411 | 0.0194055 | 3.57212131 | 0.07771 | eqtlPRS |
| rs75904234 | 11 | 66100693 | C | T | 0.080714 | 0.00035655 | 0.022607 | 3.57031008 | 0.03476 | eqtlPRS |
| rs991968 | 2 | 56433558 | A | G | 0.037289 | 0.00035874 | 0.0104489 | 3.56870101 | 0.2904 | eqtlPRS |
| rs75741689 | 11 | 47183691 | G | A | -0.141129 | 0.00036071 | 0.0395621 | -3.5672778 | 0.02147 | eqtlPRS |
| rs142291688 | 17 | 57028391 | G | A | -0.130285 | 0.00036142 | 0.0365275 | -3.5667648 | 0.03681 | eqtlPRS |
| rs59926717 | 3 | 157807890 | C | T | 0.0607676 | 0.00036276 | 0.0170418 | 3.56579704 | 0.07566 | eqtlPRS |
| rs34288899 | 14 | 76028519 | C | T | 0.0501707 | 0.00036435 | 0.0140745 | 3.56465239 | 0.1708 | eqtlPRS |
| rs62061891 | 17 | 48871965 | A | G | 0.0472192 | 0.00036482 | 0.0132478 | 3.56430502 | 0.1288 | eqtlPRS |
| rs11932784 | 4 | 56144669 | G | T | -0.0652319 | 0.00036602 | 0.0183058 | -3.5634553 | 0.06748 | eqtlPRS |
| rs6052991 | 20 | 4944263 | A | G | 0.0334513 | 0.00036883 | 0.00939266 | 3.56142988 | 0.4162 | eqtlPRS |
| rs35967356 | 16 | 69581354 | C | T | -0.0329464 | 0.00036995 | 0.00925292 | -3.560649 | 0.4192 | eqtlPRS |
| rs6676027 | 1 | 223878433 | T | C | -0.0367639 | 0.00037043 | 0.0103261 | -3.560289 | 0.2883 | eqtlPRS |
| rs62329881 | 5 | 1590648 | C | T | -0.0403616 | 0.00037301 | 0.0113424 | -3.5584709 |  | eqtlPRS |
| rs117161993 | 8 | 101678108 | T | G | -0.102675 | 0.00037304 | 0.0288537 | -3.5584691 | 0.03272 | eqtlPRS |
| rs10916726 | 1 | 20555073 | C | T | 0.129194 | 0.00037395 | 0.0363127 | 3.55781861 | 0.02352 | eqtlPRS |
| rs10908521 | 1 | 156813650 | T | C | -0.037745 | 0.00037569 | 0.0106127 | -3.5565879 | 0.2791 | eqtlPRS |
| rs9613919 | 22 | 29856079 | G | A | 0.0558404 | 0.00037642 | 0.0157028 | 3.55607917 | 0.091 | eqtlPRS |
| rs4949657 | 1 | 77991508 | T | C | -0.0381052 | 0.00038052 | 0.0107241 | -3.5532306 | 0.2955 | eqtlPRS |
| rs4675128 | 2 | 227814757 | T | C | 0.0328815 | 0.00038092 | 0.00925466 | 3.55296683 | 0.4182 | eqtlPRS |
| rs803089 | 7 | 101946385 | C | T | 0.0350352 | 0.00038121 | 0.00986139 | 3.55276487 | 0.3129 | eqtlPRS |
| rs62065901 | 17 | 39935287 | A | G | -0.0593816 | 0.00038317 | 0.0167206 | -3.5514037 | 0.1196 | eqtlPRS |
| rs1547023 | 20 | 56333411 | G | A | 0.0328733 | 0.00038333 | 0.00925668 | 3.55130565 | 0.4284 | eqtlPRS |
| rs1130866 | 2 | 85893741 | A | G | -0.0326679 | 0.00038394 | 0.00919993 | -3.5508857 | 0.4591 | eqtlPRS |
| rs1973475 | 12 | 124643458 | C | T | -0.0380828 | 0.00038435 | 0.0107257 | -3.5506121 | 0.2382 | eqtlPRS |
| rs9457750 | 6 | 160279389 | G | A | 0.0406796 | 0.00038492 | 0.0114584 | 3.55019898 | 0.1973 | eqtlPRS |
| rs4980027 | 10 | 80843548 | C | A | -0.0522163 | 0.00038511 | 0.0147085 | -3.5500765 | 0.1166 | eqtlPRS |
| rs4234379 | 3 | 150239458 | T | C | -0.0334708 | 0.00038562 | 0.00942912 | -3.5497268 | 0.3926 | eqtlPRS |
| rs9608806 | 22 | 30095706 | G | A | -0.0334647 | 0.00038678 | 0.00942947 | -3.5489481 | 0.3845 | eqtlPRS |
| rs2909788 | 5 | 112193877 | G | A | -0.108078 | 0.00038688 | 0.0304542 | -3.5488701 | 0.03067 | eqtlPRS |
| rs1362366 | 2 | 202175769 | T | C | -0.0419724 | 0.00039017 | 0.0118344 | -3.5466437 | 0.1595 | eqtlPRS |
| rs112826255 | 14 | 50433640 | C | T | 0.0777039 | 0.00039043 | 0.0219102 | 3.54647151 | 0.03988 | eqtlPRS |
| rs79977374 | 7 | 1483403 | G | A | 0.110585 | 0.00039129 | 0.0311867 | 3.54590258 | 0.03272 | eqtlPRS |
| rs11630961 | 15 | 42447919 | G | A | 0.055336 | 0.00039257 | 0.0156095 | 3.54502066 | 0.08589 | eqtlPRS |
| rs870475 | 1 | 114533394 | G | A | 0.0331626 | 0.00039366 | 0.00935659 | 3.54430407 | 0.3701 | eqtlPRS |
| rs7778914 | 7 | 4989005 | A | G | -0.0557309 | 0.00039383 | 0.0157246 | -3.5441855 | 0.07771 | eqtlPRS |
| rs73132946 | 7 | 66909958 | G | A | -0.106962 | 0.00039769 | 0.0302013 | -3.5416356 | 0.03476 | eqtlPRS |
| rs1375514 | 3 | 100771328 | G | T | -0.03458 | 0.00039801 | 0.0097645 | -3.5414 | 0.3129 | eqtlPRS |
| rs35839418 | 1 | 24901506 | C | T | -0.0388462 | 0.00040247 | 0.0109783 | -3.5384531 | 0.2096 | eqtlPRS |
| rs112238703 | 15 | 90713642 | C | T | 0.134274 | 0.00040257 | 0.0379479 | 3.53837762 | 0.0184 | eqtlPRS |
| rs4982140 | 14 | 20967644 | C | T | 0.0380507 | 0.00040391 | 0.0107563 | 3.53752684 | 0.2372 | eqtlPRS |
| rs138189163 | 1 | 35523363 | T | C | 0.126755 | 0.00040416 | 0.0358334 | 3.53734226 | 0.01636 | eqtlPRS |
| rs1843020 | 12 | 117761668 | T | C | 0.0324738 | 0.00040436 | 0.00918058 | 3.5372275 | 0.4213 | eqtlPRS |
| rs2284191 | 6 | 32976654 | G | A | -0.0568477 | 0.00040507 | 0.0160734 | -3.5367564 | 0.1063 | eqtlPRS |
| rs116480793 | 1 | 156078376 | C | T | 0.113708 | 0.00040682 | 0.0321608 | 3.53560857 | 0.01943 | eqtlPRS |
| rs253487 | 5 | 60944590 | C | T | 0.0336634 | 0.00040814 | 0.00952353 | 3.53476075 | 0.3446 | eqtlPRS |
| rs843358 | 3 | 183861243 | A | G | -0.0345609 | 0.00041014 | 0.009781 | -3.5334731 | 0.3016 | eqtlPRS |
| rs78722528 | 20 | 56947524 | C | T | -0.158902 | 0.00041045 | 0.0449731 | -3.5332677 | 0.02045 | eqtlPRS |
| rs12958166 | 18 | 60200978 | G | T | -0.0360674 | 0.00041065 | 0.0102083 | -3.5331446 | 0.2873 | eqtlPRS |
| rs62482196 | 7 | 99935121 | G | A | -0.0930423 | 0.00041147 | 0.0263381 | -3.5326125 | 0.03783 | eqtlPRS |
| rs146987102 | 3 | 40428706 | G | A | 0.124303 | 0.00041172 | 0.0351887 | 3.53246923 | 0.03067 | eqtlPRS |
| rs6946669 | 7 | 94329691 | A | G | 0.0330928 | 0.00041247 | 0.00936949 | 3.53197453 | 0.3599 | eqtlPRS |
| rs6746588 | 2 | 242172493 | C | T | 0.0437163 | 0.00041601 | 0.0123852 | 3.52972096 | 0.1391 | eqtlPRS |
| rs2923438 | 8 | 42428746 | G | A | -0.0588846 | 0.00041765 | 0.0166875 | -3.5286652 | 0.09714 | eqtlPRS |
| rs144309607 | 19 | 10492274 | C | T | 0.0817376 | 0.0004189 | 0.023169 | 3.5278864 | 0.0317 | eqtlPRS |
| rs113334173 | 4 | 48062162 | T | G | 0.0519238 | 0.00042019 | 0.0147215 | 3.52707265 | 0.1063 | eqtlPRS |
| rs80154774 | 2 | 88427838 | T | C | 0.0555546 | 0.00042058 | 0.015752 | 3.52682834 | 0.07975 | eqtlPRS |
| rs2945330 | 4 | 48606526 | A | C | -0.0321592 | 0.0004214 | 0.00911981 | -3.5263015 | 0.4898 | eqtlPRS |
| rs36072812 | 7 | 150106635 | G | A | 0.0384726 | 0.00042144 | 0.0109102 | 3.52629649 | 0.2638 | eqtlPRS |
| rs72850292 | 11 | 2741358 | C | T | 0.0777503 | 0.00042455 | 0.022061 | 3.52433253 | 0.06135 | eqtlPRS |
| rs2283577 | 19 | 629503 | G | A | 0.0481558 | 0.00042482 | 0.0136644 | 3.52417962 | 0.1902 | eqtlPRS |
| rs2292334 | 6 | 160858188 | G | A | -0.0333063 | 0.0004281 | 0.0094563 | -3.5221281 | 0.3538 | eqtlPRS |
| rs699007 | 1 | 33197772 | T | C | 0.034484 | 0.00042971 | 0.00979345 | 3.52112892 | 0.3967 | eqtlPRS |
| rs4550232 | 11 | 124305112 | A | G | -0.0350713 | 0.00043056 | 0.00996174 | -3.5205998 | 0.3047 | eqtlPRS |
| rs53726 | 5 | 82599931 | C | A | 0.0372843 | 0.00043064 | 0.0105905 | 3.520542 | 0.2924 | eqtlPRS |
| rs140250479 | 7 | 43479617 | C | T | -0.127758 | 0.00043171 | 0.036296 | -3.519892 | 0.01227 | eqtlPRS |
| rs146219780 | 17 | 1285543 | C | T | -0.0636612 | 0.00043286 | 0.0180897 | -3.519196 | 0.07771 | eqtlPRS |
| rs6818658 | 4 | 56204605 | C | A | -0.0614099 | 0.00043389 | 0.0174531 | -3.5185669 | 0.1002 | eqtlPRS |
| rs61284774 | 6 | 126252677 | C | A | 0.0480201 | 0.00043804 | 0.0136574 | 3.51604991 | 0.1135 | eqtlPRS |
| rs4778955 | 15 | 82263265 | C | T | 0.0347427 | 0.00043807 | 0.00988126 | 3.51601921 | 0.274 | eqtlPRS |
| rs12436465 | 14 | 65806877 | C | T | 0.0373172 | 0.00043862 | 0.0106145 | 3.51568138 | 0.3241 | eqtlPRS |
| rs117406095 | 14 | 103781369 | C | T | -0.124276 | 0.00043912 | 0.0353521 | -3.5153782 | 0.02352 | eqtlPRS |
| rs147509466 | 19 | 52086328 | G | A | -0.0996591 | 0.00044044 | 0.0283558 | -3.5145931 | 0.009202 | eqtlPRS |
| rs34620667 | 4 | 84404208 | C | T | -0.0326346 | 0.00044263 | 0.00928897 | -3.5132636 | 0.3793 | eqtlPRS |
| rs75257146 | 3 | 44221819 | C | T | 0.0631447 | 0.00044556 | 0.0179822 | 3.51151138 | 0.07362 | eqtlPRS |
| rs6912090 | 6 | 170404195 | T | G | -0.0474285 | 0.0004456 | 0.0135066 | -3.5115055 | 0.1748 | eqtlPRS |
| rs17023076 | 2 | 39034166 | T | C | 0.0800644 | 0.00044606 | 0.0228025 | 3.51121149 | 0.03476 | eqtlPRS |
| rs76189127 | 15 | 78838837 | C | A | -0.0576392 | 0.00045433 | 0.0164386 | -3.5063327 | 0.0726 | eqtlPRS |
| rs4620062 | 5 | 1147890 | G | A | -0.03417 | 0.00045447 | 0.00974546 | -3.506248 | 0.4816 | eqtlPRS |
| rs13192826 | 6 | 25496657 | G | A | -0.0509631 | 0.00045533 | 0.014537 | -3.5057508 | 0.135 | eqtlPRS |
| rs11030383 | 11 | 3974623 | C | A | 0.0804633 | 0.0004575 | 0.0229601 | 3.50448387 | 0.06442 | eqtlPRS |
| rs8082941 | 18 | 56958687 | C | T | 0.0336563 | 0.00045843 | 0.00960528 | 3.50393742 | 0.3773 | eqtlPRS |
| rs55905121 | 5 | 1697239 | G | A | 0.0338277 | 0.00046102 | 0.00965832 | 3.50244142 | 0.3753 | eqtlPRS |
| rs79029250 | 15 | 63639021 | C | T | -0.0673852 | 0.0004619 | 0.0192423 | -3.5019306 | 0.08282 | eqtlPRS |
| rs1741623 | 20 | 62115643 | G | A | 0.0505679 | 0.00046201 | 0.0144403 | 3.50185938 | 0.1462 | eqtlPRS |
| rs35360485 | 4 | 99857325 | A | G | 0.0355145 | 0.00046418 | 0.0101452 | 3.50062098 | 0.272 | eqtlPRS |
| rs6822371 | 4 | 103233207 | C | A | 0.0335707 | 0.00046515 | 0.00959146 | 3.50006151 | 0.3599 | eqtlPRS |
| rs33995001 | 4 | 47514685 | C | T | 0.0321189 | 0.00046668 | 0.00917896 | 3.49918727 | 0.4366 | eqtlPRS |
| rs112888546 | 20 | 61925158 | C | T | 0.0783634 | 0.00046768 | 0.0223984 | 3.49861597 | 0.06442 | eqtlPRS |
| rs2806057 | 14 | 73474613 | C | T | -0.0343143 | 0.00046792 | 0.00980837 | -3.4984712 | 0.3149 | eqtlPRS |
| rs60799361 | 8 | 27558931 | T | C | -0.0477369 | 0.00046929 | 0.0136481 | -3.4976956 | 0.1258 | eqtlPRS |
| rs8072916 | 17 | 78223534 | G | A | -0.103578 | 0.00047178 | 0.0296251 | -3.496292 | 0.02863 | eqtlPRS |
| rs4802510 | 19 | 49310765 | C | T | 0.040763 | 0.00047181 | 0.011659 | 3.49626898 | 0.2014 | eqtlPRS |
| rs12527259 | 6 | 150907937 | C | T | -0.14749 | 0.00047222 | 0.0421876 | -3.496051 | 0.01022 | eqtlPRS |
| rs76152782 | 5 | 108757660 | C | T | -0.0773778 | 0.00047258 | 0.0221343 | -3.4958323 | 0.05624 | eqtlPRS |
| rs68136852 | 17 | 27382061 | C | A | 0.0446874 | 0.00047386 | 0.0127857 | 3.49510782 | 0.1575 | eqtlPRS |
| rs77763884 | 19 | 4508905 | C | T | 0.140634 | 0.00047399 | 0.0402382 | 3.49503705 | 0.01329 | eqtlPRS |
| rs56151242 | 10 | 102787296 | G | A | 0.0859341 | 0.00047639 | 0.0245969 | 3.49369636 | 0.04294 | eqtlPRS |
| rs17124174 | 12 | 50334781 | C | T | -0.0615183 | 0.00047651 | 0.0176087 | -3.493631 | 0.07362 | eqtlPRS |
| rs2269964 | 22 | 30866380 | T | C | 0.0373789 | 0.00047726 | 0.0107005 | 3.49319191 | 0.2321 | eqtlPRS |
| rs71578278 | 7 | 128230712 | G | A | 0.116809 | 0.00047788 | 0.0334422 | 3.49286231 | 0.01943 | eqtlPRS |
| rs79220634 | 11 | 117996402 | C | A | -0.0810177 | 0.0004779 | 0.0231953 | -3.4928498 | 0.05112 | eqtlPRS |
| rs2918015 | 2 | 86143565 | C | T | 0.0322842 | 0.00048039 | 0.00924663 | 3.49145581 | 0.4305 | eqtlPRS |
| rs147072238 | 5 | 139216744 | C | T | 0.116385 | 0.00048046 | 0.0333346 | 3.49141733 | 0.01227 | eqtlPRS |
| rs735055 | 20 | 61585807 | G | A | -0.0435838 | 0.00048089 | 0.012484 | -3.4911727 | 0.1708 | eqtlPRS |
| rs116852010 | 19 | 7430273 | A | G | 0.112214 | 0.00048124 | 0.0321439 | 3.49098896 | 0.0317 | eqtlPRS |
| rs79572662 | 13 | 114183543 | A | G | 0.108584 | 0.00048165 | 0.0311062 | 3.49075104 | 0.02147 | eqtlPRS |
| rs1583522 | 10 | 52014345 | A | G | 0.0439303 | 0.00048167 | 0.0125848 | 3.4907428 | 0.1585 | eqtlPRS |
| rs3811185 | 14 | 23369378 | T | C | 0.0364347 | 0.00048696 | 0.0104462 | 3.48784247 | 0.2658 | eqtlPRS |
| rs7999854 | 13 | 111250160 | C | T | 0.0507798 | 0.00049023 | 0.0145666 | 3.48604341 | 0.09918 | eqtlPRS |
| rs12693971 | 2 | 203415977 | T | C | 0.0363506 | 0.00049393 | 0.0104335 | 3.48402741 | 0.2607 | eqtlPRS |
| rs174771 | 22 | 29774736 | T | C | -0.0319642 | 0.0005018 | 0.00918564 | -3.4798011 | 0.4335 | eqtlPRS |
| rs461328 | 5 | 34860624 | T | C | 0.0911062 | 0.00050256 | 0.0261845 | 3.4793943 | 0.0409 | eqtlPRS |
| rs927332 | 6 | 6331873 | T | C | 0.0317008 | 0.00050733 | 0.00911766 | 3.476857 | 0.453 | eqtlPRS |
| rs9800194 | 5 | 149797134 | G | A | 0.0343508 | 0.0005079 | 0.00988069 | 3.47655882 | 0.4397 | eqtlPRS |
| rs17405738 | 2 | 187439007 | G | T | 0.0376755 | 0.00050793 | 0.0108371 | 3.4765297 | 0.2301 | eqtlPRS |
| rs143598667 | 7 | 73091122 | C | T | 0.075744 | 0.00050974 | 0.0217932 | 3.47557954 | 0.03885 | eqtlPRS |
| rs146145094 | 1 | 220427839 | G | A | -0.0607282 | 0.00051437 | 0.017485 | -3.4731599 | 0.07464 | eqtlPRS |
| rs11177790 | 12 | 70077983 | G | A | 0.0543871 | 0.00051493 | 0.0156606 | 3.47286183 | 0.1033 | eqtlPRS |
| rs59707717 | 22 | 32634524 | C | T | 0.124047 | 0.00051554 | 0.0357222 | 3.47254648 | 0.02249 | eqtlPRS |
| rs28618997 | 15 | 101047048 | G | A | -0.0526263 | 0.00051789 | 0.0151603 | -3.4713231 | 0.09714 | eqtlPRS |
| rs2059755 | 2 | 231429220 | T | C | 0.0318804 | 0.0005184 | 0.00918462 | 3.47106358 | 0.4407 | eqtlPRS |
| rs11928907 | 3 | 134037378 | A | C | 0.0375996 | 0.00051929 | 0.0108337 | 3.47061484 | 0.2178 | eqtlPRS |
| rs7939034 | 11 | 528585 | C | T | 0.0455124 | 0.00051968 | 0.0131145 | 3.47038774 | 0.1544 | eqtlPRS |
| rs4671797 | 2 | 67607135 | G | A | 0.0352462 | 0.00051977 | 0.0101564 | 3.47034382 | 0.2669 | eqtlPRS |
| rs3135878 | 4 | 1804377 | A | G | -0.0423135 | 0.00052248 | 0.0121977 | -3.4689737 | 0.2331 | eqtlPRS |
| rs1519312 | 4 | 187008844 | A | G | -0.0326205 | 0.00052261 | 0.00940374 | -3.4688858 | 0.3916 | eqtlPRS |
| rs113238829 | 16 | 4760867 | T | C | 0.0917817 | 0.00052275 | 0.0264591 | 3.46881413 | 0.02454 | eqtlPRS |
| rs62449806 | 7 | 24626860 | G | A | 0.0471616 | 0.00052638 | 0.0136032 | 3.46694895 | 0.1534 | eqtlPRS |
| rs9517510 | 13 | 99607550 | G | A | 0.0556158 | 0.00053111 | 0.0160528 | 3.46455447 | 0.08282 | eqtlPRS |
| rs232928 | 12 | 113112650 | C | T | 0.0327277 | 0.00053309 | 0.00944916 | 3.46355655 | 0.3793 | eqtlPRS |
| rs12450550 | 17 | 48456193 | T | C | 0.0353657 | 0.00053606 | 0.0102152 | 3.46206633 | 0.2423 | eqtlPRS |
| rs1322818 | 6 | 109691101 | A | G | 0.0358311 | 0.00053661 | 0.0103505 | 3.46177479 | 0.2751 | eqtlPRS |
| rs4725893 | 7 | 150112138 | G | A | -0.0396798 | 0.00053788 | 0.0114643 | -3.461162 | 0.2147 | eqtlPRS |
| rs5744256 | 11 | 112022848 | A | G | 0.0362373 | 0.00053844 | 0.0104706 | 3.46086184 | 0.2229 | eqtlPRS |
| rs111336205 | 11 | 57535606 | C | T | 0.0720445 | 0.0005412 | 0.0208252 | 3.45948658 | 0.05624 | eqtlPRS |
| rs115036942 | 17 | 78330135 | C | T | 0.0969141 | 0.00054193 | 0.0280169 | 3.45913003 | 0.02658 | eqtlPRS |
| rs3115406 | 4 | 8543598 | T | G | -0.0408366 | 0.00054785 | 0.0118154 | -3.4562182 | 0.1994 | eqtlPRS |
| rs7177866 | 15 | 44998963 | A | G | 0.083495 | 0.00055111 | 0.0241692 | 3.45460338 | 0.03783 | eqtlPRS |
| rs77064903 | 5 | 1091372 | C | T | -0.102926 | 0.00055127 | 0.0297947 | -3.454507 | 0.03374 | eqtlPRS |
| rs9789865 | 21 | 46728605 | A | G | 0.0415043 | 0.00055296 | 0.0120174 | 3.45368383 | 0.1892 | eqtlPRS |
| rs225183 | 17 | 30884028 | A | G | -0.0331193 | 0.00055304 | 0.00958964 | -3.4536542 | 0.4908 | eqtlPRS |
| rs3132557 | 6 | 31106409 | A | G | 0.0317199 | 0.00055624 | 0.00918859 | 3.45209657 | 0.4376 | eqtlPRS |
| rs12905138 | 15 | 83327999 | G | A | 0.0777977 | 0.00055843 | 0.0225433 | 3.45103423 | 0.03885 | eqtlPRS |
| rs75614054 | 9 | 98275789 | C | T | 0.0540704 | 0.00056088 | 0.0156732 | 3.44986346 | 0.091 | eqtlPRS |
| rs7692440 | 4 | 120008490 | C | A | 0.0330859 | 0.00056135 | 0.00959113 | 3.44963524 | 0.3978 | eqtlPRS |
| rs12362300 | 11 | 111307100 | C | T | 0.0528471 | 0.0005625 | 0.0153221 | 3.44907682 | 0.1002 | eqtlPRS |
| rs2209358 | 1 | 157274427 | G | T | 0.0333303 | 0.00056338 | 0.00966472 | 3.44865656 | 0.3384 | eqtlPRS |
| rs144112342 | 6 | 32858077 | T | C | 0.109417 | 0.000564 | 0.0317303 | 3.44834433 | 0.02045 | eqtlPRS |
| rs73342084 | 10 | 103034532 | T | C | -0.0630323 | 0.00056406 | 0.0182791 | -3.4483262 | 0.06851 | eqtlPRS |
| rs16852267 | 3 | 142032423 | C | T | -0.0393453 | 0.00056623 | 0.0114134 | -3.44729 | 0.1973 | eqtlPRS |
| rs2252878 | 22 | 25602708 | A | G | -0.0318069 | 0.00056644 | 0.00922688 | -3.4471999 | 0.454 | eqtlPRS |
| rs321110 | 11 | 112582512 | T | C | 0.0607574 | 0.00056675 | 0.0176259 | 3.44705235 | 0.1033 | eqtlPRS |
| rs7185312 | 16 | 15898999 | A | G | 0.0348418 | 0.00056858 | 0.0101103 | 3.44616876 | 0.3538 | eqtlPRS |
| rs4982815 | 14 | 24302116 | T | C | 0.0356189 | 0.00056981 | 0.0103375 | 3.44560097 | 0.316 | eqtlPRS |
| rs17581764 | 8 | 21766101 | C | T | -0.0492736 | 0.00057089 | 0.0143026 | -3.4450799 | 0.1411 | eqtlPRS |
| rs4646512 | 19 | 15764220 | C | T | 0.0328099 | 0.00057274 | 0.00952612 | 3.44420394 | 0.3517 | eqtlPRS |
| rs4926915 | 1 | 48083479 | C | T | -0.0316378 | 0.0005733 | 0.0091865 | -3.4439449 | 0.4427 | eqtlPRS |
| rs62205862 | 20 | 56817339 | G | A | -0.0647805 | 0.0005737 | 0.018811 | -3.4437563 | 0.0593 | eqtlPRS |
| rs10158601 | 1 | 16437298 | T | G | 0.0335794 | 0.0005739 | 0.00975108 | 3.44365957 | 0.4141 | eqtlPRS |
| rs7196129 | 16 | 30471109 | C | T | 0.0321259 | 0.00057451 | 0.0093298 | 3.44336427 | 0.4417 | eqtlPRS |
| rs12117457 | 1 | 95231974 | T | G | 0.0414468 | 0.00057566 | 0.0120386 | 3.44282558 | 0.1421 | eqtlPRS |
| rs17398598 | 1 | 52024908 | A | C | -0.0362265 | 0.00057857 | 0.0105265 | -3.4414573 | 0.2413 | eqtlPRS |
| rs4075835 | 8 | 95833822 | C | T | -0.0313655 | 0.00058554 | 0.00912257 | -3.4382307 | 0.4652 | eqtlPRS |
| rs1493990 | 1 | 34037669 | C | T | -0.032682 | 0.00058605 | 0.00950613 | -3.4379921 | 0.3354 | eqtlPRS |
| rs148628498 | 20 | 43114999 | G | A | 0.124726 | 0.00058701 | 0.0362834 | 3.43754995 | 0.01431 | eqtlPRS |
| rs12727188 | 1 | 159486328 | T | G | -0.0560227 | 0.00058816 | 0.0162998 | -3.4370176 | 0.09509 | eqtlPRS |
| rs1070361 | 1 | 226882660 | G | A | -0.044312 | 0.00058858 | 0.0128933 | -3.4368238 | 0.138 | eqtlPRS |
| rs138859050 | 3 | 157058595 | G | T | 0.116439 | 0.00059122 | 0.0338917 | 3.43561993 | 0.02556 | eqtlPRS |
| rs762009 | 14 | 35890861 | G | A | -0.0355929 | 0.00059297 | 0.0103624 | -3.4348124 | 0.2904 | eqtlPRS |
| rs7302963 | 12 | 8290569 | T | C | -0.0538999 | 0.00059438 | 0.0156952 | -3.4341646 |  | eqtlPRS |
| rs1443839 | 9 | 106662879 | C | T | -0.0320035 | 0.00059458 | 0.00931941 | -3.4340693 | 0.4029 | eqtlPRS |
| rs1455873 | 3 | 160508778 | G | T | 0.032187 | 0.00059579 | 0.00937433 | 3.43352538 | 0.4335 | eqtlPRS |
| rs12288228 | 11 | 125417616 | A | C | -0.120521 | 0.00059979 | 0.0351198 | -3.4317109 | 0.0184 | eqtlPRS |
| rs4723274 | 7 | 33235084 | C | T | 0.0326552 | 0.00060166 | 0.00951806 | 3.43086721 | 0.364 | eqtlPRS |
| rs10939621 | 4 | 15578068 | C | T | 0.0314223 | 0.00060478 | 0.00916246 | 3.42946108 | 0.4305 | eqtlPRS |
| rs9653568 | 2 | 85466885 | A | G | 0.0317981 | 0.00060536 | 0.00927273 | 3.42920585 | 0.3732 | eqtlPRS |
| rs117597345 | 19 | 18968723 | C | T | -0.149411 | 0.00060594 | 0.0435735 | -3.4289419 | 0.01636 | eqtlPRS |
| rs77793792 | 1 | 2301212 | C | T | 0.110391 | 0.0006081 | 0.0322028 | 3.42799384 | 0.0184 | eqtlPRS |
| rs76796396 | 1 | 112864075 | C | T | 0.105813 | 0.00060872 | 0.0308701 | 3.42768569 | 0.0184 | eqtlPRS |
| rs73163587 | 13 | 28112802 | C | T | 0.152955 | 0.00061087 | 0.0446356 | 3.42674905 | 0.02045 | eqtlPRS |
| rs60747167 | 4 | 2681789 | T | C | 0.126732 | 0.00061107 | 0.0369842 | 3.42665246 | 0.01738 | eqtlPRS |
| rs13178437 | 5 | 95882162 | A | G | 0.0477515 | 0.00061112 | 0.0139354 | 3.42663289 | 0.138 | eqtlPRS |
| rs17589398 | 4 | 151496759 | C | T | -0.0353178 | 0.00061357 | 0.0103101 | -3.4255536 | 0.2873 | eqtlPRS |
| rs6601648 | 8 | 11857173 | G | A | -0.0799274 | 0.00061654 | 0.0233417 | -3.4242322 | 0.04908 | eqtlPRS |
| rs117198731 | 21 | 38763723 | G | A | 0.0943315 | 0.00062234 | 0.0275687 | 3.42168836 | 0.02761 | eqtlPRS |
| rs116962538 | 6 | 107540817 | C | T | -0.0589985 | 0.0006266 | 0.0172519 | -3.4198262 | 0.08998 | eqtlPRS |
| rs74678824 | 5 | 1504472 | G | A | -0.0693761 | 0.000627 | 0.0202875 | -3.4196476 | 0.06033 | eqtlPRS |
| rs117506956 | 12 | 55154223 | C | T | -0.0659246 | 0.00062748 | 0.0192793 | -3.4194499 | 0.08078 | eqtlPRS |
| rs17636026 | 15 | 91493059 | T | C | -0.071336 | 0.00062984 | 0.0208681 | -3.4184233 | 0.04806 | eqtlPRS |
| rs73162142 | 22 | 32161567 | G | A | 0.0965282 | 0.00063174 | 0.0282444 | 3.41760491 | 0.0317 | eqtlPRS |
| rs115631735 | 5 | 154140442 | A | G | 0.0911355 | 0.00063271 | 0.0266697 | 3.41719254 | 0.02761 | eqtlPRS |
| rs147409473 | 19 | 55400136 | G | A | 0.081248 | 0.00063407 | 0.0237803 | 3.41660955 | 0.04499 | eqtlPRS |
| rs62393719 | 6 | 25679658 | A | G | -0.0864976 | 0.00063526 | 0.0253206 | -3.416096 | 0.03374 | eqtlPRS |
| rs35288226 | 9 | 139345366 | G | A | 0.0345599 | 0.00063594 | 0.0101176 | 3.41581996 | 0.3139 | eqtlPRS |
| rs9930725 | 16 | 1844176 | G | A | 0.0563947 | 0.00063684 | 0.0165118 | 3.41541806 | 0.0726 | eqtlPRS |
| rs10152984 | 15 | 84016917 | C | T | 0.0322785 | 0.00063702 | 0.00945103 | 3.41534203 | 0.3978 | eqtlPRS |
| rs61830201 | 10 | 4745126 | G | A | 0.128623 | 0.00063803 | 0.0376651 | 3.41491195 | 0.01943 | eqtlPRS |
| rs2738786 | 20 | 62325690 | T | C | -0.0923862 | 0.00063897 | 0.027057 | -3.4145027 | 0.02863 | eqtlPRS |
| rs12571524 | 10 | 90466543 | G | T | 0.0372407 | 0.00064787 | 0.0109186 | 3.41075779 | 0.2403 | eqtlPRS |
| rs11171007 | 12 | 55053543 | C | T | 0.0337052 | 0.00065193 | 0.00988701 | 3.40903873 | 0.2914 | eqtlPRS |
| rs62010551 | 15 | 78975416 | G | A | -0.125371 | 0.00065207 | 0.0367767 | -3.4089791 | 0.01636 | eqtlPRS |
| rs113173094 | 22 | 23856954 | T | C | 0.0465309 | 0.0006531 | 0.0136512 | 3.40855749 | 0.137 | eqtlPRS |
| rs12611168 | 19 | 49065294 | A | G | 0.0439938 | 0.00065343 | 0.0129074 | 3.40841688 | 0.1554 | eqtlPRS |
| rs549412 | 11 | 96048533 | G | A | 0.0330609 | 0.00065557 | 0.00970234 | 3.40751819 | 0.3671 | eqtlPRS |
| rs11264271 | 1 | 154802392 | T | C | 0.0474412 | 0.00065567 | 0.0139227 | 3.40747125 | 0.1186 | eqtlPRS |
| rs16909969 | 12 | 14702204 | C | T | 0.0492322 | 0.00065727 | 0.0144511 | 3.40681332 | 0.09611 | eqtlPRS |
| rs7842788 | 8 | 23406985 | G | A | 0.0590205 | 0.00065873 | 0.0173274 | 3.40619481 | 0.05726 | eqtlPRS |
| rs73149197 | 7 | 97732680 | G | A | 0.0612375 | 0.00066093 | 0.017983 | 3.40529945 | 0.08487 | eqtlPRS |
| rs6764523 | 3 | 46830734 | A | C | -0.0585904 | 0.00066133 | 0.0172065 | -3.4051318 | 0.09305 | eqtlPRS |
| rs74003282 | 16 | 4367269 | G | A | 0.0659167 | 0.00066278 | 0.0193615 | 3.40452444 | 0.06851 | eqtlPRS |
| rs75892544 | 8 | 104309006 | A | G | 0.0468092 | 0.00066338 | 0.0137501 | 3.4042807 | 0.1299 | eqtlPRS |
| rs13000695 | 2 | 220703292 | C | A | 0.0474894 | 0.00066584 | 0.013954 | 3.40328221 | 0.1227 | eqtlPRS |
| rs1065359 | 7 | 44259706 | G | A | -0.0319195 | 0.00066644 | 0.00937975 | -3.4030225 | 0.4121 | eqtlPRS |
| rs12138928 | 1 | 29554506 | T | C | -0.0345535 | 0.00066761 | 0.0101552 | -3.4025425 | 0.2464 | eqtlPRS |
| rs3744768 | 17 | 1800886 | G | A | 0.0375099 | 0.00066899 | 0.0110259 | 3.40198079 | 0.2168 | eqtlPRS |
| rs7678161 | 4 | 2901600 | C | T | -0.0334674 | 0.00066993 | 0.00983872 | -3.401601 | 0.3395 | eqtlPRS |
| rs10034495 | 4 | 164411073 | G | A | 0.0315622 | 0.00067118 | 0.00928005 | 3.40108081 | 0.4172 | eqtlPRS |
| rs74389777 | 3 | 129698420 | G | A | -0.0732554 | 0.00067263 | 0.0215425 | -3.400506 | 0.04499 | eqtlPRS |
| rs1728537 | 7 | 29690658 | A | C | -0.0379965 | 0.00067275 | 0.0111739 | -3.4004689 | 0.2485 | eqtlPRS |
| rs2116546 | 2 | 166723533 | T | C | 0.0335438 | 0.00067496 | 0.00986712 | 3.39955326 | 0.3088 | eqtlPRS |
| rs76846981 | 17 | 27202885 | T | C | -0.0521698 | 0.00068171 | 0.0153584 | -3.3968252 | 0.1207 | eqtlPRS |
| rs219500 | 4 | 109362923 | G | A | -0.033392 | 0.00068295 | 0.00983177 | -3.3963366 | 0.317 | eqtlPRS |
| rs16880636 | 5 | 52301091 | C | T | -0.0407204 | 0.00068326 | 0.01199 | -3.3961968 | 0.184 | eqtlPRS |
| rs1267469 | 11 | 34339834 | C | T | -0.0451175 | 0.00068443 | 0.0132865 | -3.39574 | 0.2076 | eqtlPRS |
| rs67585595 | 1 | 155243922 | C | A | 0.0782496 | 0.00068465 | 0.0230441 | 3.39564574 | 0.04192 | eqtlPRS |
| rs2249393 | 7 | 6697003 | C | T | -0.036238 | 0.00068542 | 0.0106728 | -3.3953602 | 0.2423 | eqtlPRS |
| rs77414117 | 1 | 10946903 | A | G | 0.119794 | 0.00068778 | 0.0352916 | 3.39440547 | 0.02965 | eqtlPRS |
| rs12575328 | 11 | 9314298 | G | A | -0.0407882 | 0.00069242 | 0.0120228 | -3.3925708 | 0.1605 | eqtlPRS |
| rs487624 | 6 | 25879539 | C | A | -0.031131 | 0.0006961 | 0.00918017 | -3.3911137 | 0.4356 | eqtlPRS |
| rs3754935 | 2 | 202119272 | A | C | -0.0571826 | 0.0006973 | 0.0168649 | -3.3906279 | 0.07362 | eqtlPRS |
| rs36081056 | 8 | 27338135 | C | T | -0.0330556 | 0.00070091 | 0.00975317 | -3.389216 | 0.3548 | eqtlPRS |
| rs8121408 | 20 | 17953571 | C | T | -0.0360127 | 0.0007023 | 0.0106274 | -3.3886651 | 0.2526 | eqtlPRS |
| rs11072972 | 15 | 81304132 | A | G | 0.039502 | 0.00070267 | 0.0116576 | 3.38851908 | 0.2045 | eqtlPRS |
| rs6665486 | 1 | 12117901 | A | G | 0.0330272 | 0.00070488 | 0.00974922 | 3.38767614 | 0.3221 | eqtlPRS |
| rs10931221 | 2 | 186941519 | G | T | -0.0378188 | 0.00070874 | 0.0111686 | -3.3861719 | 0.2423 | eqtlPRS |
| rs13068335 | 3 | 196552008 | G | T | 0.112133 | 0.00070968 | 0.0331185 | 3.38581156 | 0.02863 | eqtlPRS |
| rs35030867 | 15 | 50409400 | C | A | -0.0344117 | 0.0007098 | 0.0101636 | -3.3857787 | 0.2628 | eqtlPRS |
| rs59581960 | 19 | 42112638 | G | T | 0.0900906 | 0.00070981 | 0.0266087 | 3.38575729 | 0.04908 | eqtlPRS |
| rs12702093 | 7 | 44878540 | G | A | 0.0793455 | 0.00071359 | 0.0234451 | 3.38431058 | 0.04703 | eqtlPRS |
| rs138459820 | 11 | 33288193 | G | A | -0.0660656 | 0.00071557 | 0.0195255 | -3.3835548 | 0.07055 | eqtlPRS |
| rs114156841 | 6 | 25427508 | C | T | -0.10155 | 0.00071797 | 0.0300212 | -3.3826096 | 0.03579 | eqtlPRS |
| rs150616863 | 9 | 36362092 | G | A | 0.131097 | 0.00071863 | 0.038759 | 3.38236281 | 0.009202 | eqtlPRS |
| rs7038821 | 9 | 132169240 | T | C | -0.0342394 | 0.00072261 | 0.0101275 | -3.3808344 | 0.2853 | eqtlPRS |
| rs743271 | 14 | 24548567 | A | G | -0.0334506 | 0.00072533 | 0.00989716 | -3.379818 | 0.3425 | eqtlPRS |
| rs2023407 | 7 | 101957518 | C | A | 0.0372411 | 0.00072612 | 0.0110197 | 3.37950216 | 0.3344 | eqtlPRS |
| rs2477937 | 10 | 27222874 | A | G | 0.0424307 | 0.00072622 | 0.0125554 | 3.37947815 | 0.1984 | eqtlPRS |
| rs11950218 | 5 | 141076999 | C | T | -0.0687745 | 0.00072656 | 0.0203514 | -3.3793498 | 0.06135 | eqtlPRS |
| rs72669966 | 4 | 106514711 | A | G | -0.0619975 | 0.00073191 | 0.0183569 | -3.3773404 | 0.07464 | eqtlPRS |
| rs115414536 | 1 | 29180902 | C | T | 0.04986 | 0.00073234 | 0.0147638 | 3.37717932 | 0.1493 | eqtlPRS |
| rs1291126 | 20 | 35509409 | G | T | -0.0443165 | 0.00073282 | 0.0131231 | -3.3769841 | 0.1452 | eqtlPRS |
| rs144065514 | 10 | 70071300 | G | A | 0.0878263 | 0.00073316 | 0.0260082 | 3.3768696 | 0.0317 | eqtlPRS |
| rs34968118 | 17 | 6723997 | C | T | 0.0682135 | 0.00073636 | 0.0202074 | 3.37566931 | 0.05624 | eqtlPRS |
| rs7555137 | 1 | 160747381 | G | T | 0.0346584 | 0.00073966 | 0.0102708 | 3.37445963 | 0.2618 | eqtlPRS |
| rs73065423 | 12 | 12777584 | C | T | 0.0494727 | 0.00074088 | 0.014663 | 3.37398213 | 0.1094 | eqtlPRS |
| rs6690362 | 1 | 209608020 | A | C | -0.0321723 | 0.00074093 | 0.00953544 | -3.3739712 | 0.3241 | eqtlPRS |
| rs117361536 | 12 | 10479564 | A | C | -0.132788 | 0.00074095 | 0.0393568 | -3.3739532 | 0.01227 | eqtlPRS |
| rs115276522 | 1 | 227361336 | G | A | -0.115929 | 0.00074102 | 0.0343602 | -3.3739326 | 0.01943 | eqtlPRS |
| rs831605 | 11 | 33717125 | A | G | -0.033122 | 0.00074214 | 0.00981823 | -3.3735205 | 0.5 | eqtlPRS |
| rs57238953 | 16 | 27919526 | G | A | 0.0595637 | 0.00074255 | 0.017657 | 3.373376 | 0.08487 | eqtlPRS |
| rs1841178 | 3 | 48397046 | G | A | -0.0311117 | 0.00074295 | 0.00922314 | -3.3732221 | 0.4489 | eqtlPRS |
| rs1879793 | 8 | 22313986 | G | A | -0.0309503 | 0.00074641 | 0.0091788 | -3.3719332 | 0.4581 | eqtlPRS |
| rs74893260 | 19 | 49485719 | G | T | 0.0517827 | 0.00074832 | 0.0153601 | 3.37124758 | 0.1186 | eqtlPRS |
| rs74321826 | 1 | 153931835 | G | T | -0.0995497 | 0.00074928 | 0.0295322 | -3.3708867 | 0.02863 | eqtlPRS |
| rs2265549 | 10 | 72540534 | G | A | 0.0432456 | 0.00074976 | 0.0128298 | 3.37071505 | 0.1708 | eqtlPRS |
| rs116289854 | 1 | 110920474 | C | T | 0.11365 | 0.00074982 | 0.0337172 | 3.37068321 | 0.01431 | eqtlPRS |
| rs56739653 | 15 | 40426441 | C | T | 0.0928596 | 0.00075254 | 0.0275573 | 3.36969152 | 0.03988 | eqtlPRS |
| rs58598743 | 14 | 101810277 | T | G | 0.0935723 | 0.00075364 | 0.0277722 | 3.36927935 | 0.02658 | eqtlPRS |
| rs73094281 | 7 | 32819356 | A | G | -0.0523921 | 0.00075558 | 0.0155532 | -3.3685737 | 0.09816 | eqtlPRS |
| rs77196557 | 2 | 38099388 | G | A | 0.10579 | 0.00075797 | 0.0314132 | 3.36769256 | 0.02863 | eqtlPRS |
| rs9626457 | 22 | 44883779 | C | T | -0.0371382 | 0.00075858 | 0.0110285 | -3.3674752 | 0.2587 | eqtlPRS |
| rs79780973 | 5 | 150174810 | T | C | -0.0583785 | 0.00076611 | 0.01735 | -3.364755 | 0.1053 | eqtlPRS |
| rs10518980 | 15 | 44822774 | G | A | 0.0389554 | 0.00077086 | 0.0115833 | 3.36306579 | 0.1943 | eqtlPRS |
| rs7291412 | 22 | 46459132 | G | T | -0.0322357 | 0.000771 | 0.00958541 | -3.3629965 | 0.3354 | eqtlPRS |
| rs149074870 | 19 | 41606220 | A | C | 0.0687285 | 0.00077171 | 0.0204382 | 3.36274721 | 0.04397 | eqtlPRS |
| rs56083862 | 10 | 127718651 | G | T | 0.0579072 | 0.00077491 | 0.017226 | 3.36161616 | 0.06748 | eqtlPRS |
| rs9491385 | 6 | 125633031 | T | C | 0.030946 | 0.00077511 | 0.00920592 | 3.36153258 | 0.4499 | eqtlPRS |
| rs149386393 | 1 | 229414963 | A | G | 0.0985842 | 0.00077745 | 0.0293344 | 3.36070279 | 0.02045 | eqtlPRS |
| rs115512007 | 8 | 57078062 | G | A | -0.0822776 | 0.00078184 | 0.0244936 | -3.3591469 | 0.03885 | eqtlPRS |
| rs7803124 | 7 | 4888463 | T | C | -0.0314015 | 0.00078794 | 0.00935406 | -3.3569915 | 0.3855 | eqtlPRS |
| rs1182473 | 20 | 58323914 | G | A | -0.0335903 | 0.00078831 | 0.0100064 | -3.3568816 | 0.3211 | eqtlPRS |
| rs3810859 | 5 | 851582 | G | A | 0.0352335 | 0.00078908 | 0.0104968 | 3.35659439 | 0.2382 | eqtlPRS |
| rs2273113 | 1 | 17287414 | T | C | -0.032271 | 0.00079122 | 0.00961634 | -3.3558506 | 0.4325 | eqtlPRS |
| rs66494567 | 18 | 71690376 | A | G | -0.0482872 | 0.00079574 | 0.0143957 | -3.3542794 | 0.1043 | eqtlPRS |
| rs1154003 | 7 | 158616885 | A | G | -0.036769 | 0.00079726 | 0.0109636 | -3.3537342 | 0.2229 | eqtlPRS |
| rs76737203 | 15 | 80134882 | T | C | -0.0561977 | 0.00079766 | 0.0167574 | -3.353605 | 0.08998 | eqtlPRS |
| rs12322269 | 12 | 8525952 | G | A | -0.0386844 | 0.00080404 | 0.0115428 | -3.3513879 |  | eqtlPRS |
| rs1340980 | 1 | 161677389 | A | G | 0.0319046 | 0.00080434 | 0.00952008 | 3.35129537 | 0.3405 | eqtlPRS |
| rs12434998 | 14 | 94455554 | T | C | -0.0328881 | 0.00080489 | 0.00981409 | -3.3511105 | 0.3517 | eqtlPRS |
| rs9426359 | 1 | 29781530 | C | T | 0.0478022 | 0.00080746 | 0.0142683 | 3.35023794 | 0.09816 | eqtlPRS |
| rs16901235 | 5 | 31507079 | G | A | 0.102151 | 0.00080777 | 0.0304916 | 3.35013578 | 0.02454 | eqtlPRS |
| rs2422 | 12 | 120565188 | A | G | 0.0318583 | 0.00080835 | 0.00951017 | 3.34991909 | 0.4274 | eqtlPRS |
| rs9567670 | 13 | 32984197 | T | G | 0.0394389 | 0.00080862 | 0.0117734 | 3.34983097 | 0.1595 | eqtlPRS |
| rs12577841 | 11 | 118194599 | C | T | -0.0338379 | 0.00080872 | 0.0101015 | -3.3497896 | 0.317 | eqtlPRS |
| rs17886276 | 19 | 4934696 | C | T | 0.130529 | 0.00080907 | 0.0389676 | 3.34968025 | 0.01534 | eqtlPRS |
| rs80341248 | 4 | 146981453 | C | T | 0.0917073 | 0.0008101 | 0.0273809 | 3.34931649 | 0.03885 | eqtlPRS |
| rs62130411 | 19 | 3939373 | G | A | -0.0577452 | 0.00081141 | 0.0172432 | -3.348868 | 0.08793 | eqtlPRS |
| rs12508543 | 4 | 5871485 | G | T | 0.0313986 | 0.00081345 | 0.00937781 | 3.34818044 | 0.3988 | eqtlPRS |
| rs140895602 | 6 | 31024244 | A | G | -0.105085 | 0.00081432 | 0.0313885 | -3.3478822 | 0.03374 | eqtlPRS |
| rs79213347 | 15 | 42791676 | A | C | -0.063548 | 0.00081676 | 0.0189863 | -3.347045 | 0.07055 | eqtlPRS |
| rs595564 | 15 | 46008404 | G | A | -0.0376427 | 0.00081722 | 0.011247 | -3.3469103 | 0.2157 | eqtlPRS |
| rs17582478 | 15 | 44271206 | A | G | 0.0464465 | 0.00081939 | 0.0138805 | 3.34616909 | 0.1442 | eqtlPRS |
| rs139222955 | 5 | 180301417 | T | C | 0.0792895 | 0.00082268 | 0.0237035 | 3.34505453 | 0.05521 | eqtlPRS |
| rs16891512 | 6 | 26270609 | G | A | -0.0823135 | 0.00082606 | 0.0246159 | -3.3439159 | 0.06339 | eqtlPRS |
| rs12574466 | 11 | 119813518 | G | A | -0.0518087 | 0.00082672 | 0.0154945 | -3.3436832 | 0.1094 | eqtlPRS |
| rs1739124 | 1 | 25370099 | G | A | 0.0427674 | 0.0008272 | 0.0127911 | 3.34352792 | 0.1524 | eqtlPRS |
| rs12616906 | 2 | 153336661 | A | C | -0.0330104 | 0.00082739 | 0.0098731 | -3.3434686 | 0.2863 | eqtlPRS |
| rs61820088 | 1 | 204531554 | A | G | -0.11184 | 0.00083006 | 0.0334593 | -3.3425684 | 0.02045 | eqtlPRS |
| rs4652452 | 1 | 179708950 | A | G | 0.0350615 | 0.00083078 | 0.0104901 | 3.34234183 | 0.2423 | eqtlPRS |
| rs150173588 | 7 | 98432090 | A | C | 0.114121 | 0.00083337 | 0.0341529 | 3.3414732 | 0.01943 | eqtlPRS |
| rs55964797 | 22 | 23980989 | A | G | 0.140732 | 0.00083612 | 0.0421283 | 3.3405573 | 0.01636 | eqtlPRS |
| rs12608795 | 19 | 58896477 | G | A | -0.031845 | 0.00083688 | 0.00953358 | -3.3402982 | 0.3507 | eqtlPRS |
| rs35010362 | 19 | 57419162 | C | T | -0.0307077 | 0.00083773 | 0.00919386 | -3.3400226 | 0.41 | eqtlPRS |
| rs145688653 | 19 | 24116402 | C | T | 0.121383 | 0.00084168 | 0.0363562 | 3.33871527 | 0.03272 | eqtlPRS |
| rs1052165 | 12 | 56351346 | G | A | 0.0336173 | 0.00084216 | 0.0100694 | 3.33856039 | 0.271 | eqtlPRS |
| rs111234526 | 21 | 38255219 | C | A | 0.102024 | 0.00084249 | 0.0305604 | 3.33843798 | 0.02352 | eqtlPRS |
| rs906278 | 1 | 154718738 | C | T | 0.0402699 | 0.00084384 | 0.0120641 | 3.33799455 | 0.181 | eqtlPRS |
| rs77206031 | 19 | 58464819 | G | A | 0.164731 | 0.00084409 | 0.0493515 | 3.33791273 | 0.01431 | eqtlPRS |
| rs12514505 | 5 | 10760119 | G | A | -0.101053 | 0.00084443 | 0.0302753 | -3.3378034 | 0.03272 | eqtlPRS |
| rs11685572 | 2 | 47875888 | C | T | 0.030964 | 0.00084465 | 0.00927695 | 3.33773492 | 0.4121 | eqtlPRS |
| rs75248668 | 21 | 33655252 | G | A | -0.0986601 | 0.00084841 | 0.0295699 | -3.3365044 | 0.03476 | eqtlPRS |
| rs2151761 | 14 | 101811705 | T | C | 0.037588 | 0.00084937 | 0.0112668 | 3.33617354 | 0.2014 | eqtlPRS |
| rs2304970 | 17 | 4642787 | A | C | 0.0312778 | 0.00085026 | 0.00937613 | 3.33589658 | 0.4274 | eqtlPRS |
| rs56221686 | 5 | 608706 | C | T | -0.0635527 | 0.0008544 | 0.0190589 | -3.3345419 | 0.06748 | eqtlPRS |
| rs59252423 | 6 | 37114558 | C | T | 0.119613 | 0.0008558 | 0.0358757 | 3.33409522 | 0.02045 | eqtlPRS |
| rs149847304 | 17 | 76308339 | C | T | -0.134869 | 0.00085827 | 0.0404613 | -3.3332839 | 0.01431 | eqtlPRS |
| rs73059125 | 19 | 53896634 | G | A | -0.13593 | 0.00085834 | 0.0407798 | -3.3332679 | 0.0184 | eqtlPRS |
| rs1019474 | 19 | 14552518 | G | A | -0.038877 | 0.0008587 | 0.0116638 | -3.3331333 | 0.3037 | eqtlPRS |
| rs12459901 | 19 | 57287998 | T | C | -0.0304324 | 0.00086198 | 0.00913314 | -3.3320851 | 0.4519 | eqtlPRS |
| rs73018364 | 19 | 17155987 | C | T | -0.101457 | 0.00086304 | 0.0304516 | -3.3317461 | 0.01329 | eqtlPRS |
| rs36166264 | 8 | 144026403 | C | T | -0.0335684 | 0.0008638 | 0.0100761 | -3.3314874 | 0.3323 | eqtlPRS |
| rs11632005 | 15 | 86254487 | C | T | 0.0363454 | 0.00086509 | 0.010911 | 3.33107873 | 0.2382 | eqtlPRS |
| rs146341789 | 15 | 45788656 | C | T | -0.127028 | 0.00086771 | 0.0381437 | -3.3302485 | 0.01431 | eqtlPRS |
| rs158391 | 10 | 43143078 | G | T | -0.0336759 | 0.00087041 | 0.0101148 | -3.3293688 | 0.3354 | eqtlPRS |
| rs7869570 | 9 | 136036303 | A | G | 0.0386682 | 0.00087213 | 0.0116162 | 3.32881665 | 0.183 | eqtlPRS |
| rs76796388 | 1 | 159450413 | C | T | -0.0958987 | 0.00087392 | 0.0288135 | -3.3282559 | 0.0317 | eqtlPRS |
| rs6496714 | 15 | 91174904 | T | C | 0.0384578 | 0.0008793 | 0.0115609 | 3.32654032 | 0.2004 | eqtlPRS |
| rs10186582 | 2 | 239017580 | C | A | 0.043198 | 0.00088115 | 0.0129881 | 3.32596762 | 0.137 | eqtlPRS |
| rs8042947 | 15 | 40325860 | A | G | -0.0348229 | 0.00088344 | 0.0104723 | -3.325239 | 0.2883 | eqtlPRS |
| rs11683600 | 2 | 48835096 | A | G | 0.0307155 | 0.00088411 | 0.00923769 | 3.32501957 | 0.4264 | eqtlPRS |
| rs6889109 | 5 | 130474409 | T | G | -0.0348689 | 0.00088617 | 0.0104889 | -3.3243619 | 0.272 | eqtlPRS |
| rs11668173 | 19 | 53115594 | G | A | -0.0740369 | 0.00088681 | 0.0222723 | -3.3241695 | 0.05112 | eqtlPRS |
| rs573010 | 5 | 31444434 | C | A | -0.0344448 | 0.00088959 | 0.0103646 | -3.323312 | 0.2658 | eqtlPRS |
| rs3740468 | 10 | 105991895 | G | T | -0.0882208 | 0.00089122 | 0.0265502 | -3.3227923 | 0.0317 | eqtlPRS |
| rs57780061 | 15 | 89526948 | G | T | -0.0326218 | 0.00089291 | 0.00981915 | -3.3222631 | 0.3599 | eqtlPRS |
| rs4505789 | 4 | 89825615 | A | C | 0.0305637 | 0.00089696 | 0.00920317 | 3.320997 | 0.4233 | eqtlPRS |
| rs34882515 | 8 | 142201124 | A | G | 0.0393164 | 0.00089719 | 0.011839 | 3.32092238 | 0.1892 | eqtlPRS |
| rs901886 | 19 | 10402131 | T | C | -0.0313831 | 0.00089733 | 0.00945021 | -3.3208892 | 0.4775 | eqtlPRS |
| rs147290493 | 11 | 125395432 | G | A | 0.118474 | 0.00090085 | 0.0356873 | 3.31978043 | 0.01738 | eqtlPRS |
| rs11726253 | 4 | 8524676 | C | T | -0.0532433 | 0.00090347 | 0.0160421 | -3.3189732 | 0.08589 | eqtlPRS |
| rs4962054 | 9 | 136336933 | G | A | -0.0788813 | 0.00090449 | 0.023769 | -3.318663 | 0.04397 | eqtlPRS |
| rs2032423 | 14 | 54425147 | C | T | -0.030684 | 0.00090607 | 0.00924726 | -3.3181721 | 0.4008 | eqtlPRS |
| rs11931318 | 4 | 110564140 | A | G | 0.0307525 | 0.00090764 | 0.00926922 | 3.31770095 | 0.4417 | eqtlPRS |
| rs12765929 | 10 | 88682555 | G | T | 0.0341153 | 0.00090916 | 0.0102843 | 3.3172214 | 0.2751 | eqtlPRS |
| rs12638831 | 3 | 158521218 | A | C | -0.0328168 | 0.00091342 | 0.00989673 | -3.3159235 | 0.3395 | eqtlPRS |
| rs7507400 | 19 | 41330179 | G | T | -0.0406817 | 0.00091609 | 0.0122716 | -3.3151097 | 0.316 | eqtlPRS |
| rs2665116 | 15 | 82403864 | G | A | 0.0315976 | 0.000919 | 0.00953396 | 3.31421571 | 0.3855 | eqtlPRS |
| rs78689927 | 19 | 5413875 | T | C | 0.0613396 | 0.00092076 | 0.018511 | 3.31368376 | 0.07362 | eqtlPRS |
| rs56170012 | 3 | 50588176 | A | G | -0.149977 | 0.00092089 | 0.0452605 | -3.3136399 | 0.01125 | eqtlPRS |
| rs6289 | 4 | 47408709 | A | G | 0.0325938 | 0.00092122 | 0.00983653 | 3.31354655 | 0.3088 | eqtlPRS |
| rs77449251 | 1 | 168876181 | C | T | -0.0803656 | 0.00092173 | 0.0242548 | -3.3133895 | 0.03988 | eqtlPRS |
| rs1981583 | 1 | 55171439 | A | G | 0.0331538 | 0.00092315 | 0.0100073 | 3.31296154 | 0.3221 | eqtlPRS |
| rs16990494 | 19 | 530109 | A | G | 0.0931605 | 0.00092355 | 0.0281211 | 3.31283271 | 0.03681 | eqtlPRS |
| rs17581463 | 17 | 57858706 | C | T | -0.077507 | 0.00092499 | 0.023399 | -3.3124065 | 0.05112 | eqtlPRS |
| rs72745571 | 15 | 57993438 | A | C | 0.0661482 | 0.00092537 | 0.0199705 | 3.31229564 | 0.06646 | eqtlPRS |
| rs2064279 | 20 | 42094474 | G | T | 0.0315065 | 0.00092575 | 0.00951235 | 3.31216787 | 0.4059 | eqtlPRS |
| rs58582808 | 17 | 45056864 | G | A | -0.034301 | 0.00092583 | 0.0103561 | -3.3121542 | 0.3057 | eqtlPRS |
| rs139127662 | 2 | 242507737 | G | A | 0.0888904 | 0.00092649 | 0.0268393 | 3.31194927 | 0.04499 | eqtlPRS |
| rs35999920 | 1 | 249168433 | G | A | 0.0509755 | 0.00092726 | 0.0153925 | 3.31171025 | 0.09407 | eqtlPRS |
| rs76624186 | 17 | 72654959 | T | C | -0.059265 | 0.00092759 | 0.0178961 | -3.3116154 | 0.07873 | eqtlPRS |
| rs74508318 | 19 | 57172210 | G | T | 0.0375385 | 0.00092813 | 0.011336 | 3.31144143 | 0.227 | eqtlPRS |
| rs62182790 | 2 | 219081809 | G | A | -0.0883581 | 0.00092901 | 0.0266847 | -3.3111896 | 0.04601 | eqtlPRS |
| rs12489383 | 3 | 149829015 | C | T | 0.0752557 | 0.00092948 | 0.0227287 | 3.31104287 | 0.04397 | eqtlPRS |
| rs117404771 | 12 | 55971207 | C | T | -0.110999 | 0.00093439 | 0.0335386 | -3.3095895 | 0.0317 | eqtlPRS |
| rs12807728 | 11 | 9592236 | C | T | -0.0907625 | 0.00093476 | 0.0274252 | -3.3094563 | 0.03579 | eqtlPRS |
| rs6468804 | 8 | 102972616 | A | G | 0.0307857 | 0.00093669 | 0.00930396 | 3.30888138 | 0.3691 | eqtlPRS |
| rs739190 | 22 | 37758272 | C | T | -0.0765114 | 0.00093698 | 0.0231236 | -3.3088014 | 0.03783 | eqtlPRS |
| rs7033339 | 9 | 123503438 | A | G | -0.0331823 | 0.00093722 | 0.0100287 | -3.3087339 | 0.3149 | eqtlPRS |
| rs79088892 | 8 | 17440946 | G | T | -0.0877327 | 0.00093787 | 0.0265171 | -3.308533 | 0.03885 | eqtlPRS |
| rs2952286 | 17 | 66469400 | G | T | 0.0373049 | 0.00093885 | 0.0112764 | 3.3082278 | 0.2117 | eqtlPRS |
| rs4574552 | 5 | 43731692 | A | G | 0.0318227 | 0.0009439 | 0.00962359 | 3.30673896 | 0.409 | eqtlPRS |
| rs8183571 | 20 | 3790679 | T | C | 0.031749 | 0.00094703 | 0.009604 | 3.30581008 | 0.3885 | eqtlPRS |
| rs9989432 | 16 | 31225189 | C | T | 0.0438405 | 0.00094723 | 0.0132619 | 3.30574805 | 0.135 | eqtlPRS |
| rs74890890 | 1 | 89723013 | A | G | 0.0887899 | 0.00094774 | 0.0268605 | 3.30559372 | 0.0409 | eqtlPRS |
| rs10857567 | 10 | 49651113 | T | C | 0.0315769 | 0.00094973 | 0.00955424 | 3.30501432 | 0.3129 | eqtlPRS |
| rs9649862 | 7 | 56303746 | T | C | -0.0317948 | 0.00094998 | 0.0096204 | -3.3049353 | 0.316 | eqtlPRS |
| rs3851674 | 12 | 132085047 | G | A | -0.0424516 | 0.00095071 | 0.0128458 | -3.3047066 | 0.1503 | eqtlPRS |
| rs9424135 | 10 | 5700843 | G | T | -0.0534114 | 0.00095264 | 0.0161649 | -3.304159 | 0.09611 | eqtlPRS |
| rs72999491 | 11 | 118539151 | G | A | -0.0457152 | 0.00095424 | 0.0138377 | -3.3036704 | 0.1217 | eqtlPRS |
| rs6076369 | 20 | 25576783 | G | A | 0.0304543 | 0.00095456 | 0.00921856 | 3.30358538 | 0.41 | eqtlPRS |
| rs1973489 | 5 | 1381260 | G | A | 0.0348463 | 0.00095771 | 0.010551 | 3.30265378 | 0.2382 | eqtlPRS |
| rs117777471 | 16 | 56660685 | A | G | -0.0526878 | 0.00095893 | 0.0159548 | -3.3023165 | 0.1125 | eqtlPRS |
| rs72730672 | 15 | 79256314 | C | T | 0.0639454 | 0.00095899 | 0.019364 | 3.30228259 | 0.1012 | eqtlPRS |
| rs664220 | 1 | 234786161 | G | A | 0.0325687 | 0.00095948 | 0.00986289 | 3.30214572 | 0.3088 | eqtlPRS |
| rs340909 | 9 | 6128646 | A | G | -0.0328141 | 0.00096132 | 0.00993883 | -3.3016059 | 0.3119 | eqtlPRS |
| rs13218316 | 6 | 112083034 | C | T | -0.0402347 | 0.00096165 | 0.0121867 | -3.3015254 | 0.1789 | eqtlPRS |
| rs6560407 | 9 | 71486972 | G | A | -0.0707005 | 0.00096369 | 0.0214184 | -3.3009235 | 0.04397 | eqtlPRS |
| rs17469450 | 15 | 77619544 | T | C | 0.0766424 | 0.00096466 | 0.0232205 | 3.30063521 | 0.03783 | eqtlPRS |
| rs4755346 | 11 | 45917780 | C | T | 0.0346259 | 0.00096519 | 0.0104912 | 3.30047087 | 0.2781 | eqtlPRS |
| rs12023499 | 1 | 155031376 | C | T | 0.0374293 | 0.00096617 | 0.0113415 | 3.3002072 | 0.1902 | eqtlPRS |
| rs76305941 | 7 | 138598084 | T | G | 0.099295 | 0.00096882 | 0.0300946 | 3.29942913 | 0.01329 | eqtlPRS |
| rs11712099 | 3 | 196316530 | G | A | 0.0334445 | 0.00097023 | 0.0101377 | 3.29902246 | 0.4202 | eqtlPRS |
| rs8013647 | 14 | 35822984 | T | C | 0.0376923 | 0.00097063 | 0.0114257 | 3.2989051 | 0.2147 | eqtlPRS |
| rs41447547 | 11 | 1101864 | C | A | -0.0634538 | 0.00097452 | 0.0192413 | -3.2977917 | 0.0818 | eqtlPRS |
| rs10403012 | 19 | 39330058 | A | G | -0.0305295 | 0.00097453 | 0.0092576 | -3.297777 | 0.3957 | eqtlPRS |
| rs7635895 | 3 | 126799624 | A | G | -0.0464134 | 0.00097637 | 0.0140764 | -3.2972493 | 0.1697 | eqtlPRS |
| rs10409238 | 19 | 21888264 | T | C | 0.0320668 | 0.00098141 | 0.00972958 | 3.29580516 | 0.3579 | eqtlPRS |
| rs62204582 | 20 | 43376268 | G | A | 0.0946485 | 0.00098626 | 0.02873 | 3.29441351 | 0.03476 | eqtlPRS |
| rs1549697 | 19 | 376008 | C | T | -0.0371858 | 0.00098684 | 0.0112881 | -3.2942479 | 0.3078 | eqtlPRS |
| rs4648933 | 1 | 24435434 | C | A | -0.0319841 | 0.00098881 | 0.00971071 | -3.2936933 | 0.3425 | eqtlPRS |
| rs12476436 | 2 | 10527833 | A | C | -0.0440329 | 0.00098903 | 0.0133691 | -3.2936323 | 0.1687 | eqtlPRS |
| rs118127650 | 15 | 49372303 | C | T | -0.108411 | 0.00098973 | 0.0329172 | -3.2934454 | 0.03885 | eqtlPRS |
| rs61991614 | 14 | 77484709 | G | A | 0.0344415 | 0.00099064 | 0.0104585 | 3.29315867 | 0.2413 | eqtlPRS |
| rs12185167 | 16 | 2109953 | C | T | -0.0391618 | 0.00099202 | 0.0118932 | -3.2927892 | 0.2249 | eqtlPRS |
| rs4372812 | 19 | 697514 | C | A | -0.0351197 | 0.00099294 | 0.0106665 | -3.2925233 | 0.274 | eqtlPRS |
| rs57648594 | 17 | 20459947 | A | G | -0.0383622 | 0.00099424 | 0.0116526 | -3.292158 | 0.2679 | eqtlPRS |
| rs730275 | 20 | 58254093 | A | G | 0.0451062 | 0.00099881 | 0.0137065 | 3.290862 | 0.135 | eqtlPRS |
| rs12404496 | 1 | 47465048 | A | C | 0.0315139 | 0.0010013 | 0.00957824 | 3.2901556 | 0.4264 | eqtlPRS |
| rs11863728 | 16 | 56964942 | G | A | -0.0331018 | 0.00100157 | 0.0100611 | -3.2900776 | 0.2965 | eqtlPRS |
| rs1878026 | 10 | 127710314 | C | T | 0.0483929 | 0.00100323 | 0.0147108 | 3.28961715 | 0.1125 | eqtlPRS |
| rs28411352 | 1 | 38278579 | C | T | -0.0356502 | 0.00100515 | 0.0108389 | -3.2890976 | 0.2546 | eqtlPRS |
| rs73004911 | 11 | 119503612 | A | G | -0.0806386 | 0.00100538 | 0.0245175 | -3.2890221 | 0.04499 | eqtlPRS |
| rs116892768 | 22 | 25287815 | G | A | 0.115442 | 0.00100573 | 0.0351004 | 3.28890839 | 0.0317 | eqtlPRS |
| rs4609758 | 14 | 23262787 | C | T | -0.0573463 | 0.00100574 | 0.0174362 | -3.2889219 | 0.08793 | eqtlPRS |
| rs139666113 | 12 | 123940755 | A | G | 0.133403 | 0.00100703 | 0.0405657 | 3.28856645 | 0.02863 | eqtlPRS |
| rs6780355 | 3 | 150417593 | A | G | 0.0301422 | 0.0010075 | 0.00916617 | 3.28841817 | 0.455 | eqtlPRS |
| rs76588278 | 1 | 207044272 | G | A | -0.17167 | 0.00100886 | 0.0522104 | -3.2880422 | 0.01431 | eqtlPRS |
| rs4744178 | 9 | 95890162 | T | C | -0.0332696 | 0.00101133 | 0.0101205 | -3.2873475 | 0.2843 | eqtlPRS |
| rs7518754 | 1 | 42356437 | T | C | 0.0925518 | 0.00101475 | 0.028162 | 3.28640722 | 0.02863 | eqtlPRS |
| rs9749388 | 19 | 48265232 | C | A | -0.0602566 | 0.00101629 | 0.0183375 | -3.2859768 | 0.07055 | eqtlPRS |
| rs72732580 | 5 | 36785541 | A | G | -0.0338537 | 0.0010189 | 0.0103047 | -3.2852679 | 0.2352 | eqtlPRS |
| rs1313761 | 14 | 25609247 | C | T | 0.0399846 | 0.00101919 | 0.0121712 | 3.28518141 | 0.1534 | eqtlPRS |
| rs150557869 | 12 | 122081403 | G | A | -0.0928175 | 0.00101954 | 0.0282543 | -3.2850752 | 0.04601 | eqtlPRS |
| rs35579338 | 9 | 36739204 | T | G | 0.0397096 | 0.00102008 | 0.0120884 | 3.28493432 | 0.1881 | eqtlPRS |
| rs56122389 | 12 | 53212107 | G | A | 0.112378 | 0.00102112 | 0.0342133 | 3.28462908 | 0.02045 | eqtlPRS |
| rs61990526 | 14 | 71285675 | A | G | 0.0351124 | 0.0010238 | 0.0106923 | 3.28389589 | 0.2454 | eqtlPRS |
| rs36046157 | 22 | 24169370 | T | G | -0.0869629 | 0.00102415 | 0.0264823 | -3.2838122 | 0.03374 | eqtlPRS |
| rs141563399 | 11 | 36766822 | C | T | -0.134674 | 0.00102525 | 0.0410154 | -3.2834984 | 0.01125 | eqtlPRS |
| rs2843633 | 21 | 38328886 | A | C | 0.0314238 | 0.00103173 | 0.00957539 | 3.28172534 | 0.3937 | eqtlPRS |
| rs113870709 | 14 | 35931135 | A | G | -0.080704 | 0.00103549 | 0.0245996 | -3.2807038 | 0.04601 | eqtlPRS |
| rs7581528 | 2 | 47240149 | G | A | -0.0577998 | 0.001038 | 0.0176218 | -3.2800168 | 0.09202 | eqtlPRS |
| rs4905125 | 14 | 94442455 | T | C | 0.0370714 | 0.00103924 | 0.0113034 | 3.27966806 | 0.2014 | eqtlPRS |
| rs9680017 | 20 | 56518646 | A | C | -0.034208 | 0.00104715 | 0.0104371 | -3.2775388 | 0.274 | eqtlPRS |
| rs6794342 | 3 | 141766643 | A | G | -0.0318609 | 0.00104828 | 0.00972187 | -3.2772399 | 0.3282 | eqtlPRS |
| rs1647385 | 15 | 57138501 | A | G | 0.038651 | 0.00104839 | 0.0117939 | 3.27720262 | 0.1933 | eqtlPRS |
| rs11618615 | 13 | 51523052 | A | G | -0.0534497 | 0.00104878 | 0.0163101 | -3.2770921 | 0.08487 | eqtlPRS |
| rs216904 | 12 | 6106018 | T | C | -0.0305985 | 0.00105358 | 0.00934072 | -3.2758181 | 0.362 | eqtlPRS |
| rs1965067 | 12 | 11427558 | G | T | 0.0729609 | 0.00105534 | 0.0222758 | 3.27534365 | 0.04499 | eqtlPRS |
| rs6691659 | 1 | 156065345 | A | G | 0.039776 | 0.00105678 | 0.0121455 | 3.2749578 | 0.1503 | eqtlPRS |
| rs231597 | 19 | 36236120 | G | A | 0.0708199 | 0.00105756 | 0.021626 | 3.27475724 | 0.05112 | eqtlPRS |
| rs8109438 | 19 | 13963848 | A | G | 0.0435432 | 0.00107014 | 0.0133102 | 3.27141591 | 0.1697 | eqtlPRS |
| rs7639036 | 3 | 112928762 | A | C | -0.0372496 | 0.00107048 | 0.0113867 | -3.2713253 | 0.2168 | eqtlPRS |
| rs75086830 | 5 | 37560455 | C | T | -0.0395322 | 0.00107358 | 0.0120875 | -3.2705026 | 0.1575 | eqtlPRS |
| rs2053395 | 2 | 42296499 | G | A | -0.0321052 | 0.00107845 | 0.00982047 | -3.2692122 | 0.4652 | eqtlPRS |
| rs1808860 | 4 | 89234709 | T | C | 0.0302382 | 0.00107883 | 0.00924965 | 3.26911829 | 0.4744 | eqtlPRS |
| rs13054018 | 22 | 26144627 | C | T | -0.0489278 | 0.00108432 | 0.0149732 | -3.2676916 | 0.1053 | eqtlPRS |
| rs55830103 | 20 | 57465943 | T | G | -0.042602 | 0.00108635 | 0.0130395 | -3.2671498 | 0.1994 | eqtlPRS |
| rs17556675 | 18 | 32291160 | G | A | 0.0540733 | 0.00108655 | 0.0165508 | 3.26711096 | 0.07771 | eqtlPRS |
| rs72895440 | 2 | 69263595 | G | A | -0.09268 | 0.00109773 | 0.0283928 | -3.2642078 | 0.03885 | eqtlPRS |
| rs7162408 | 15 | 52403526 | C | T | 0.0318647 | 0.00109816 | 0.00976219 | 3.26409341 | 0.3241 | eqtlPRS |
| rs601863 | 11 | 65646557 | A | C | 0.0373627 | 0.00109956 | 0.0114479 | 3.26371649 | 0.1902 | eqtlPRS |
| rs61781966 | 1 | 20938889 | A | C | 0.0633503 | 0.00110191 | 0.019414 | 3.26312455 | 0.05419 | eqtlPRS |
| rs2804922 | 6 | 13572686 | C | T | 0.0321093 | 0.00110415 | 0.00984178 | 3.26255007 | 0.3006 | eqtlPRS |
| rs6717619 | 2 | 45571350 | C | T | 0.0307442 | 0.00110531 | 0.00942421 | 3.26225753 | 0.3937 | eqtlPRS |
| rs9870893 | 3 | 16863267 | T | G | 0.0307041 | 0.00111431 | 0.00941856 | 3.25995694 | 0.3589 | eqtlPRS |
| rs66560125 | 7 | 107370034 | G | A | -0.0341689 | 0.00111461 | 0.0104816 | -3.2598935 | 0.2648 | eqtlPRS |
| rs1831872 | 20 | 49001560 | G | A | -0.0511455 | 0.00111649 | 0.0156917 | -3.2593983 | 0.1033 | eqtlPRS |
| rs79032535 | 5 | 78563558 | C | T | 0.114216 | 0.00111711 | 0.0350438 | 3.25923559 | 0.01431 | eqtlPRS |
| rs11085758 | 19 | 11185014 | A | G | 0.0323668 | 0.00111797 | 0.00993145 | 3.25902059 | 0.41 | eqtlPRS |
| rs11607839 | 11 | 119680383 | A | C | -0.0321985 | 0.00111934 | 0.00988087 | -3.2586705 | 0.3303 | eqtlPRS |
| rs202041 | 6 | 13288425 | A | C | 0.0318958 | 0.00112075 | 0.00978903 | 3.25832079 | 0.3027 | eqtlPRS |
| rs143798894 | 21 | 27113589 | C | T | -0.119572 | 0.00112419 | 0.0367073 | -3.2574447 | 0.01738 | eqtlPRS |
| rs62342731 | 4 | 178426345 | C | T | 0.0337969 | 0.0011251 | 0.010376 | 3.25721858 | 0.2761 | eqtlPRS |
| rs12705091 | 7 | 100472380 | C | T | 0.0380458 | 0.00112579 | 0.0116811 | 3.25703915 | 0.1912 | eqtlPRS |
| rs10131273 | 14 | 65692342 | A | C | 0.0338722 | 0.00112697 | 0.0104006 | 3.25675442 | 0.2423 | eqtlPRS |
| rs144669720 | 1 | 40226630 | C | T | 0.105251 | 0.00112934 | 0.0323238 | 3.25614563 | 0.02352 | eqtlPRS |
| rs73581971 | 14 | 21264372 | G | A | 0.0317919 | 0.00112967 | 0.00976388 | 3.25607238 | 0.3722 | eqtlPRS |
| rs75406176 | 10 | 95855835 | T | C | -0.0844955 | 0.0011316 | 0.025954 | -3.2555868 | 0.04397 | eqtlPRS |
| rs3773855 | 3 | 196867871 | G | A | 0.0344918 | 0.00113211 | 0.0105951 | 3.25544827 | 0.2699 | eqtlPRS |
| rs11651717 | 17 | 19776516 | C | T | -0.0349953 | 0.0011337 | 0.010751 | -3.2550739 | 0.2751 | eqtlPRS |
| rs73212635 | 8 | 17783013 | T | G | 0.0582148 | 0.00113419 | 0.0178851 | 3.25493288 | 0.1043 | eqtlPRS |
| rs79393352 | 3 | 45971018 | G | A | 0.0722828 | 0.0011366 | 0.0222113 | 3.2543255 | 0.04908 | eqtlPRS |
| rs72854332 | 1 | 6357659 | G | A | 0.122507 | 0.00113669 | 0.0376446 | 3.25430473 | 0.01738 | eqtlPRS |
| rs3847193 | 9 | 133769169 | G | A | 0.0342128 | 0.0011372 | 0.0105135 | 3.25417796 | 0.2454 | eqtlPRS |
| rs12902823 | 15 | 100352218 | A | G | -0.0300452 | 0.00113767 | 0.00923314 | -3.2540609 | 0.498 | eqtlPRS |
| rs11236433 | 11 | 75161525 | C | T | 0.141387 | 0.00113842 | 0.0434518 | 3.25388131 | 0.02147 | eqtlPRS |
| rs10740109 | 10 | 65040139 | A | C | 0.0329762 | 0.00114241 | 0.0101375 | 3.25289273 | 0.2771 | eqtlPRS |
| rs16950051 | 12 | 120729505 | G | A | 0.0603389 | 0.0011438 | 0.0185513 | 3.25254295 | 0.05828 | eqtlPRS |
| rs4752701 | 10 | 124243587 | A | G | 0.0838666 | 0.00114617 | 0.0257897 | 3.25194167 | 0.0409 | eqtlPRS |
| rs11774633 | 8 | 131190243 | C | T | 0.0296992 | 0.00114655 | 0.009133 | 3.25185591 | 0.4888 | eqtlPRS |
| rs2489052 | 10 | 102405390 | C | A | 0.0300091 | 0.00114692 | 0.00922856 | 3.25176409 | 0.4172 | eqtlPRS |
| rs58450802 | 7 | 98743717 | G | A | 0.0358823 | 0.00114717 | 0.0110349 | 3.25171048 | 0.2249 | eqtlPRS |
| rs73229742 | 4 | 40060168 | A | C | 0.0574517 | 0.00115029 | 0.0176724 | 3.250928 | 0.06851 | eqtlPRS |
| rs111361623 | 11 | 125948708 | G | A | -0.0859636 | 0.00115072 | 0.0264436 | -3.2508282 | 0.0409 | eqtlPRS |
| rs72755478 | 15 | 70939320 | C | T | 0.0485353 | 0.00115252 | 0.0149322 | 3.25037838 | 0.09714 | eqtlPRS |
| rs9904970 | 17 | 61201358 | G | A | 0.0319745 | 0.00115475 | 0.00983883 | 3.24982747 | 0.3681 | eqtlPRS |
| rs150353 | 15 | 89928189 | T | G | -0.0297439 | 0.00115586 | 0.00915322 | -3.2495559 | 0.4325 | eqtlPRS |
| rs1405830 | 7 | 141134375 | C | T | 0.0600062 | 0.00115776 | 0.0184686 | 3.24909306 | 0.07669 | eqtlPRS |
| rs3819091 | 18 | 12364185 | C | A | 0.0898941 | 0.00115875 | 0.0276696 | 3.24883988 | 0.03579 | eqtlPRS |
| rs41291502 | 1 | 46476864 | G | A | -0.0693245 | 0.00115875 | 0.0213382 | -3.2488448 | 0.0501 | eqtlPRS |
| rs7483870 | 11 | 976019 | G | A | -0.0373956 | 0.00115921 | 0.0115108 | -3.2487403 | 0.2311 | eqtlPRS |
| rs111927578 | 19 | 2072843 | G | A | -0.147222 | 0.00115951 | 0.0453179 | -3.2486501 | 0.01636 | eqtlPRS |

**Supplementary Table 5 - Association between PRS and lung cancer in the TCGA case cohorts. Association in lung cancer versus all other cancers.**

| **TCGA cohort** | sm.PRS | | | eQTL.PRS | | | combined.PRS | | |
| --- | --- | --- | --- | --- | --- | --- | --- | --- | --- |
|  | OR | SE | Pval | OR | SE | Pval | OR | SE | Pval |
| All cohorts included (781 lung cancer, 7555 other cancers) | 1.185 | 0.039 | 1.12E-05 | 1.263 | 0.039 | 2.23E-09 | 1.256 | 0.039 | 3.90E-09 |
| Smoking related cancers (BLCA, HNSC) removed  (781 lung cancer, 6822 other cancers) | 1.208 | 0.039 | 1.51E-06 | 1.270 | 0.039 | 1.38E-09 | 1.270 | 0.039 | 1.02E-09 |
| All cohorts with smoking information with smoking adjustment  (762 lung cancer, 1390 other cancers) | 1.108 | 0.050 | 3.83E-02 | 1.209 | 0.051 | 1.80E-04 | 1.172 | 0.050 | 1.53E-03 |

**Supplementary Table 6 - Association of tobacco-smoking PRS (sm-PRS) with mutation load and SBS4 by TCGA cohort**

| **TE** | **seTE** | **pval** | **Fdr** | **PRS** | **Cohort** | **Outcome** |
| --- | --- | --- | --- | --- | --- | --- |
| 0,17 | 0,05 | 2.36E-03 | 3.06E-02 | sm-PRS | LUAD | Mutation load |
| 0,27 | 0,09 | 2.45E-03 | 3.06E-02 | sm-PRS | ESCA | Mutation load |
| 0,28 | 0,10 | 7.25E-03 | 6.04E-02 | sm-PRS | STAD | Mutation load |
| 0,33 | 0,13 | 1.14E-02 | 7.13E-02 | sm-PRS | PRAD | Mutation load |
| 0,48 | 0,23 | 4.20E-02 | 2.10E-01 | sm-PRS | LGG | Mutation load |
| -0,05 | 0,03 | 1.18E-01 | 4.92E-01 | sm-PRS | KIRC | Mutation load |
| 0,06 | 0,05 | 2.20E-01 | 7.00E-01 | sm-PRS | OV* | Mutation load |
| -0,30 | 0,25 | 2.37E-01 | 7.00E-01 | sm-PRS | THYM | Mutation load |
| 0,26 | 0,23 | 2.65E-01 | 7.00E-01 | sm-PRS | CESC* | Mutation load |
| 0,42 | 0,39 | 2.85E-01 | 7.00E-01 | sm-PRS | GBM | Mutation load |
| 0,09 | 0,09 | 3.08E-01 | 7.00E-01 | sm-PRS | LIHC | Mutation load |
| 0,03 | 0,03 | 3.74E-01 | 7.79E-01 | sm-PRS | KIRP | Mutation load |
| 0,03 | 0,04 | 4.28E-01 | 7.94E-01 | sm-PRS | LUSC | Mutation load |
| -0,04 | 0,06 | 4.77E-01 | 7.94E-01 | sm-PRS | TGCT* | Mutation load |
| 0,11 | 0,16 | 5.01E-01 | 7.94E-01 | sm-PRS | UCEC | Mutation load |
| 0,02 | 0,04 | 5.40E-01 | 7.94E-01 | sm-PRS | THCA | Mutation load |
| 0,07 | 0,12 | 5.61E-01 | 7.94E-01 | sm-PRS | BRCA* | Mutation load |
| 0,04 | 0,07 | 5.72E-01 | 7.94E-01 | sm-PRS | SKCM | Mutation load |
| -0,03 | 0,07 | 6.32E-01 | 8.32E-01 | sm-PRS | BLCA | Mutation load |
| -0,09 | 0,31 | 7.64E-01 | 9.49E-01 | sm-PRS | LAML | Mutation load |
| -0,02 | 0,06 | 7.99E-01 | 9.49E-01 | sm-PRS | HNSC | Mutation load |
| 0,42 | 2,18 | 8.49E-01 | 9.49E-01 | sm-PRS | READ | Mutation load |
| -0,01 | 0,13 | 9.07E-01 | 9.49E-01 | sm-PRS | COAD | Mutation load |
| 0,00 | 0,04 | 9.45E-01 | 9.49E-01 | sm-PRS | PCPG | Mutation load |
| -0,01 | 0,13 | 9.49E-01 | 9.49E-01 | sm-PRS | SARC | Mutation load |
| 0,20 | 0,07 | 3.71E-03 | 1.48E-02 | sm-PRS | LUAD | SBS4 |
| 0,10 | 0,04 | 1.92E-02 | 3.84E-02 | sm-PRS | LUSC | SBS4 |
| 0,19 | 0,16 | 2.53E-01 | 3.37E-01 | sm-PRS | HNSC | SBS4 |
| -0,01 | 0,17 | 9.68E-01 | 9.68E-01 | sm-PRS | LIHC | SBS4 |

* gender not included as covariate

In SBS4 analysis, only cohorts where SBS4 can be attributed to more than 100 cases were considered.

Quasipoisson model adjusted by the covariates (age,gender,tumour_purity(cat),Ancestory_PC1-5)

**Supplementary Figures**

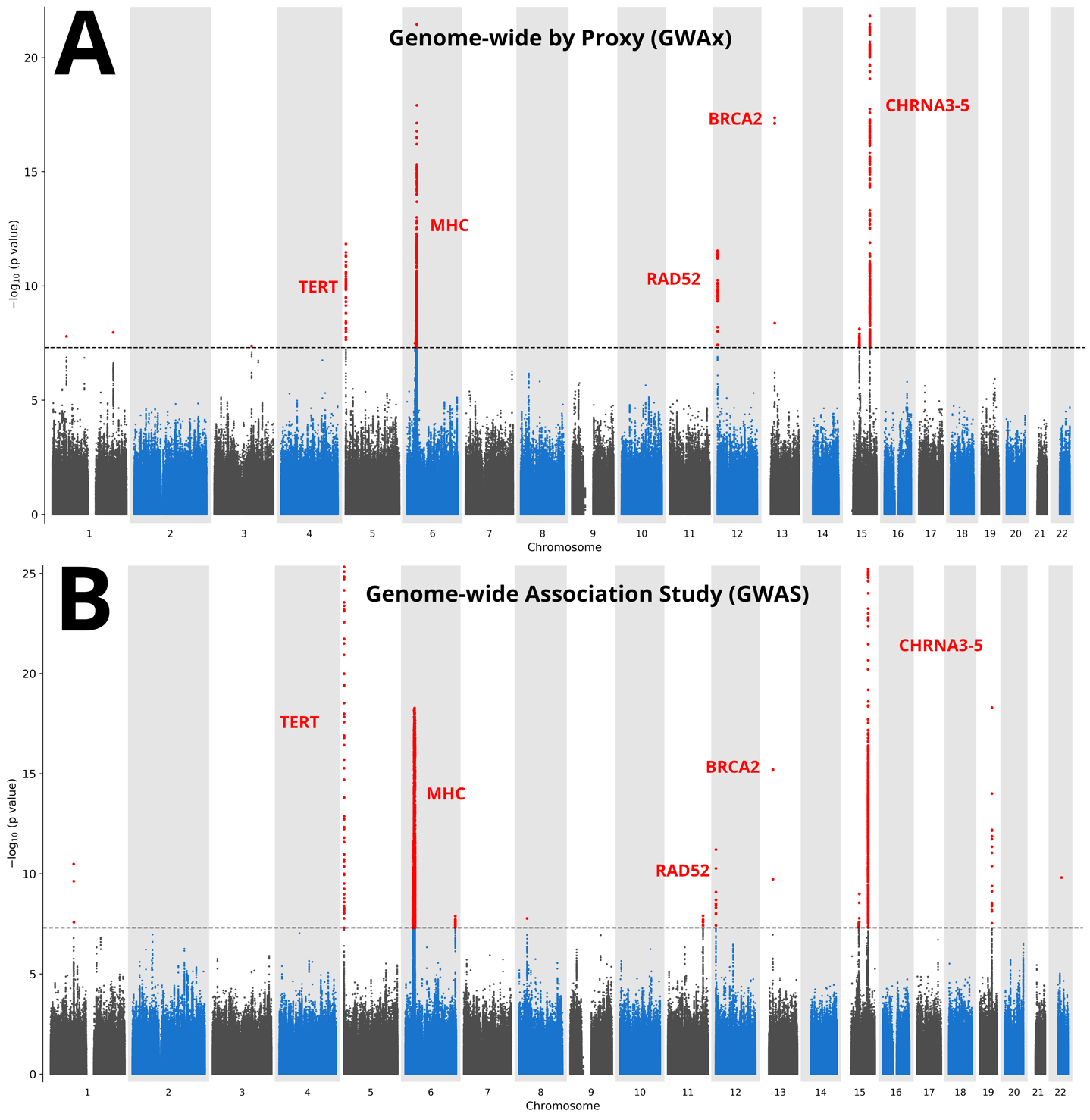

***Supplementary Figure 2: Visual validation of genome-wide by proxy method by Manhattan plot compared to the lung cancer genome-wide association study.***

To visually display the findings of both the genome-wide association by proxy (**A**) and the lung cancer genome-wide study (**B**), summary statistics from both scans were plotted as Manhattan plots (<https://github.com/pgxcentre/manhattan_generator>) with the common peaks highlighted with likely gene targets. Many loci for case/control GWAS replicated out in the proxy case/proxy control GWAx method. This included the genes implicated in these regions including *TERT*, the major histocompatibility complex (MHC) locus, *RAD52*, *BRCA2* and *CHRNA3-5* locus.

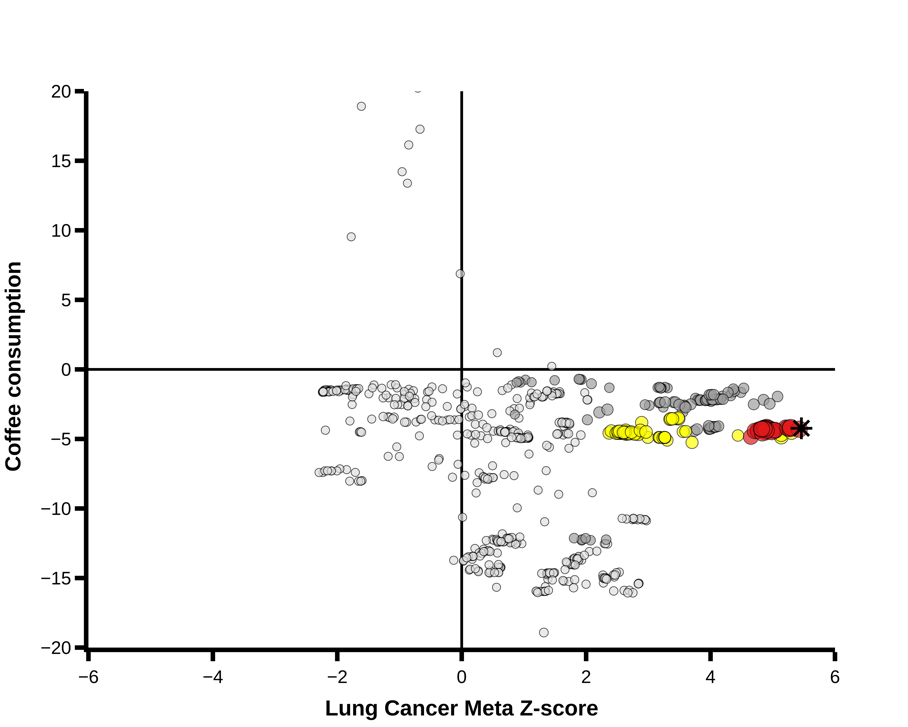

**A**

Colocalisation between coffee consumption and LC PP = 0.003%

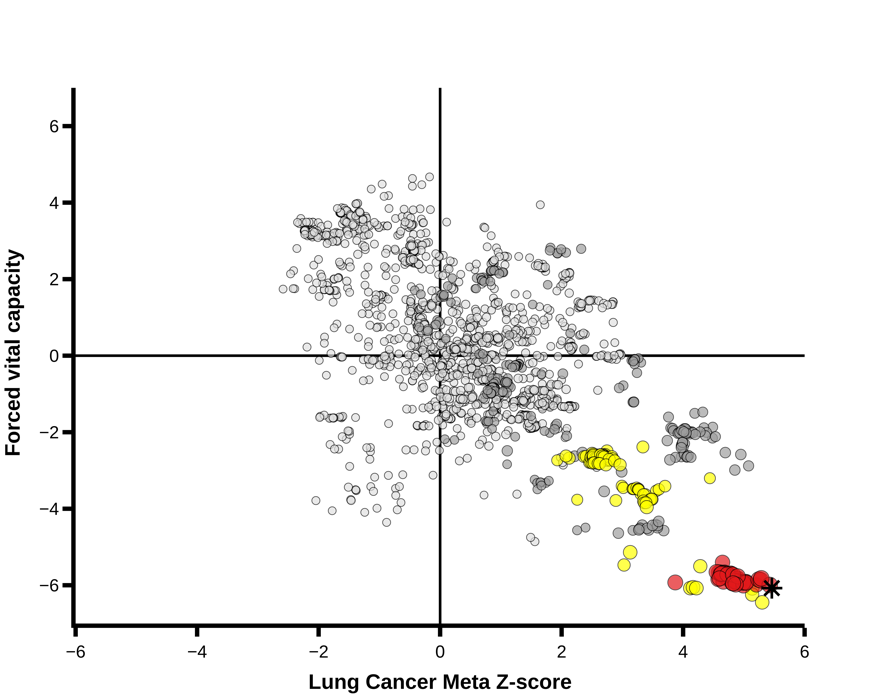

**B**

Colocalisation between FEV and LC PP = 97.05%

***Supplementary Figure 3*:** ***Z-statistic plots for variants associated with traits at 15q24(CYP1A1) compared to lung cancer***

Z-statistics for coffee consumption (**A**) and forced vital capacity (FVC) (**B**) were compared to z-statistic from the lung cancer meta-analysis results. The posterior probability (PP) for colocalisation was calculated using COLOC.

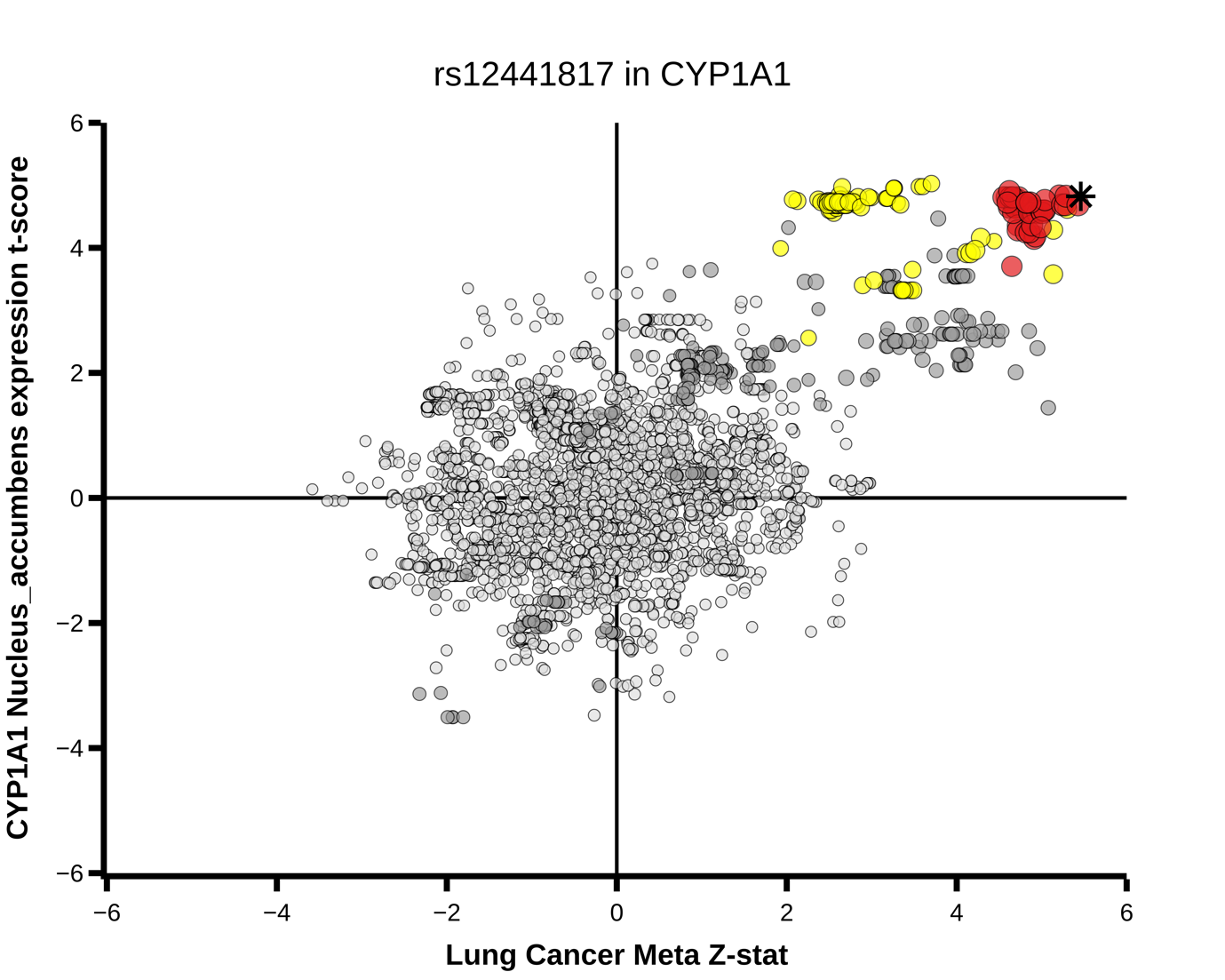

Colocalisation between *CYP1A1* nucleus accumbens expression and LC PP = 70.26%

***Supplementary Figure 4: CYPA1A expression in the nucleus accumbens***

*Co-localisation between lung cancer (x axis) and CYPA1A1 nucleus accumbens expression. The variant rs12441817 and nucleus accumbens eQTL status were compared using COLOC for colocalisation. The posterior probability (PP) for hypothesis 4 (Colocalisation with lung cancer and CYPA1A, by rs12441817) was 70.26%.*

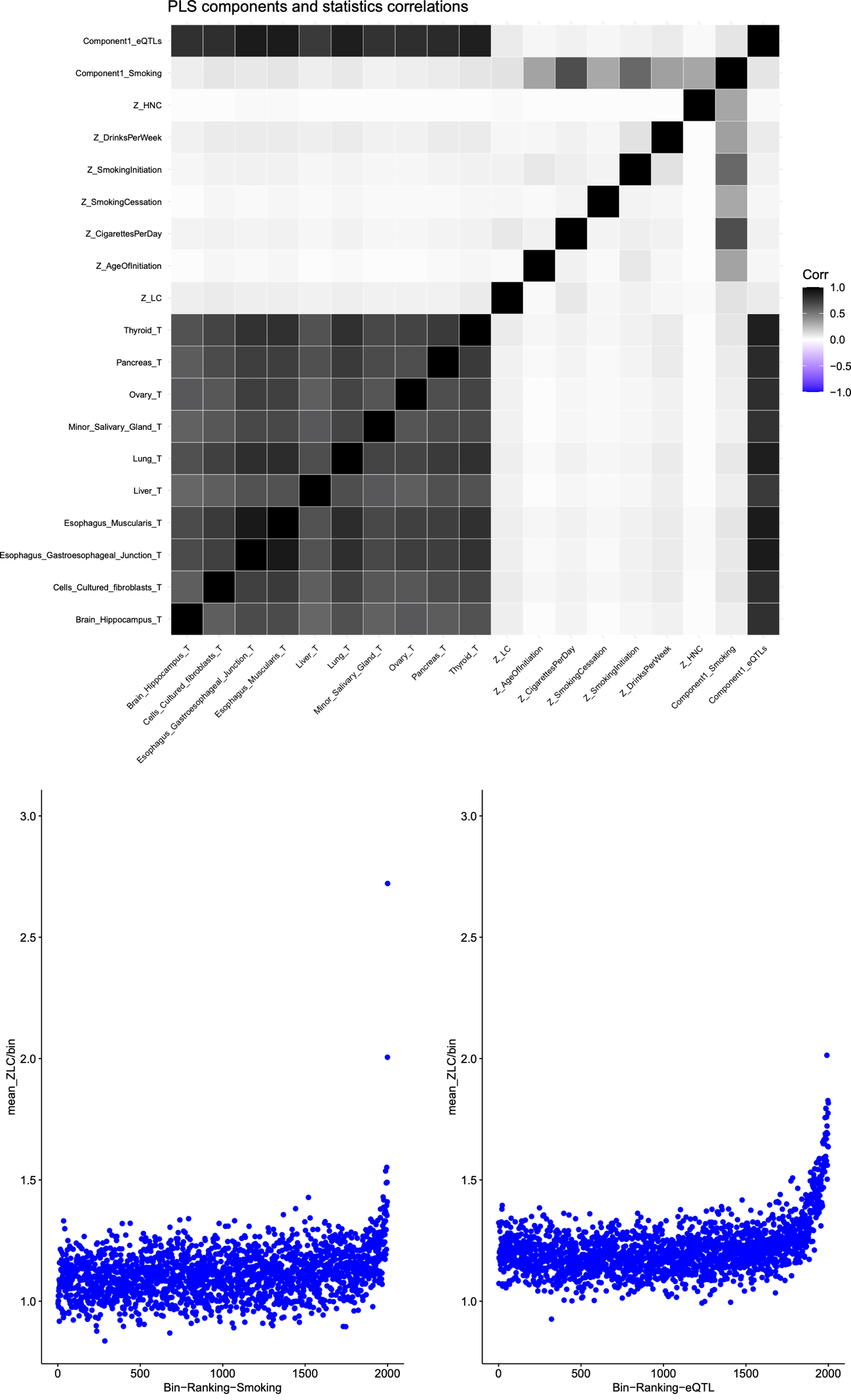

***Supplementary Figure 5*: *Partial least squares of mean z-scores for lung cancer for the polygenic risk scores construction and correlation across smoking traits and eQTLs***

Correlation plot between the first PLS components resulting from the PLS based on eQTLs and smoking-related traits statistics with various smoking-related traits (Z scores statistics resulted from GWAS on smoking related traits: age of initiation, cigarettes per day, smoking cessation, smoking initiation, drinking and head and neck cancer and eQTL T statistics for different tissues). The first eQTL component (Component1_eQTLs) is highly correlated with being an eQTL in multiple tissue types (including lung), whereas the smoking component (Component1_smoking) appears to be correlated with smoking traits.

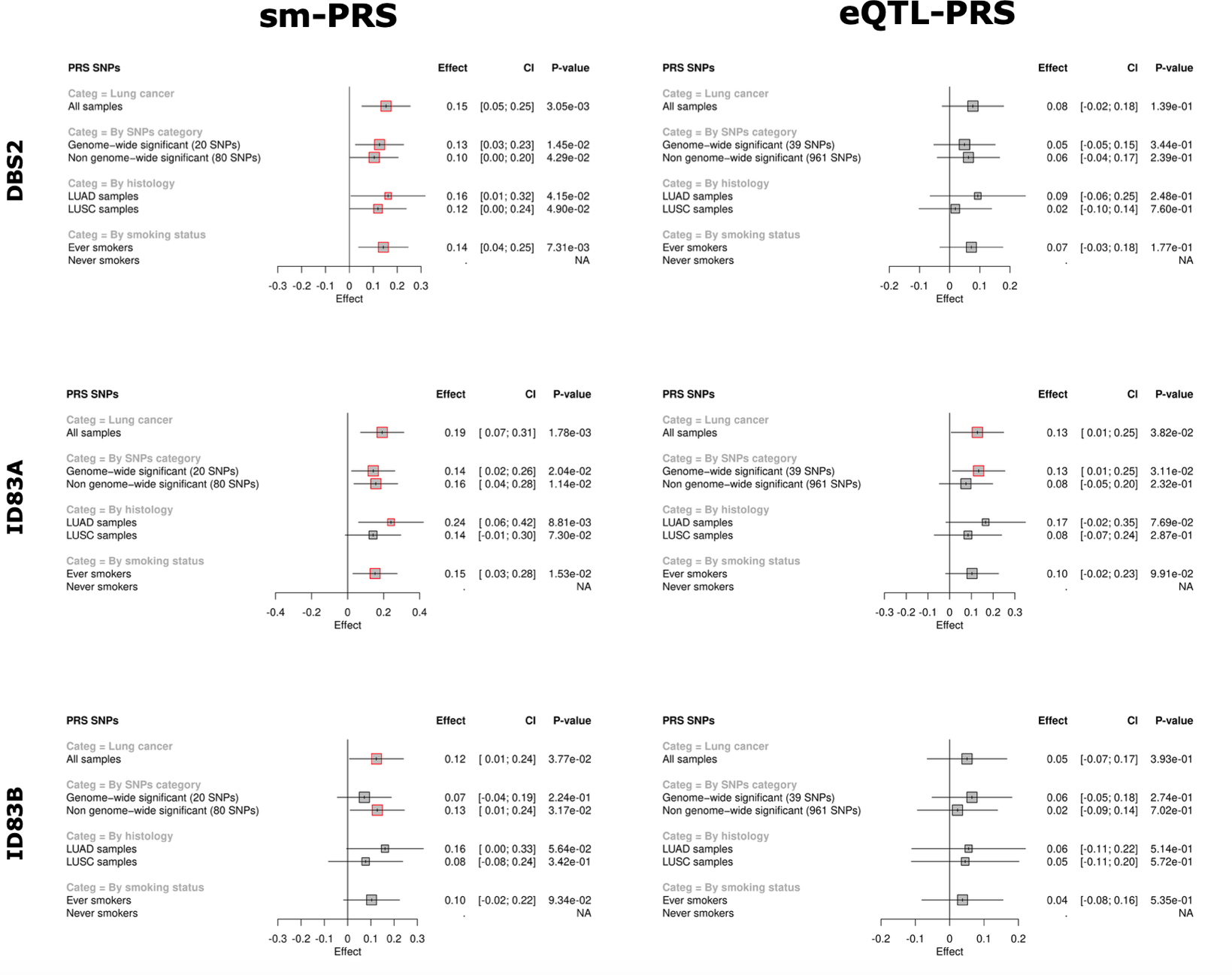

***Supplementary Figure 6: The smPRS and eQTLPRS associations with mutational signatures related to smoking attributed to tobacco***

This figure represents, in rows, the forest plots of sm-PRS (left column) and eQTL-PRS (right column) associations with DBS2, ID83A, ID83B. For each PRS, the association was tested: i) in all lung cancer cases when considering all SNPs in the sm (left) and eQTL (right) SNPs selection, ii) in all lung cancer cases when considering different subsets of SNPs in the PRS computation, iii) stratifying by histology, and iv) stratifying by smoking status. Gray squares correspond to the estimate resulting from the quasi-poisson models. Those squares are highlighted in red when the associated P-value is bellow 0.05.

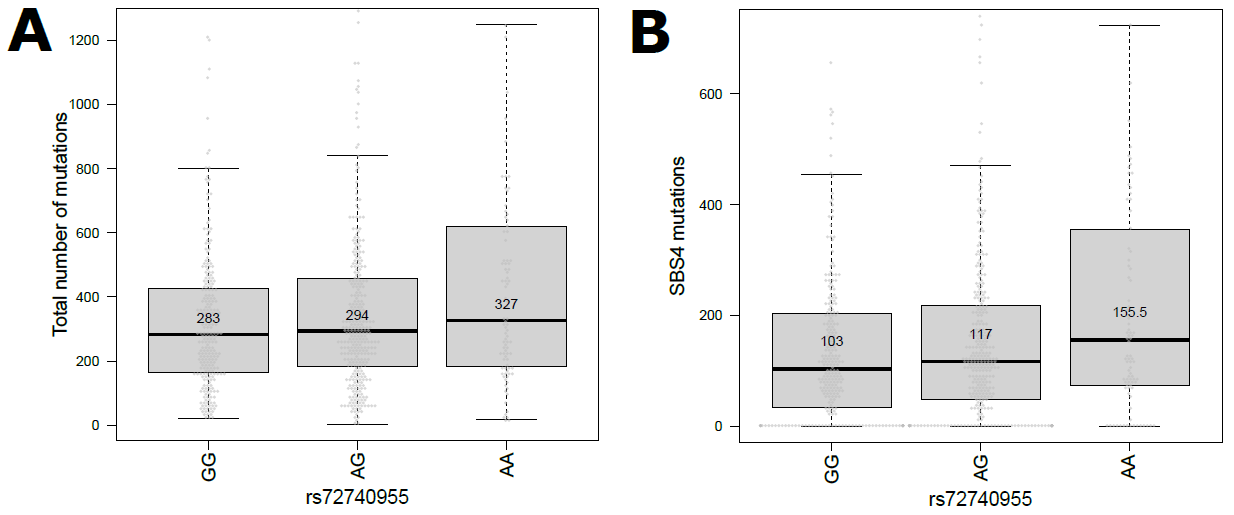

***Supplementary Figure 7: Mutational burden in lung tumours across rs72740955 genotype categories.*** Boxplots representing the distributions of the total number of mutations (**A**) and SBS4 mutations (**B**) in the three genotype categories for rs72740955 (A allele is the effect allele).

**A**

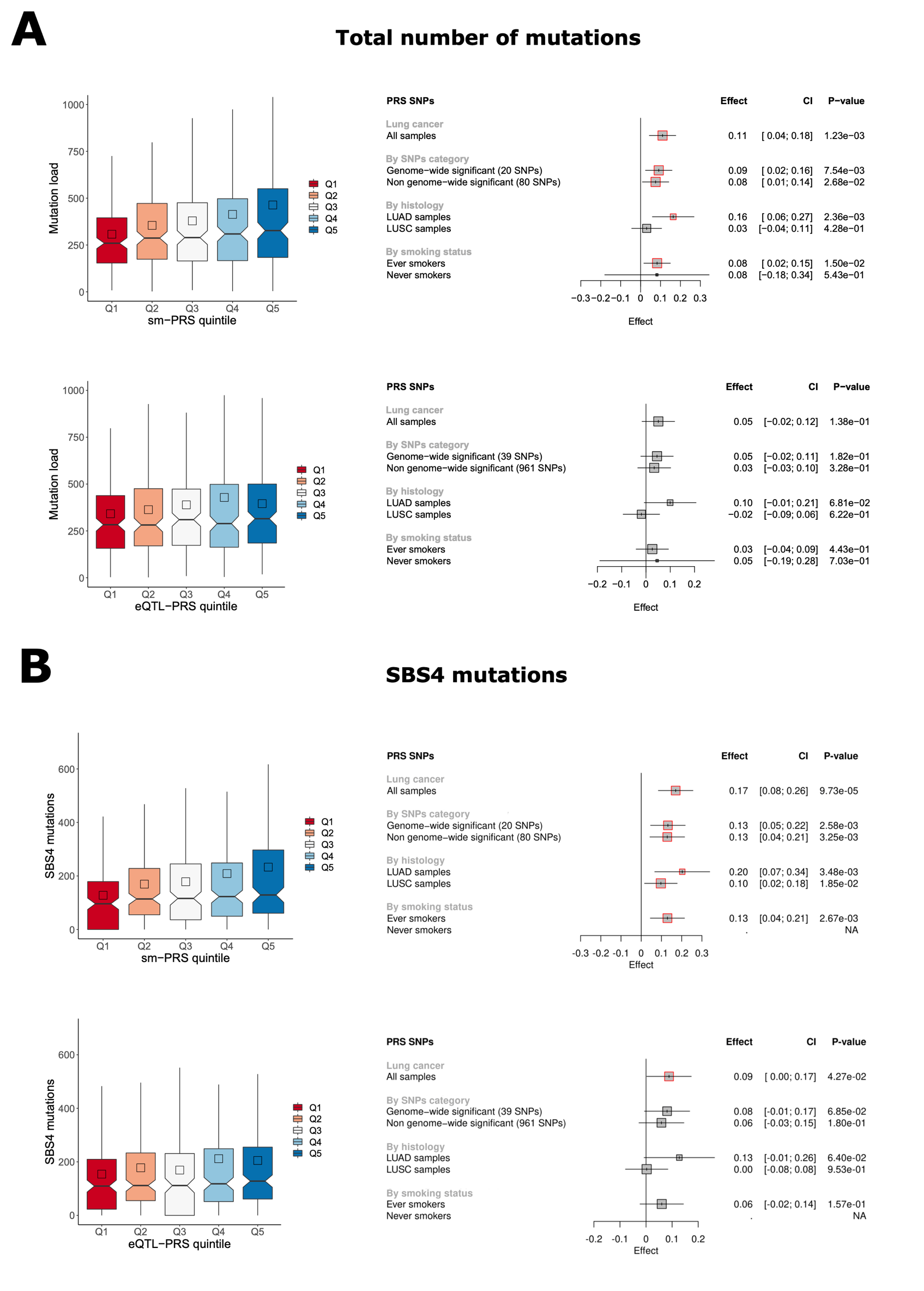

**B**

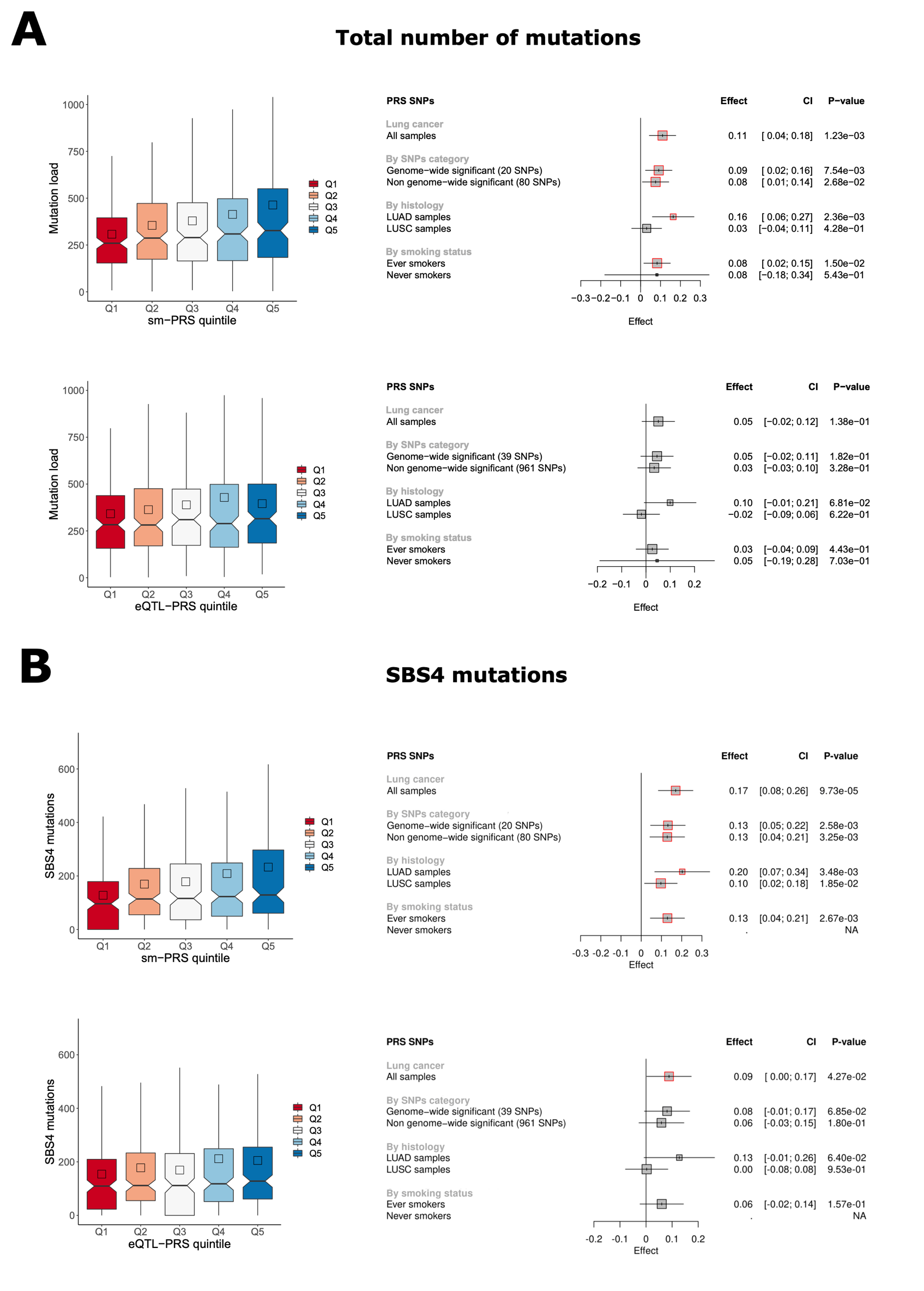

**Supplementary Figure 8: eQTLPRS associations with total number of mutations and mutations attributable to SBS4 in the TCGA cohort**

(**A**) Associations with total number of mutations. (**B**) Associations with SBS4 mutations. The left panels represent the distribution of the number of mutations in eQTL-PRS quintiles respectively. The right panels correspond, respectively, to the forest plots of eQTL-PRS associations with total mutational burden (**A**) and SBS4 mutations (**B**).

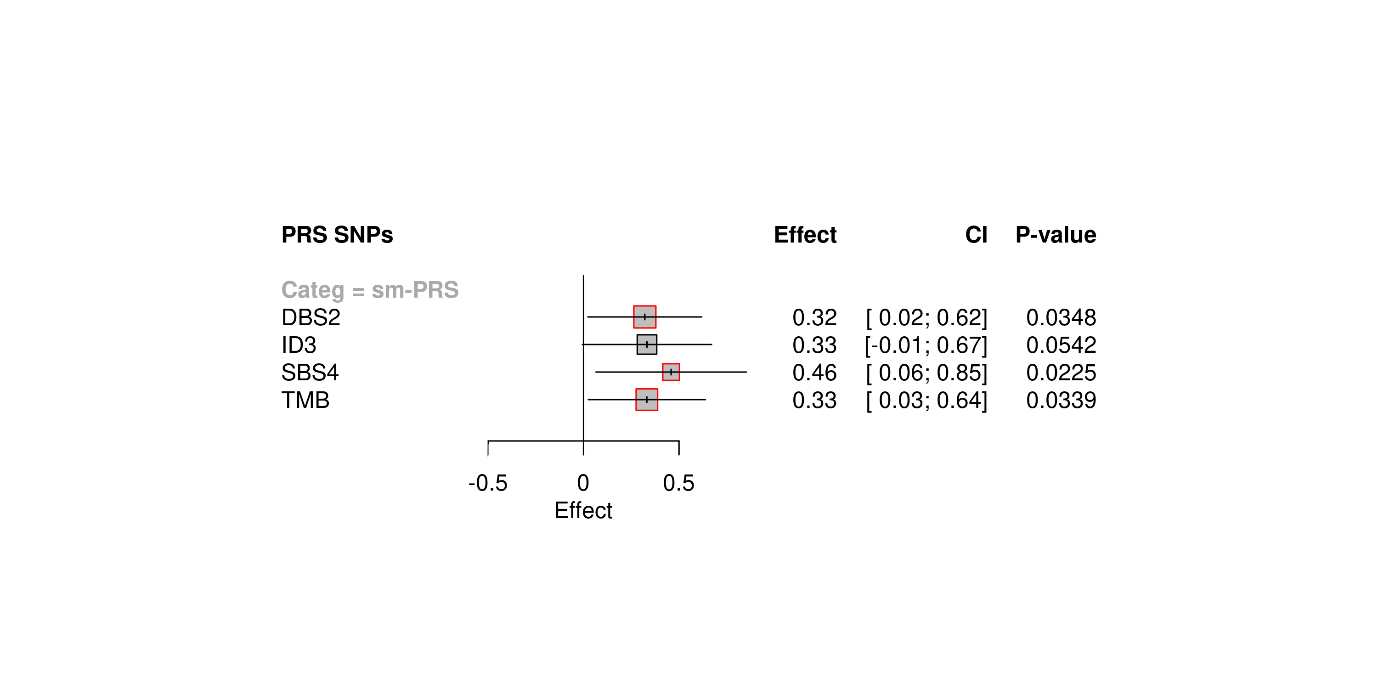

***Supplementary Figure 9: Replication analysis for the association of PRS with somatic mutational load in the GeniLuc cohort***

The forest plot represents the results of sm-PRS associations with DBS2, ID3, SBS4 and total mutation burden (TMB). Gray squares correspond to the estimate resulting from the Quasi-Poisson models. Those squares are highlighted in red when the associated P-value < 0.05.
